# Harms of short-course systemic corticosteroids among children and adolescents: a systematic review and meta-analysis of randomized controlled trials

**DOI:** 10.1101/2025.02.06.25321820

**Authors:** João Pedro Lima, Saifur R. Chowdhury, Wimonchat Tangamornsuksan, Chunjuan Zhai, Xiiajing Chu, Jessyca Matos Silva, Mahmudur Rahman Chowdhury, Humayun Kabir, Rachel Couban, Mohamed Eltorki, Gordon H Guyatt, Derek Chu

**Affiliations:** Department of Health Research Methods, Evidence, and Impact, McMaster University, Hamilton, Ontario, Canada; Department of Anesthesia, McMaster University, Hamilton, Ontario, Canada; Michael G. DeGroote Centre for Transfusion Research, Department of Medicine, McMaster University, Hamilton, Ontario, Canada; Department of Public Health, North South University, Dhaka, Bangladesh; Department of Pediatrics, McMaster University, Hamilton, ON, Canada; Department of Pediatrics, Cumming School of Medicine, University of Calgary, AB, Canada; Department of Medicine, McMaster University, Hamilton, Ontario, Canada; MAGIC Evidence Ecosystem Foundation [www.magicevidence.org], Lovisenberggata 17C, 0456 Oslo, Norway

## Abstract

**Introduction:** Short courses of systemic corticosteroids are used in the clinical management of a number of acute clinical conditions. In this systematic review and meta-analysis of randomized controlled trials we document the harms of the short-term use (≤ 14 days) of systemic corticosteroids in children and adolescents (1-18 years old) across different clinical conditions.

**Methods:** We searched MEDLINE, Embase, and the Cochrane Central Register of Controlled Trials (CENTRAL) databases from inception to January 2024 for randomized controlled trials evaluating the harms of short-course systemic corticosteroids We performed pairwise meta-analyses using the Mantel-Haenszel methods with risk difference. We assessed the certainty of evidence using the GRADE approach and subgroup analysis credibility with the ICEMAN instrument. PROSPERO registration (CRD42023400934).

**Results:** We identified 45 trials that included 6,470 children. Corticosteroids probably cause few if any serious adverse events (RD 1 fewer per 1000 [95% CI 9 fewer to 7 more]; moderate certainty), but probably do result in adverse events leading to discontinuation (RD 4 more per 1000 [95%CI 3 fewer to 11 more]; moderate certainty) compared to usual care. Corticosteroids probably increase the risk of hyperglycemia (RD 38 more per 1000 [95%CI 11 to 64 more]; moderate certainty), sleep problems (RD 15 more [95% CI 1 to 28 more]; moderate certainty), change in behavior (RD 8 more [95% CI 5 fewer to 21 more]; moderate certainty) and gastrointestinal bleeding (RD 13 more per 1000 [95% CI 3 to 23 more]; moderate certainty).

**Conclusion:** Corticosteroids likely increase the risk of hyperglycemia, sleep problems, change in behavior and gastrointestinal bleeding, but these adverse events are very seldom if ever serious.

## Introduction

### Background

Corticosteroids are immunosuppressant anti-inflammatory drugs that, over the decades, have been widely used to treat a variety of children’s illnesses including asthma, croup, and urticaria(1–4) From 2012 to 2017, corticosteroid prescriptions in Canada for children between 3 and 19 years totalled 55,000, with a prevalence of 0.9%. (5) A few systematic reviews have investigated the harms of short-term use of corticosteroids in children, some synthesized safety outcomes of corticosteroids but restricted to specific medical conditions, such as acute respiratory conditions (6), while another study restricted to only oral routes of corticosteroids.(7)

Even though the management of corticosteroids-associated adverse events (AEs) represents a significant financial burden to patients and healthcare systems (8), to our knowledge, there is no systematic review and meta-analysis available on the harms of short- course systemic corticosteroids in a pediatric population across different illnesses. AEs are frequently misreported and underrepresented in systematic reviews when compared to effective outcomes.(9, 10) Observational studies, are limited by uncertainty whether apparent adverse events are actually symptoms arising from the illness for which patients are using the steroids. (11)

A major limitation of prior individual studies, and even systematic reviews of studies of individual clinical conditions, is relatively small numbers of patients and resulting imprecision of estimates and low certainty evidence. While effectiveness of corticosteroids is likely to differ across clinical conditions, adverse effects are likely to be very similar. Therefore, we undertook a systematic review and meta-analysis of RCTs to investigate the harms of the short-term use of systemic corticosteroids in patients between 1 and 18 years old that can be generalized across a variety of pediatric clinical conditions. We limited our population to those over 1 year because of the rapid physiological and behavioural changes that occur before 1 year but not after that age.

## Methods

In reporting this study, we adhered to the 2020 PRISMA (Preferred Reporting Items for Systematic reviews and Meta-Analyses) statement (12) and PRISMA harms checklist (13). We registered this systematic review in PROSPERO (CRD42023400934).

### Literature search

With the help of a health science librarian, we searched MEDLINE, Embase, and the Cochrane Central Register of Controlled Trials (CENTRAL) databases from inception to January 2024 using a combination of keywords and Medical Subject Headings (MeSH) terms related to systemic corticosteroids and adverse events (Appendix 1). We also searched for additional eligible studies from the reference lists of eligible articles and related systematic reviews.(1, 6, 7, 14) We did not use any language restrictions.

### Eligibility criteria

We included published RCTs evaluating the harms of short-course systemic (oral, sublingual, rectal, intravenous, intramuscular and subcutaneous) corticosteroids in any condition. Eligible RCTs included non-critically ill 1- to 18-year-old children and reported on AEs during short-course use of systemic corticosteroids — dexamethasone, prednisolone, prednisone, methylprednisolone plus prednisone and hydrocortisone at any dose — versus placebo or non-steroidal standard of care. We defined AEs as any unfavorable and unintended signs, symptoms, or syndromes that occur during the period of using an investigational product, regardless of their perceived relationship to the product (15) and short-course as the use of the drug for no more than 14 days at least once daily. Given the inconsistency in the terminology used to refer to AEs in RCTs, we considered all studies that reported on ‘adverse events’, ‘adverse drug reactions’, ‘side effects’, ‘harms’, ‘safety’ or ‘toxicity’ of corticosteroids. We included studies with a population of at least 80% of children within the age criteria. We excluded trials involving perioperative corticosteroids for surgical patients, patients immunosuppressed and patients with HIV/AIDS and cancer.

### Study selection

To determine eligibility, using Covidence systematic review software (Melbourne, Australia), pairs of reviewers worked independently and in duplicate to screen titles and abstracts and, subsequently, full texts of the potentially eligible trials. Reviewers resolved disagreements by consensus or, if necessary, by consultation with a third reviewer.

### Data extraction

Pairs of reviewers independently extracted data from the eligible trials using a standardized data extraction form and resolved disagreements by discussion or, when necessary, through adjudication by a third reviewer. Reviewers collected information on trial characteristics (publication year, trial registration, designs), patient characteristics (country, age, sex, comorbidities, setting, and name of conditions corticosteroids used for), intervention descriptions (name of the interventions, route of administration, and doses), and extracted data on all the AEs reported in the trials from the intention-to-treat (ITT) population, and if not reported, in preferred order: modified-ITT, per-protocol, and as-treated population. If authors reported data at different time points, we extracted data from the latest time point at which the the intervention was still in use, and the randomization groups remain preserved.

### Risk of bias assessment

Pairs of reviewers independently performed the risk of bias assessment using a modified Cochrane tool for assessing the risk of bias in randomized trials (16), and resolved any disagreements, when necessary, by consensus or consultation with another reviewer. We considered bias arising in the randomization process, bias due to inadequate allocation concealment, bias owing to unblinding of patients, healthcare providers and outcome assessors, and bias from missing outcome data. We rated each domain as either ‘low risk of bias’, ‘probably low risk of bias’, ‘probably high risk of bias’, or ‘high risk of bias’.

### Data synthesis and analysis

We conducted pairwise meta-analysis for all AEs, in which at least two studies reported at least one event. Based on the framework proposed by Xu et al. for meta-analysis containing zero-events occurring in both single and double arms (17), we performed a fixed-effects meta-analysis of risk difference (RD) using the Mantel-Haenszel methods. We summarized the effects of interventions using RD per 1000 patients with associated 95% confidence intervals (CIs). For outcomes with ≥ 10 studies, we assessed for small study effects using visual inspection of funnel plots. We performed all analyses using the *meta* and *metafor* packages in R (version 4.03, R Foundation for Statistical Computing).(18, 19)

### Subgroup and Sensitivity Analyses

When there were at least two studies per subgroup reporting at least one event, we performed a priori subgroup analyses based on medication route (intravenous/intramuscular vs. oral route), dose (high-dose vs. low-dose using 1mg/kg/day of prednisone as the threshold), treatment duration (≤ 7 days vs >7 days) and clinical condition. We hypothesized that intravenous/intramuscular, >7 days use, and higher doses of corticosteroids are associated with a higher risk of AEs. To determine the glucocorticoid effect, we used the following corticosteroid conversions: 1 mg of dexamethasone = 26.7 mg of hydrocortisone = 5.3 mg of methylprednisolone/prednisolone = 6.7 mg of prednisone.(20, 21) We assessed the credibility of statistically significant subgroups using the Instrument for Assessing the Credibility of Effect Modification Analyses (ICEMAN).(22) To test the robustness of our analyses, we performed a sensitivity analysis using Peto’s OR.

### Certainty of evidence assessment

The Grading of Recommendations Assessment, Development and Evaluation (GRADE) approach (15) guided the certainty of evidence assessment for each outcome. The outcomes were rated as high, moderate, low, or very low certainty of evidence. Eligible trials started as high certainty of evidence and were rated down for risk of bias, inconsistency, imprecision, indirectness, or publication bias. Based on a recent GRADE guidance (23), we rated imprecision using the null effect (RD=0) as the threshold of interest. Pairs of reviewers assessed the certainty of evidence independently and, when necessary, resolved disagreements through consensus or discussion with another reviewer.

As per GRADE guidelines, we presented our results using informative statements based on the certainty of evidence and the magnitude of the effect.(24) We developed a summary of findings (SoF) table (25) using the MAGICapp (https://app.magicapp.org/). In the SoF table, AEs were grouped together based on the System Organ Classes (e.g., gastrointestinal disorders) from the Medical Dictionary for Regulatory Activities (MedDRA).(26)

## Results

Our search identified 17,905 articles. After removing duplicates, we screened titles and abstracts of 15,005 articles and excluded 14,620 articles. After full text review, 45 RCTs proved eligible (27–71). The PRISMA flow diagram summarizes the study selection process (Figure 1).

**Figure 1:**
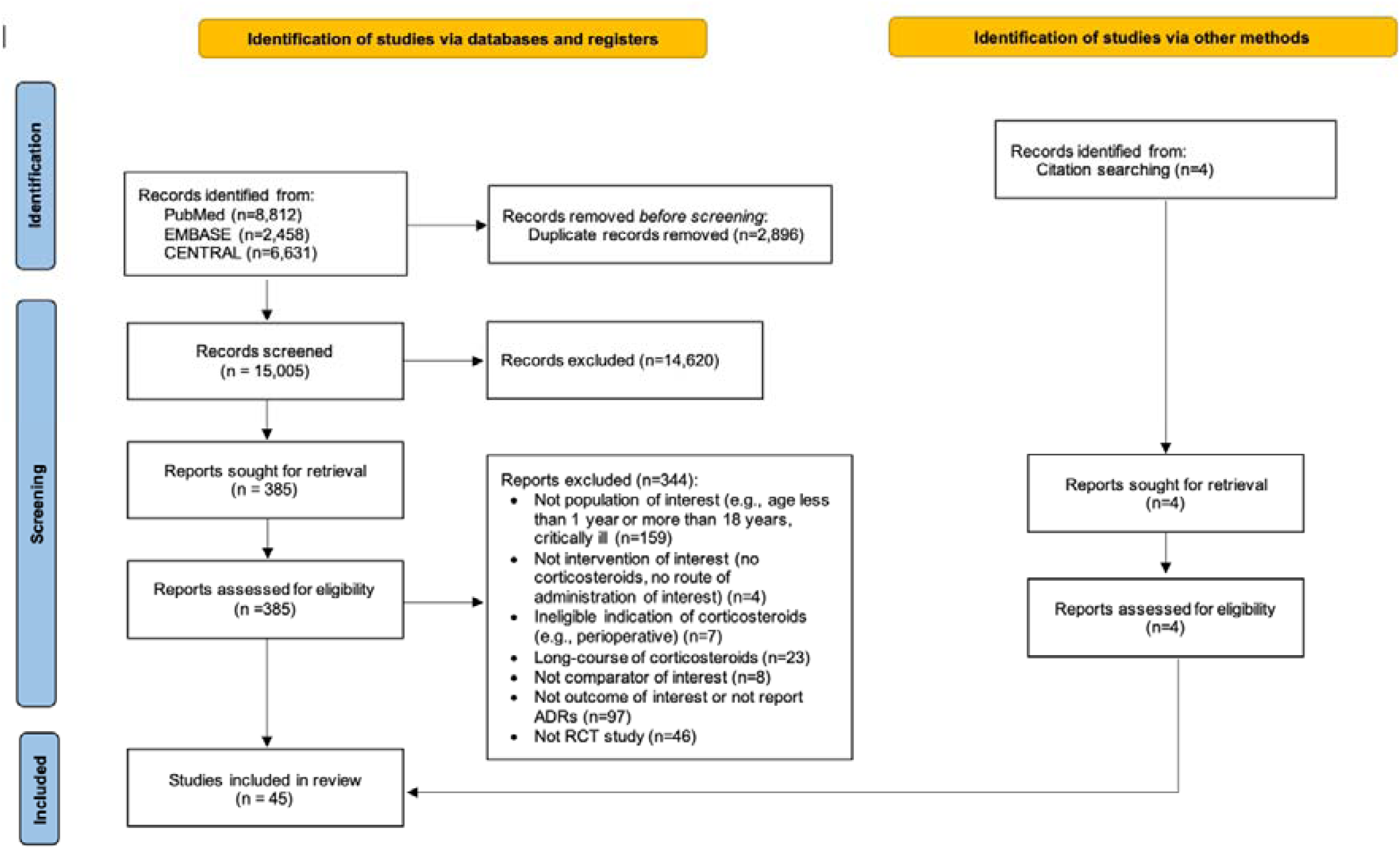
PRISMA flow diagram of selected studies

### Trial and Participants Characteristics

Forty-five studies included 6,470 children with a mean age of 5.57 years (standard deviation [SD] 3.62) and 58% male participants. The majority were conducted in Europe and North America (69%), followed by Asia (27%) and Australia and New Zealand (4%). The most common indications included were encephalitis or meningitis in six (13%) (39, 43, 44, 47, 58, 61), croup in five (11%) (29, 42, 45, 48, 65), and pneumonia in five (11%) trials (49, 51, 63, 66, 71).

Seventeen trials including 2,421 participants investigated dexamethasone (28, 29, 37–39, 42–45, 47, 48, 54, 58, 61, 62, 65, 66), 13 (n=964) methylprednisolone (30, 36, 49, 51–53, 57, 63, 64, 68–71), 12 (n=2313) prednisolone (27, 31–35, 41, 46, 50, 59, 60, 67), two studies (n=727) examined prednisone (40, 55) and one study (n=45) hydrocortisone, dexamethasone or betamethasone (56). The median duration of corticosteroid treatment was 3 days (interquartile range [IQR] 1 to 5). In 27 trials (60%) corticosteroid administration was intravenous or intramuscular and 18 trials (40%) offered the medication via the oral route. While 32 trials (71%) were conducted in an inpatient setting, eleven (24%) outpatient only, and two (4%) included a mixed population. Table 1 presents the characteristics of all included studies.

**Table 1.**
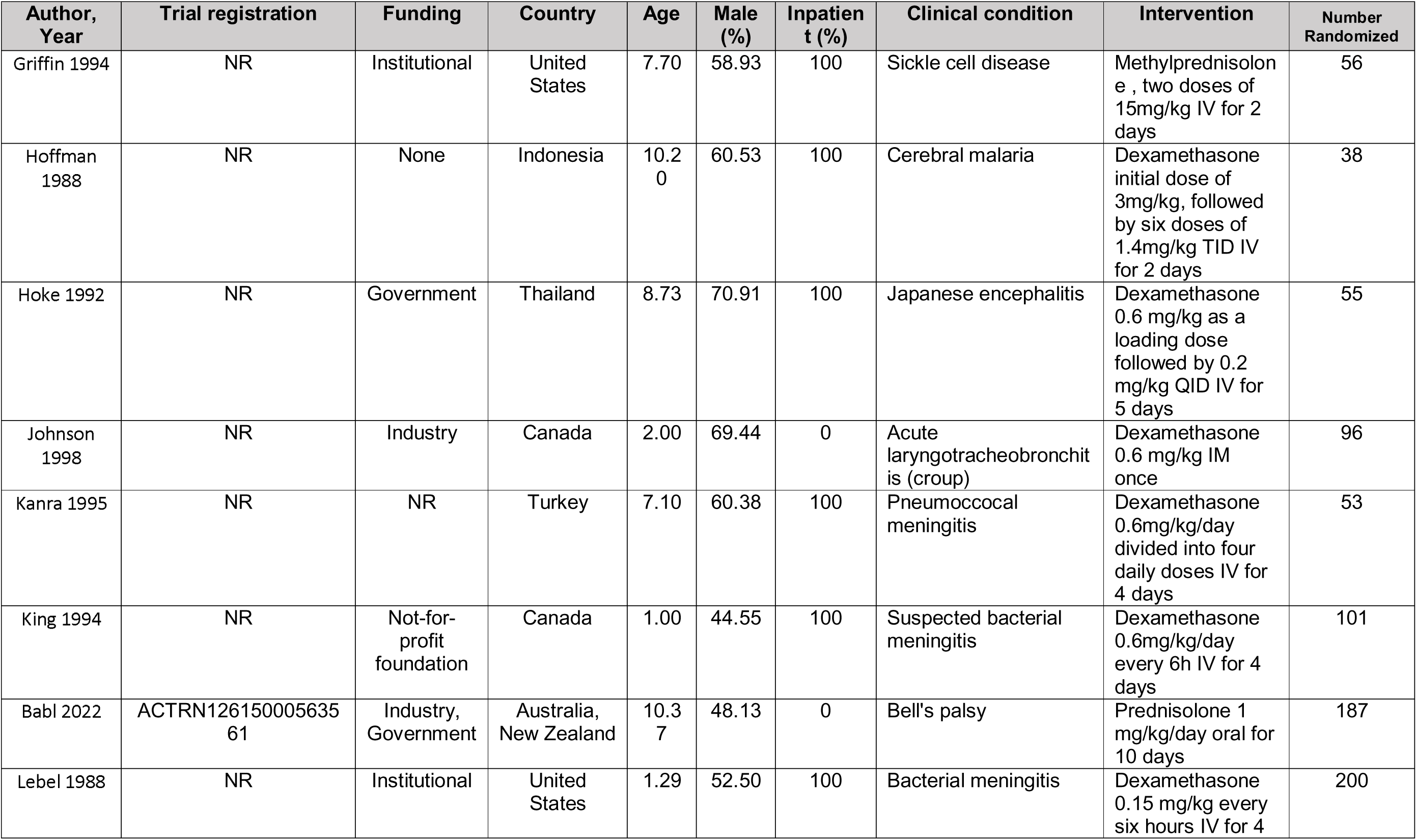
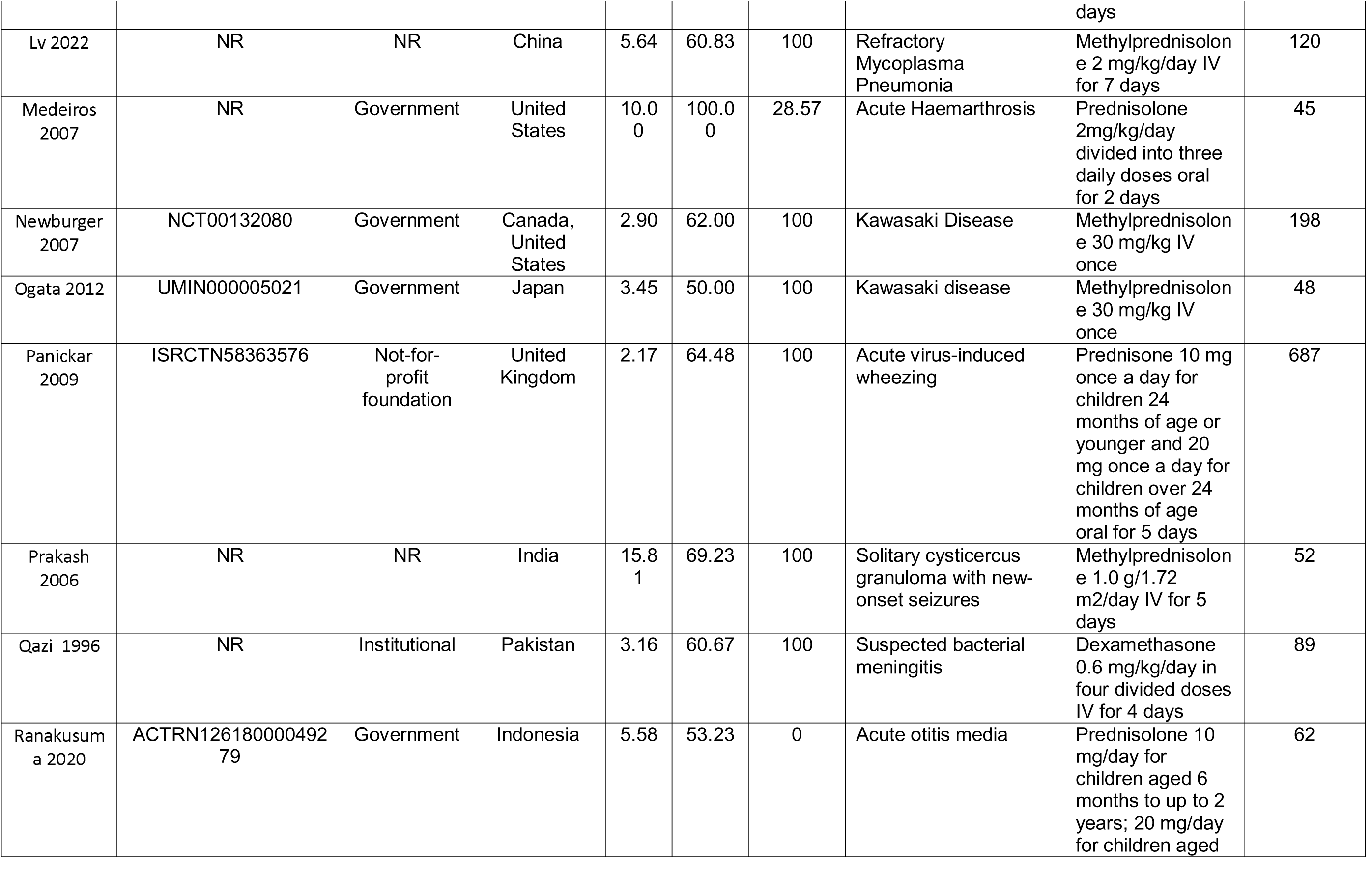
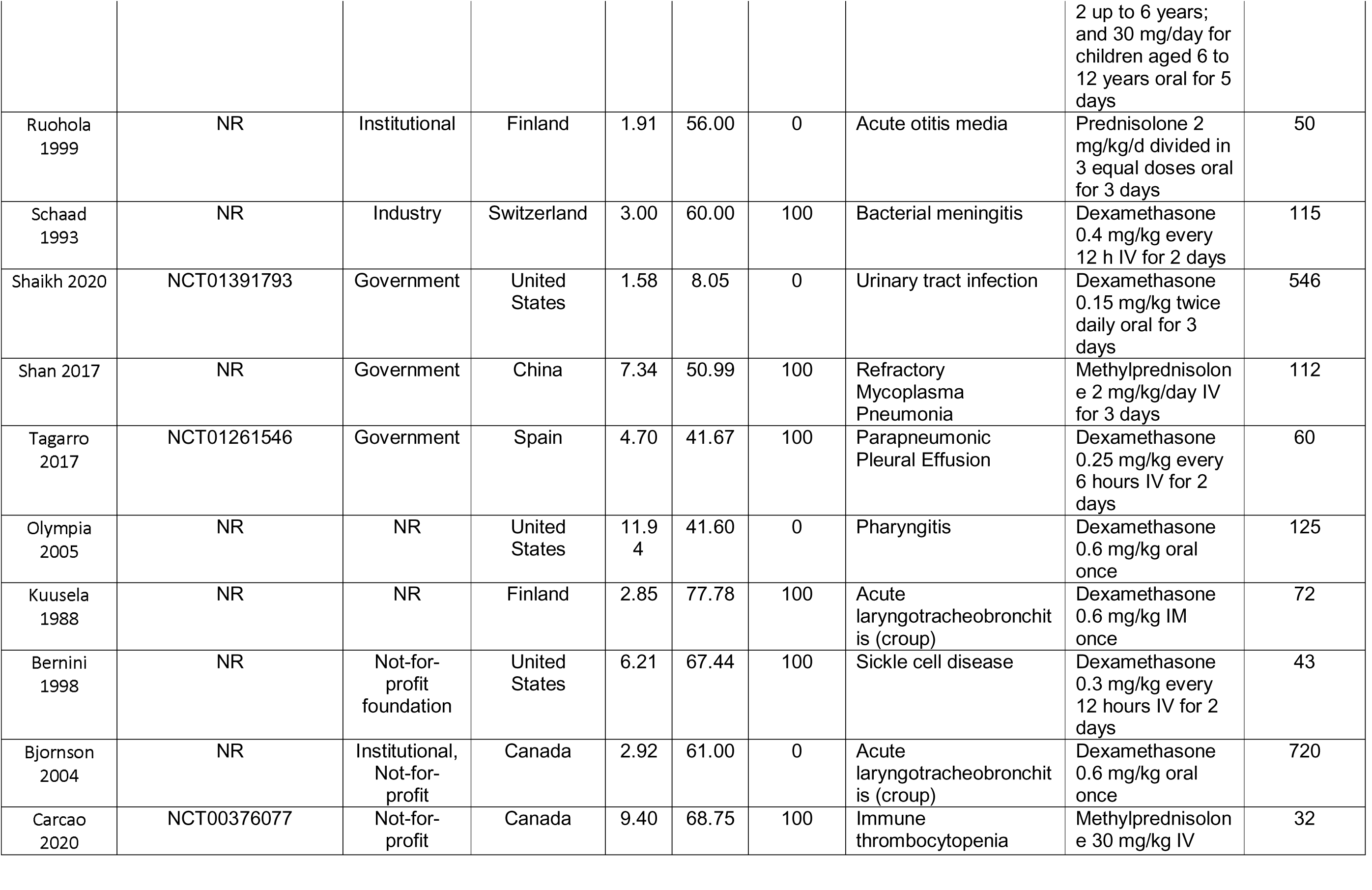
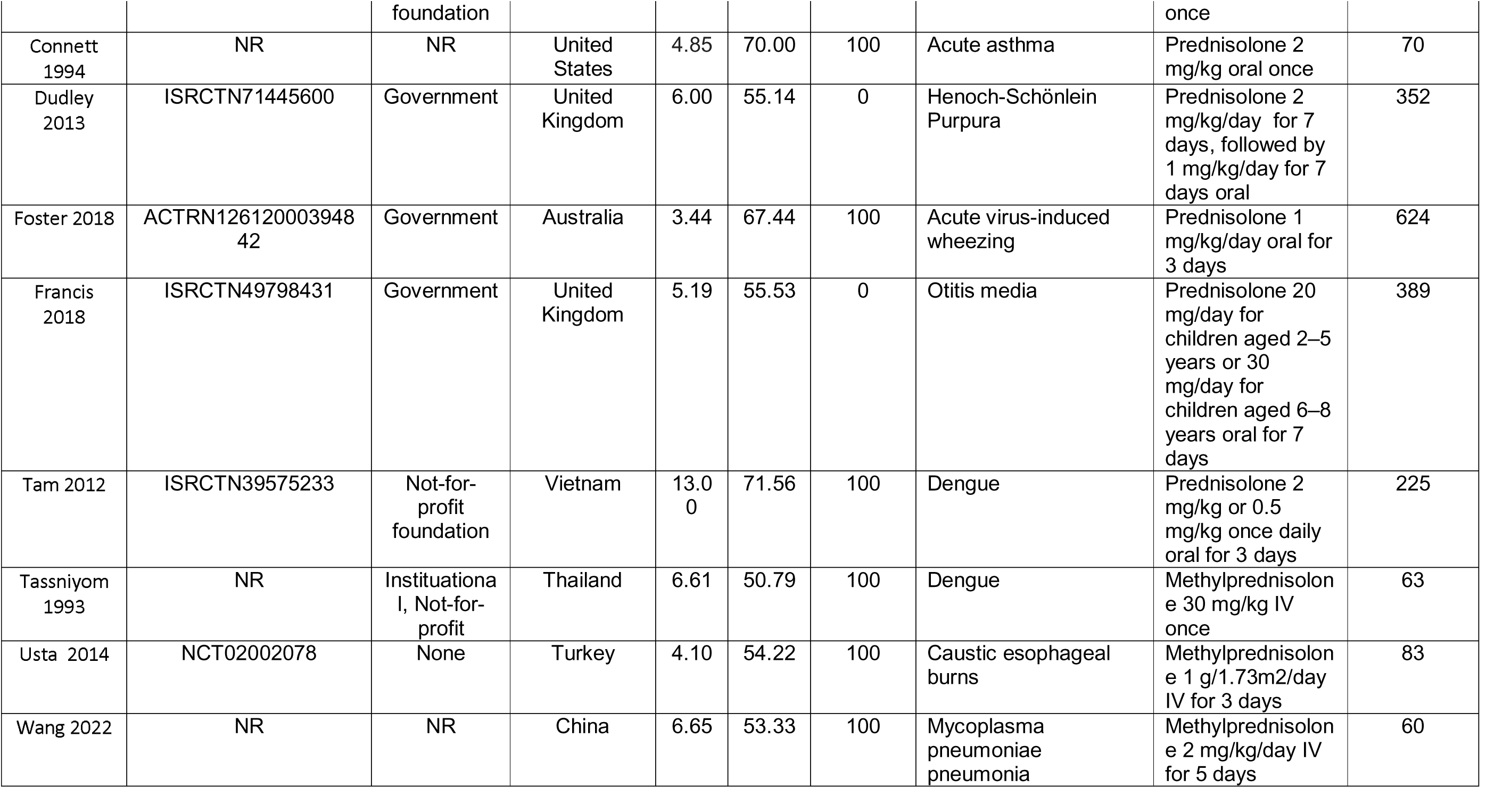
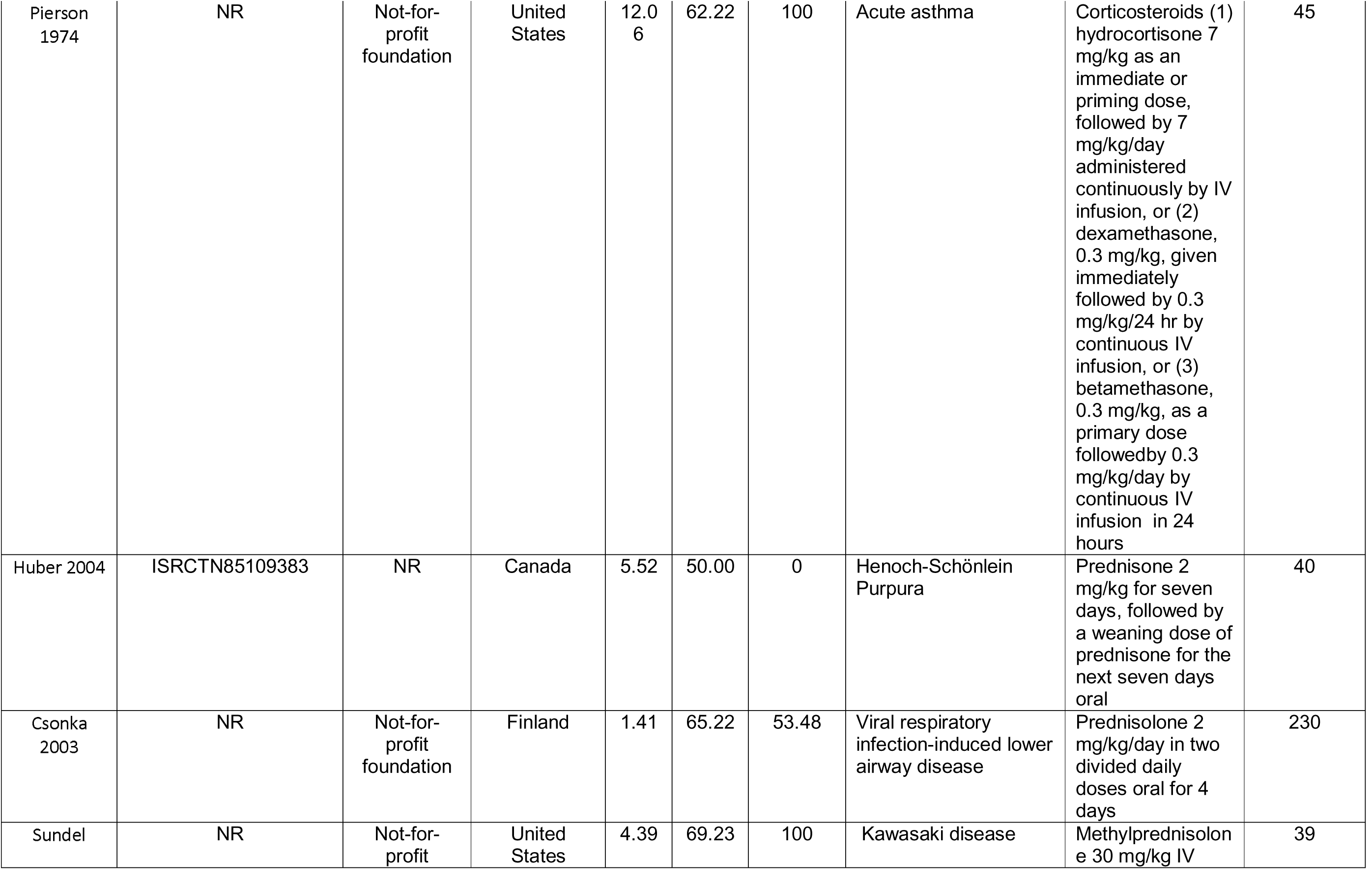
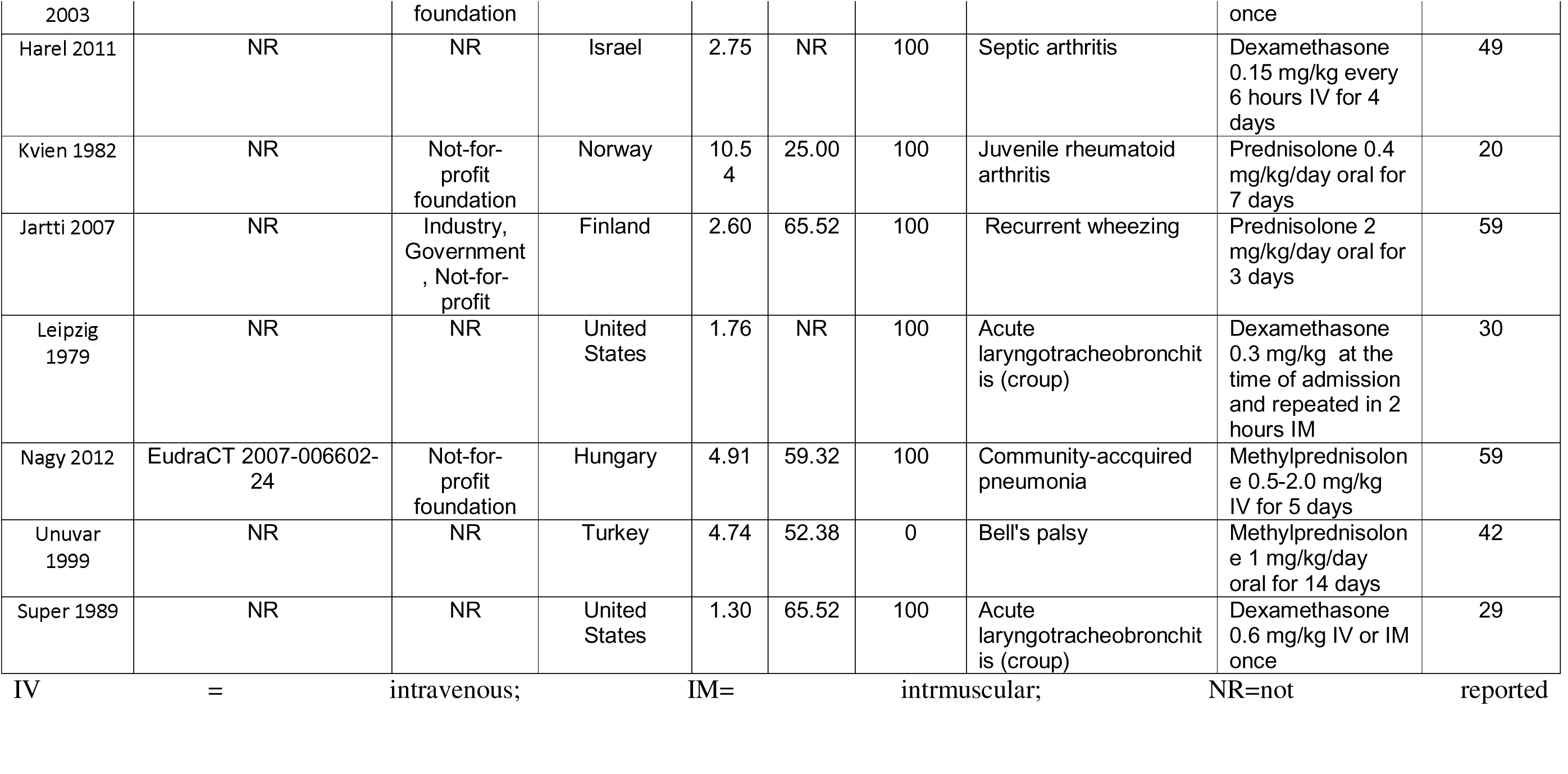
Study characteristics.

### Risk of Bias assessment

Of the 45 eligible trials, fifteen (33%) had at least one domain at probably high or high risk of bias. Failure to blind was the main reason for rating down risk of bias: eleven trials (24%) did not blind participants, eight (18%) did not blind healthcare providers, and nine (20%) did not blind data collectors. Table 2 presents the detailed risk of bias assessment.

**Table 2.**
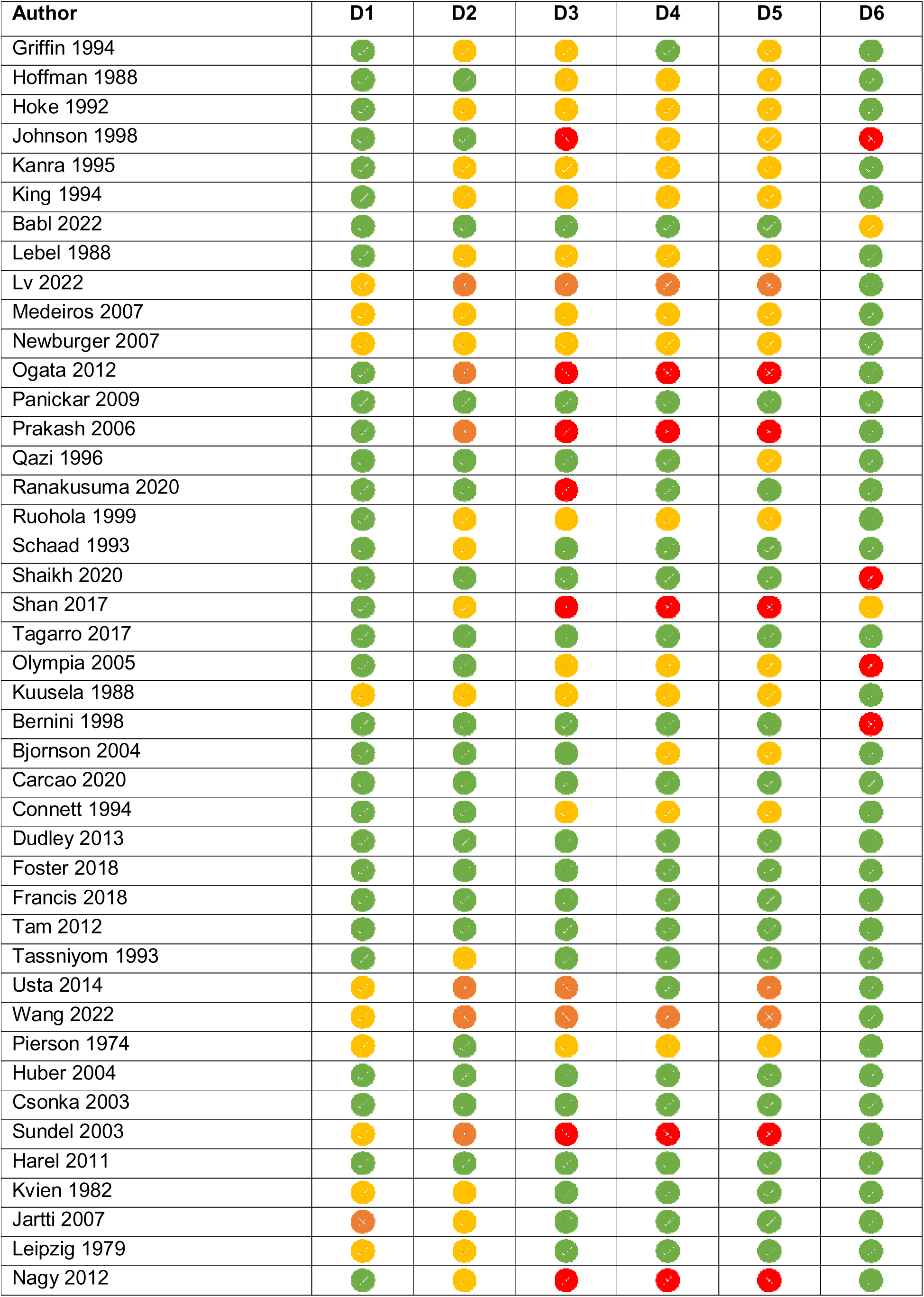
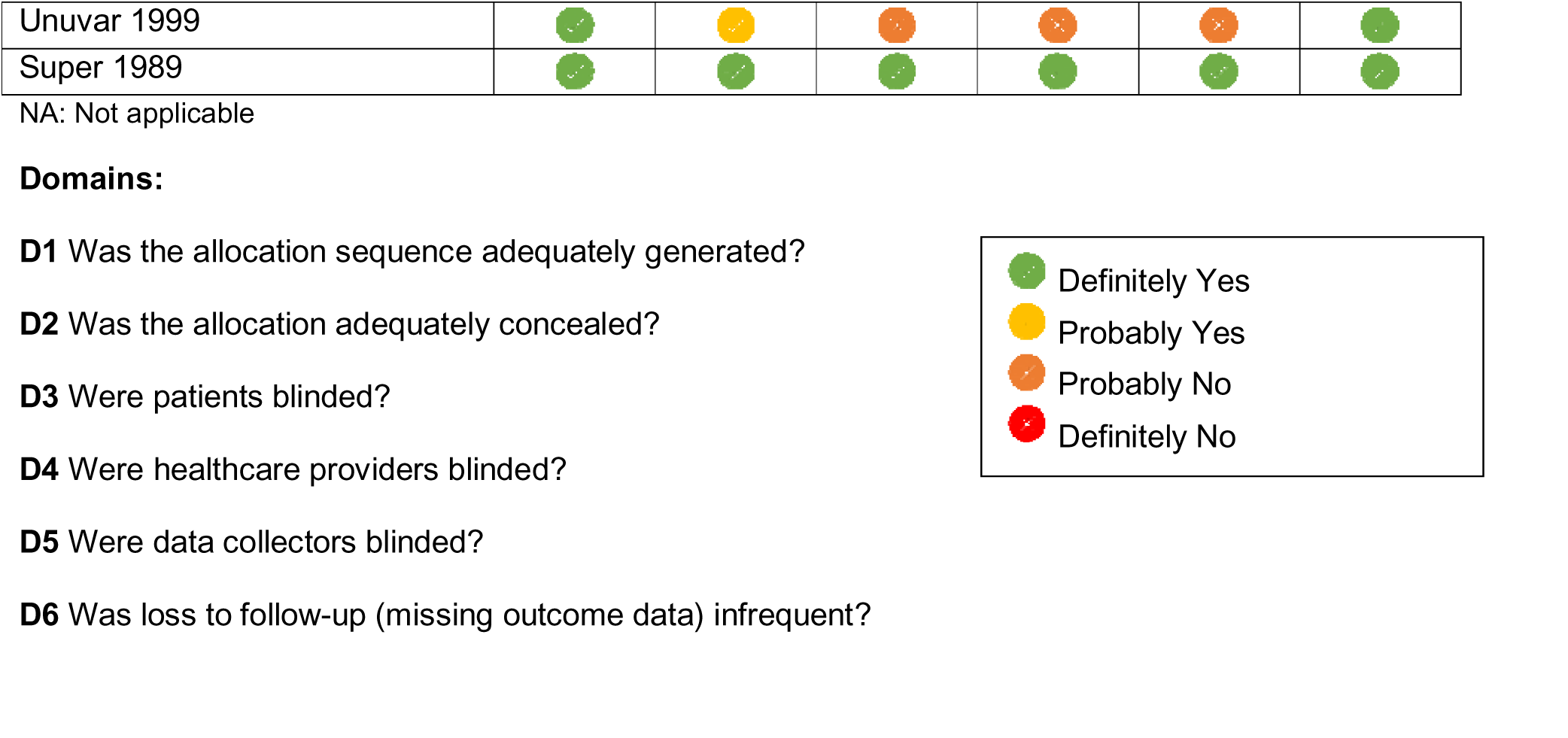
Risk of Bias Assessment.

### Adverse Events

Of the 36 adverse events that we were able to meta-analyze, 31 proved to show no compelling difference between corticosteroids and the control group. Table 3-7 presents SoF tables with all the outcomes and *Supplementary Material 4* AEs reported in a single trial. Below, we present outcomes that we deemed the most important short-term harms.

**Table 3:** Summary of Findings Table (SoF) 1 - Serious Adverse Events and Adverse Events Leading to Discontinuation. Population: Children and Adolescents 1-18 years old Intervention: Short-course (<15 days) systemic corticosteroids Comparator: Placebo or standard of care

| Outcome | Study results and measurements | Certainty of the evidence<br>(Quality of evidence) | Summary |
| --- | --- | --- | --- |
| Serious Adverse Events | Risk difference: -0.001<br>[-0.009, 0.007]<br>Based on data from 3418 participants in 19 studies | **Moderate**<sup>1</sup> | Corticosteroids probably have little to no impact on the risk of serious adverse events |
| Adverse events leading to discontinuation | Risk difference: 0.004<br>[-0.003, 0.011]<br>Based on data from 2892 participants in 16 studies | **Moderate**<sup>1</sup> | Corticosteroids probably increase the risk of adverse events leading to discontinuation slightly |

1. Imprecision: serious - confidence intervals crossing null effect (RD=0).

### Serious Adverse events (SAEs)

Nineteen studies, including 3,418 participants, reported data on SAEs. Corticosteroids (RD 1 fewer per 1,000 [95% CI 9 fewer to 7 more] moderate certainty) probably cause very few or no SAE. (Table 3)

### AE leading to discontinuation

Sixteen studies, including 2,892 participants, reporting on AE leading to discontinuation shows that corticosteroids (RD 4 more per 1,000 [95%CI 3 fewer to 11 more]; moderate certainty) probably result in a small number of AEs leading to discontinuation. (Table 3)

### Hyperglycemia

Twelve studies, including 792 participants corticosteroids (RD 38 more per 1,000 [95% CI 11 to 64 more]; moderate certainty) probably increase the risk of hyperglycemia. (Table 5)

**Table 4:** Summary of Findings Table (SoF) 2 – Gastrointestinal Adverse Events. Population: Children and Adolescents 1-18 years-old Intervention: Short-course (<15 days) systemic corticosteroids Comparator: Placebo or standard of care

| **Outcome** | **Study results and measurements** | **Certainty of the evidence**<br>(Quality of evidence) | **Summary** |
| --- | --- | --- | --- |
| Abdominal Pain | Risk difference<br>0.005 [-0.003, 0.013]<br>Based on data from<br>2410 participants in 15<br>studies | **Moderate**<sup>1</sup> | Corticosteroids probably increase the risk of abdominal pain slightly |
| Diarrhea | Risk difference:<br>-0.001 [-0.012, 0.009]<br>Based on data from<br>3280 participants in 11<br>studies | **Moderate**<sup>1</sup> | Corticosteroids probably have little to no impact on the risk of diarrhea |
| Gastritis | Risk difference:<br>-0.004 [-0.025, 0.016]<br>Based on data from<br>488 participants in 10<br>studies | **Low**<sup>2</sup> | Corticosteroids may decrease the risk of gastritis slightly |
| Gastrointestinal Bleeding | Risk difference:<br>0.013 [0.003, 0.023]<br>Based on data from<br>2555 participants in 14<br>studies | **Moderate**<sup>3</sup> | Corticosteroids probably increase the risk of gastrointestinal bleeding |
| Hemoccult-positive stool | Risk difference:<br>0.022 [-0.010, 0.054]<br>Based on data from<br>774 participants in 3<br>studies | **Very low**<sup>2,4</sup> | We are uncertain whether corticosteroids increase or decreases the risk of hemoccult-positive stool. |
| Intussusception | Risk difference: -0.008<br>[-0.017, 0.001]<br>Based on data from<br>771 participants in 10<br>studies | **Low**<sup>2</sup> | Corticosteroids may decrease the risk of intussusception slightly |
| Nausea | Risk difference: -0.008<br>[-0.022, 0.005]<br>Based on data from<br>1486 participants in 14<br>studies | **Moderate**<sup>1</sup> | Corticosteroids probably decreases risk of nausea slightly |
| Vomiting | Risk difference: -0.004<br>[-0.014, 0.006]<br>Based on data from<br>2857 participants in 19<br>studies | **Low**<sup>1,6</sup> | Corticosteroids may decrease the risk of vomiting slightly |

1. Imprecision: serious - confidence intervals crossing null effect (RD=0). 2. Imprecision: very serious - confidence intervals crossing null effect (RD=0) and a small number of total events (<30). 3. Inconsistency: serious – p -value for inconsistency = 0.01; $I^2 = 46.1\%$ 4. Indirectness: serious - surrogate outcome. 5. Risk of Bias: serious.

**Table 5:** Summary of Findings Table (SoF) 3 – Metabolism, nutrition, renal and urinary Adverse Events. Population: Children and Adolescents 1-18 years-old Intervention: Short-course (<15 days) systemic corticosteroids Comparator: Placebo or standard of care

| Outcome | Study results and measurements | Certainty of the evidence<br>(Quality of evidence) | Summary |
| --- | --- | --- | --- |
| Hyperglycemia | Risk difference: 0.038 [0.011, 0.064]<br>Based on data from 792 participants in 12 studies | **Moderate**<sup>1</sup> | Corticosteroids probably increase the risk of hyperglycemia |
| Decreased Appetite | Risk difference: -0.010 [-0.022, 0.001]<br>Based on data from 977 participants in 11 studies | **Low**<sup>2</sup> | Corticosteroids may decrease the risk of decreased appetite slightly |
| Increased Appetite | Risk difference: 0.015 [-0.006, 0.037]<br>Based on data from 1024 participants in 12 studies | **Moderate**<sup>3</sup> | Corticosteroids probably increases the risk of increased appetite slightly |
| Glycosuria | Risk difference: 0.036 [0.016, 0.055]<br>Based on data from 778 participants in 10 studies | **Low**<sup>1,3</sup> | Corticosteroids may increase the risk of glycosuria |
| Polyuria | Risk difference: -0.002 [-0.019, 0.014]<br>Based on data from 980 participants in 11 studies | **Low**<sup>2</sup> | Corticosteroids may have little to no impact on the risk of polyuria |

1. Inconsistency: serious. – Hyperglycemia ( $I^2=36.3\%$ ); Glycosuria ( $I^2=87.3\%$ , $p<0.01$ )
2. Imprecision: very serious. confidence intervals crossing null effect (RD=0) and a small number of total events (<30).
3. Imprecision: serious - confidence intervals crossing null effect (RD=0).

### Sleep Problems

Twelve studies including 1525 children reported on sleep problems. Corticosteroids (RD 15 more per 1,000 [95% 1 to 28 more]; moderate certainty) probably increase the risk of sleep problems. (Table 6)

**Table 6:**
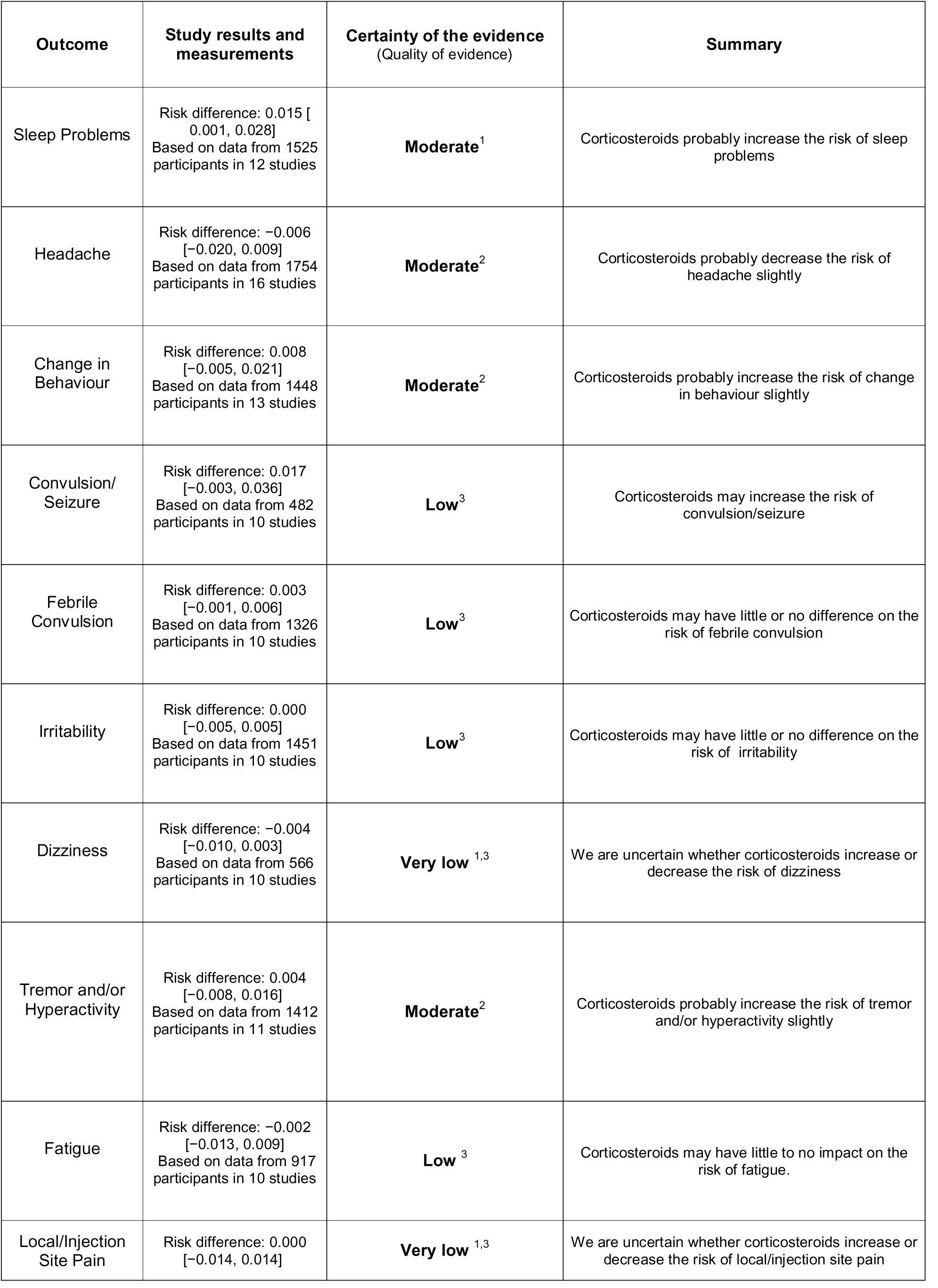
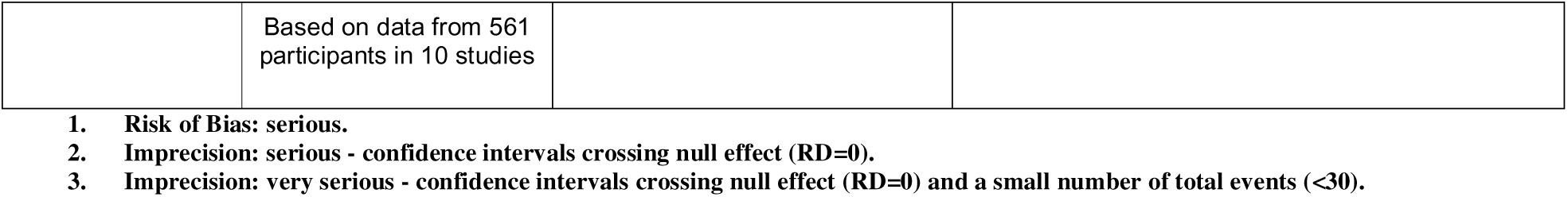
Summary of Findings Table (SoF) 4 – Central Nervous System, Psychiatric and general disorders Adverse Events. Population: Children and Adolescents 1-18 years-old Intervention: Short-course (<15 days) systemic corticosteroids Comparator: Placebo or standard of care

### Change in behavior

Thirteen studies, including 1,448 children, showed that corticosteroids (RD 8 more [95% CI 5 fewer to 21 more]; moderate certainty) probably increase the risk of a change in behavior compared to the control group. (Table 6)

### Gastrointestinal (GI) bleeding

Fourteen studies, including 2,555 participants, reported on GI bleeding. Corticosteroids (RD 13 more per 1,000 [95% 3 to 23 more]; moderate certainty) probably increase the risk of gastrointestinal bleeding. (Table 4).

### Subgroup and Sensitivity analyses

For GI Bleeding, we found a statistically significant (IV/IM RD 0.035 [95% 9 to 61 more] vs. oral RD 1 fewer per 1,000 [95% 9 fewer to 8 more]; p=0.01) subgroup effect based on the route of administration. However, this effect has low credibility using the ICEMAN criteria (*Supplementary material 5 and 7.1*). Reasons for low credibility were analysis based on comparisons between trials and the use of a fixed-effects model. We did not observe any other significant subgroup effect based on dose, duration or clinical condition (*Supplementary material 7.2 to 7.7*). Our sensitivity analyses using Peto’s OR showed similar results to those of our primary analyses. (*Supplementary material 6*).

## Discussion

### Main findings

Our results demonstrated moderate certainty evidence that using systemic corticosteroids for a short duration (<15 days) in a pediatric population likely results in a small increase in the risk of non-serious hyperglycemia, sleep problems, change in behaviour and GI bleeding and adverse events that lead to treatment discontinuation. Our results also demonstrated that corticosteroids likely have little to no impact on the risk of SAE.

### Relation to other findings

A previous systematic review investigated the AEs of short-course oral corticosteroids in children and showed similar results in terms of incidence rates. Vomiting, behavioral changes and sleep disturbance were among the most frequent AEs.(7) This review only included oral interventions, excluded studies at high risk of bias, failed to assess the certainty of evidence, included observational studies and did not perform meta-analyses. Our review confirms that those AEs were among the most frequent AEs, but — even more important to patients and clinicians — it showed that corticosteroids likely increase the risk of change in behavior and sleep problems. This prior review unfoundedly highlighted an increase in infection rates whereas we found that evidence regarding infections was only low to very low certainty evidence and failed to support an increase in the risk of any infection (Table 7). Other systematic reviews on corticosteroid-related AEs (6, 72), also reported that corticosteroids probably have little to no effect on the risk of secondary infections.

**Table 7.** Summary of Findings Table (SoF) 5 – Infections, musculoskeletal, blood, cardiac and skin and subcutaneous tissue disorders. Population: Children and Adolescents 1-18 years-old Intervention: Short-course (<15 days) systemic corticosteroids Comparator: Placebo or standard of care

| **Outcome** | **Study results and measurements** | **Certainty of the evidence**<br>(Quality of evidence) | **Summary** |
| --- | --- | --- | --- |
| Candidiasis | Risk difference: 0.008<br>[−0.007, 0.023]<br>Based on data from 503 participants in 10 studies | **Very low**<sup>1,2</sup> | We are uncertain whether corticosteroids increase or decrease the risk of candidiasis |
| Pneumonia | Risk difference: 0.004 [−0.012, 0.019]<br>Based on data from 1481 participants in 13 studies | **Low**<sup>2</sup> | Corticosteroids may increase the risk of pneumonia slightly |
| Secondary (opportunistic) infection | Risk difference: 0.008 [−0.007, 0.024]<br>Based on data from 929 participants in 12 studies | **Low**<sup>2</sup> | Corticosteroids may increase the risk of secondary (opportunistic) infection slightly |
| Otitis Media | Risk difference: −0.007 [−0.016, 0.003]<br>Based on data from 1164 participants in 10 studies | **Low**<sup>2</sup> | Corticosteroids may decrease the risk of otitis media slightly |
| Urinary Tract Infection | Risk difference: 0.002 [−0.011, 0.015]<br>Based on data from 1186 participants in 11 studies | **Low**<sup>2</sup> | Corticosteroids may have little to no difference on the risk of urinary tract infection |
| Musculoskeletal Pain | Risk difference: −0.004 [−0.009, 0.001]<br>Based on data from 1080 participants in 10 studies | **Very low**<sup>1,2</sup> | We are uncertain whether corticosteroids increase or decrease the risk of musculoskeletal pain |
| Myalgia | Risk difference: 0.003<br>[−0.003, 0.009]<br>Based on data from<br>693 participants in 10<br>studies | **Very low**<sup>1,2</sup> | We are uncertain whether corticosteroids increase or decrease the risk of myalgia |
| Anemia | Risk difference: −0.012<br>[−0.035, 0.012]<br>Based on data from<br>701 participants in 11<br>studies | **Low**<sup>2</sup> | Corticosteroids may decrease the risk of anemia |
| Hypertension | Risk difference: 0.001<br>[−0.006, 0.009]<br>Based on data from<br>1270 participants in 16<br>studies | **Low**<sup>2</sup> | Corticosteroids may have little to no difference on the risk of hypertension. |
| Congestive Heart Failure | Risk difference: −0.006<br>[−0.015, 0.003]<br>Based on data from<br>618 participants in 10<br>studies | **Very low**<sup>1,2</sup> | We are uncertain whether corticosteroids increase or decrease the risk of congestive heart failure |
| Rash and Urticaria | Risk difference: −0.003<br>[−0.009, 0.003]<br>Based on data from<br>3098 participants in 20<br>studies | **Moderate**<sup>3</sup> | Corticosteroids probably has little or no difference on rash and urticaria |

1. Risk of Bias: serious.
2. Imprecision: very serious - confidence intervals crossing null effect (RD=0) and a small number of total events (<30).
3. Imprecision: serious - confidence intervals crossing null effect (RD=0).

Findings from our study align with results from other systematic reviews and meta-analyses in adult and pediatric populations. Previous reviews , one investigating the AEs of corticosteroids in children with acute respiratory conditions and other the metabolic AEs of corticosteroids in adults, documented the increased risk of change in behavior and hyperglycemia.(6, 73)

Our results from GI bleeding, on the other hand, differ from prior reviews. Fernandes and colleagues (6), conducted a systematic review on the safety of systemic corticosteroids across acute respiratory conditions and did not report an increase in the risk of GI bleeding. This might be explained by the inclusion of observational studies leading to biased estimates and restriction to respiratory clinical syndromes in their eligibility criteria. On the other hand, an observational study, including a cohort of 23 million individuals in Taiwan, also found an increase in the incidence rate ratio (2.02 [95% 1.55-2.64]) of GI bleeding with the use of short-term corticosteroids.(74)

Even though we found no significant subgroup effect based on clinical condition, a significant proportion of GI bleeding events happened in trials involving patients with central nervous system infections (mostly meningitis). The role of adjunctive corticosteroids in pediatric bacterial meningitis, however, remains controversial. The latest Cochrane review (75) suggests no benefit of corticosteroids on the risk of hearing loss and mortality in pediatric non-*Haemophilus influenzae* meningitis. This raises concerns about the risk-benefit balance of routine corticosteroid use in this population. Current guidelines (76, 77) advocate for systemic corticosteroids, yet they do not sufficiently address potential adverse effects, particularly the increased risk of gastrointestinal complications. Given the lack of benefit and the underrecognized harms, a more cautious, individualized approach may be warranted. Research to better evaluate the impact of corticosteroids on GI bleeding, not only on CNS infections but across different conditions, is necessary. *Supplementary Material 8* presents a summary of previous systematic reviews.

### Strengths and Limitations

Strengths of our study include a comprehensive search without any restriction on language or type of clinical condition and the use of the GRADE approach for assessing the certainty of evidence.(23)

The limitations of our study relate to limitations in AE reporting in RCTs.(78) AE reporting was not a primary focus of the included studies, and authors seldom report either the definition of AEs or the methods used to capture AEs (*see Supplementary Material 3*). It is possible that these problems resulted in an underestimation of adverse events.

### Implications for clinical practice and research

The importance of our work is that by looking across clinical conditions we were able to achieve sample sizes that were sufficient to result in sufficiently narrow confidence intervals to provide moderate certainty evidence of steroid impact on AE. Our research shows that corticosteroids — even when prescribed for a short period of time — are not harmless but rather increase the risk of AEs, including GI bleeding, sleep disturbance and hyperglycemia. The magnitude of the effect is, however, small – less than 4% for hyperglycemia and less than 1.5% for the other outcomes – and there is no increase in SAEs. These results can inform both clinical practice guidelines and clinical practice. The impact on decision-making will depend on the importance patients and families place on the small increase in relatively minor adverse events.

The methodology used in this systematic review and meta-analysis — pooling data on AEs reported in RCTs regardless of clinical condition—should be widely used for medications, such as corticosteroids, with several clinical indications. Future research will not only focus on similar reviews for other classes of medications but also on developing methodological guidance for those interested in conducting their own reviews.

## Data Availability

All data produced in the present study are available upon reasonable request to the authors

## Declarations

### Authors’ contributions

Conceptualization: JPL, SRC, DC and GHG; Methodology: JPL, SRC, WT; Resource: JPL, WT, RC; Investigation: JPL, SRC, WT, CZ, XC, JMS, MRC, HK; Data curation: JPL; Writing-original draft: JPL; Writing-review and editing: all authors; Visualization: JPL; Project administration: JPL; Supervision: DC, GHG, ME

## Abbreviations

AE: adverse event
AIDS: acquired immunodeficiency syndrome
CI: confidence intervals
GRADE: The Grading of Recommendations Assessment, Development and Evaluation
GI: gastrointestinal bleeding
HIV: human immunodeficiency virus
MeSH: Medical Subject Headings
PRISMA: Preferred Reporting Items for Systematic reviews and Meta-Analyses
RCT: randomized controlled trial
RD: risk difference
SAE: serious adverse events

## Appendix 1. Search strategy

**Ovid MEDLINE(R) and Epub Ahead of Print, In-Process, In-Data-Review & Other Non-Indexed Citations, Daily and Versions 1946 to January 22, 2024**

| # | Query | January 22, 2024 |
| --- | --- | --- |
| 1 | exp Glucocorticoids/ | 209077 |
| 2 | (steroid* or corticosteroid* or glucocorticoid* or prednisolone* or dexamethasone* or hydrocortisone* or methylprednisolone*).mp. | 711896 |
| 3 | 1 or 2 | 747413 |
| 4 | exp Child/ | 2183979 |
| 5 | (child* or stepchild* or step-child* or kid or kids or girl or girls or boy or boys or teen* or youth* or youngster* or adolescent* or adolescence or preschool* or pre-school* or kindergarten* or school* or juvenile* or minors or p?ediatric*).mp. | 4380835 |
| 6 | 4 or 5 | 4380835 |
| 7 | exp "Drug-Related Side Effects and Adverse Reactions"/ | 134494 |
| 8 | (ae or to or po or co).fs. | 4538295 |
| 9 | (safe or safety).ti,ab. | 1070134 |
| 10 | side effect\$.ti,ab. | 306521 |
| 11 | ((adverse or undesirable or harms\$ or serious or toxic) adj3 (effect\$ or reaction\$ or event\$ or outcome\$)).ti,ab. | 702072 |
| 12 | exp Product Surveillance, Postmarketing/ | 18535 |
| 13 | adverse drug reaction reporting systems/ | 8986 |
| 14 | exp Poisoning/ | 170444 |
| 15 | exp Substance-Related Disorders/ | 314497 |
| 16 | Abnormalities, Drug-Induced/ | 14746 |
| 17 | Drug Monitoring/ | 23752 |
| 18 | (toxicity or complication\$ or noxious or tolerability).ti,ab. | 1674179 |
| 19 | or/7-18 | 6825213 |
| 20 | (ae or de or co or ci).fs. | 6986870 |
| 21 | safe*.ti,ab. | 1184052 |
| 22 | (safe* or adverse or side effect* or complication* or tolerated or tolerance or harm or toxicity).ti,ab. | 3509560 |
| 23 | risk.ti. | 604713 |
| 24 | tolerability.ti,ab. | 60648 |
| 25 | to.fs. | 487577 |
| 26 | Toxicology/ | 27283 |
| 27 | drug induced.ti,ab. | 45092 |
| 28 | negative effects.ti,ab. | 40034 |
| 29 | or/20-28 | 9703479 |
| 30 | 19 or 29 | 9997859 |
| 31 | randomized controlled trial.pt. | 607255 |
| 32 | controlled clinical trial.pt. | 95538 |
| 33 | randomi?ed.ab. | 754370 |
| 34 | placebo.ab. | 244999 |
| 35 | drug therapy.fs. | 2659840 |
| 36 | randomly.ab. | 425508 |
| 37 | trial.ab. | 681656 |
| 38 | groups.ab. | 2625460 |
| 39 | or/31-38 | 5880814 |
| 40 | exp animals/ not humans.sh. | 5189493 |
| 41 | 39 not 40 | 5142172 |
| 42 | 3 and 6 and 30 and 41 | 38292 |
| 43 | 3 and 6 and 41 | 63493 |
| 44 | (steroid* or corticosteroid* or glucocorticoid* or prednisolone* or dexamethasone* or hydrocortisone* or methylprednisolone*).ti. | 174922 |
| 45 | 44 or 1 | 308990 |
| 46 | 6 and 30 and 41 and 44 | 8812 |

Embase 1974 to 2024 January 22

| # | Query | January 22, 2024 |
| --- | --- | --- |
| 1 | exp Glucocorticoids/ | 877108 |
| 2 | (steroid* or corticosteroid* or glucocorticoid* or prednisolone* or dexamethasone* or hydrocortisone* or methylprednisolone*).mp. [mp=title, abstract, heading word, drug trade name, original title, device manufacturer, drug manufacturer, device trade name, keyword heading word, floating subheading word, candidate term word] | 1360218 |
| 3 | 1 or 2 | 1515797 |
| 4 | (steroid* or corticosteroid* or glucocorticoid* or prednisolone* or dexamethasone* or hydrocortisone* or methylprednisolone*).m_titl. | 201690 |
| 5 | exp Child/ | 3153221 |
| 6 | (child* or stepchild* or step-child* or kid or kids or girl or girls or boy or boys or teen* or youth* or youngster* or adolescent* or adolescence or preschool* or pre-school* or kindergarten* or school* or juvenile* or minors or p?ediatric*).mp. [mp=title, abstract, heading word, drug trade name, original title, device manufacturer, drug manufacturer, device trade name, keyword heading word, floating subheading word, candidate term word] | 4638464 |
| 7 | 5 or 6 | 5158495 |
| 8 | exp "Drug-Related Side Effects and Adverse Reactions"/ | 651171 |
| 9 | (ae or to or po or co).fx. | 4023407 |
| 10 | (safe or safety).ti,ab. | 1614518 |
| 11 | side effect\$.ti,ab. | 453716 |
| 12 | ((adverse or undesirable or harms\$ or serious or toxic) adj3 (effect\$ or reaction\$ or event\$ or outcome\$)).ti,ab. | 1073337 |
| 13 | exp Product Surveillance, Postmarketing/ | 39629 |
| 14 | adverse drug reaction reporting systems/ | 6031 |
| 15 | exp Poisoning/ | 448574 |
| 16 | exp Substance-Related Disorders/ | 281792 |
| 17 | Abnormalities, Drug-Induced/ | 111 |
| 18 | Drug Monitoring/ | 60705 |
| 19 | (toxicity or complication\$ or noxious or tolerability).ti,ab. | 2440654 |
| 20 | safe*.ti,ab. | 1778233 |
| 21 | (safe* or adverse or side effect* or complication* or tolerated | 5023883 |
|  | or tolerance or harm or toxicity).ti,ab. |  |
| 22 | risk.ti. | 840842 |
| 23 | (ae or de or co or ci).fx. | 3081786 |
| 24 | tolerability.ti,ab. | 113578 |
| 25 | to.fx. | 607438 |
| 26 | Toxicology/ | 41478 |
| 27 | drug induced.ti,ab. | 63060 |
| 28 | negative effects.ti,ab. | 46797 |
| 29 | or/8-28 | 9032776 |
| 30 | randomized controlled trial.pt. | 0 |
| 31 | controlled clinical trial.pt. | 0 |
| 32 | randomi?ed.ab. | 1095263 |
| 33 | placebo.ab. | 362291 |
| 34 | drug therapy.fx. | 4533827 |
| 35 | randomly.ab. | 566168 |
| 36 | trial.ab. | 1004254 |
| 37 | groups.ab. | 3681105 |
| 38 | or/30-37 | 8828517 |
| 39 | exp human/ | 26101879 |
| 40 | exp animals/ | 31304609 |
| 41 | 40 not 39 | 5202730 |
| 42 | 38 not 41 | 7859558 |
| 43 | 4 and 7 and 29 and 42 | 9746 |
| 44 | limit 43 to "remove medline records" | 2458 |

Cochrane Library CDSR

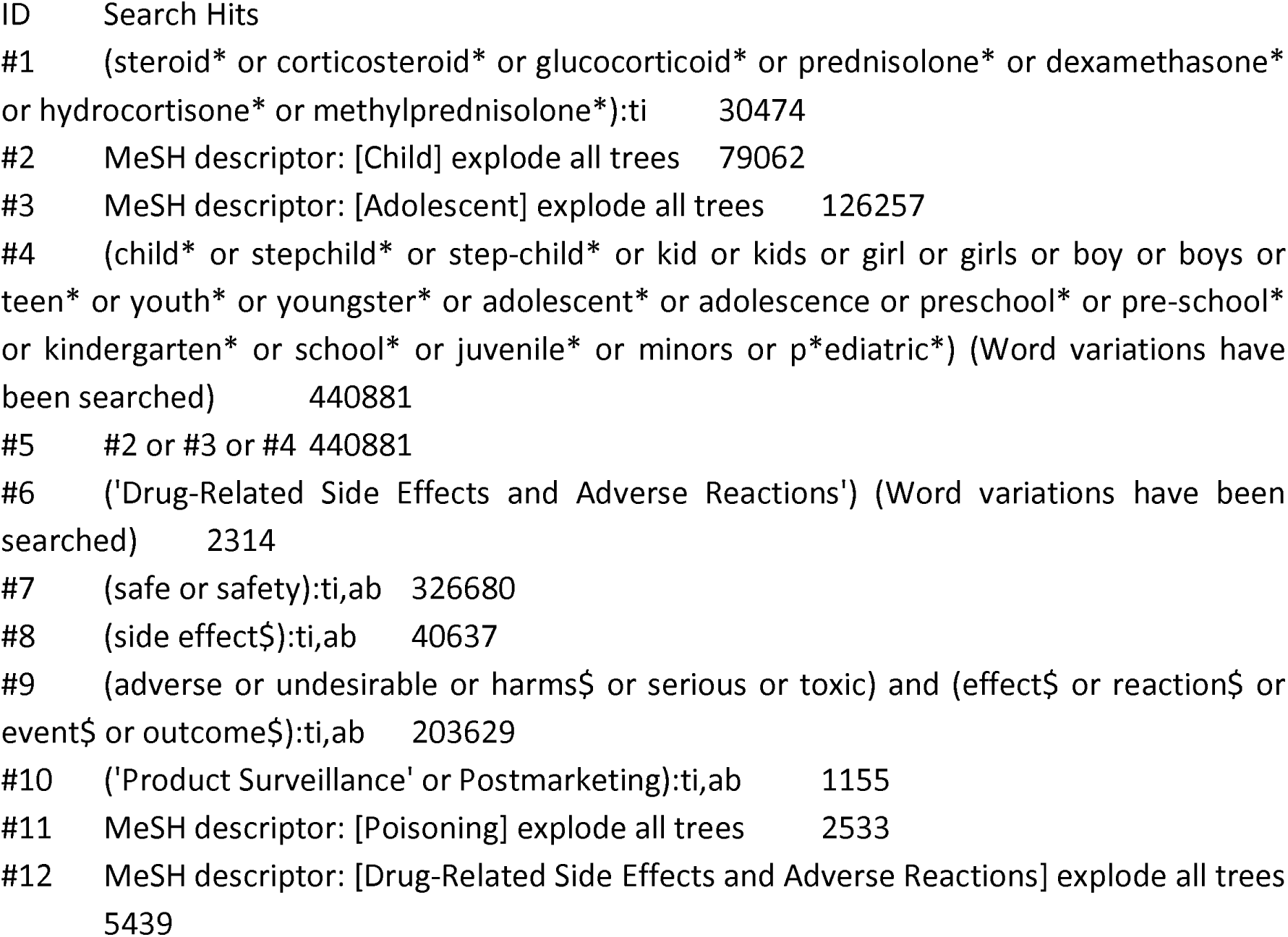
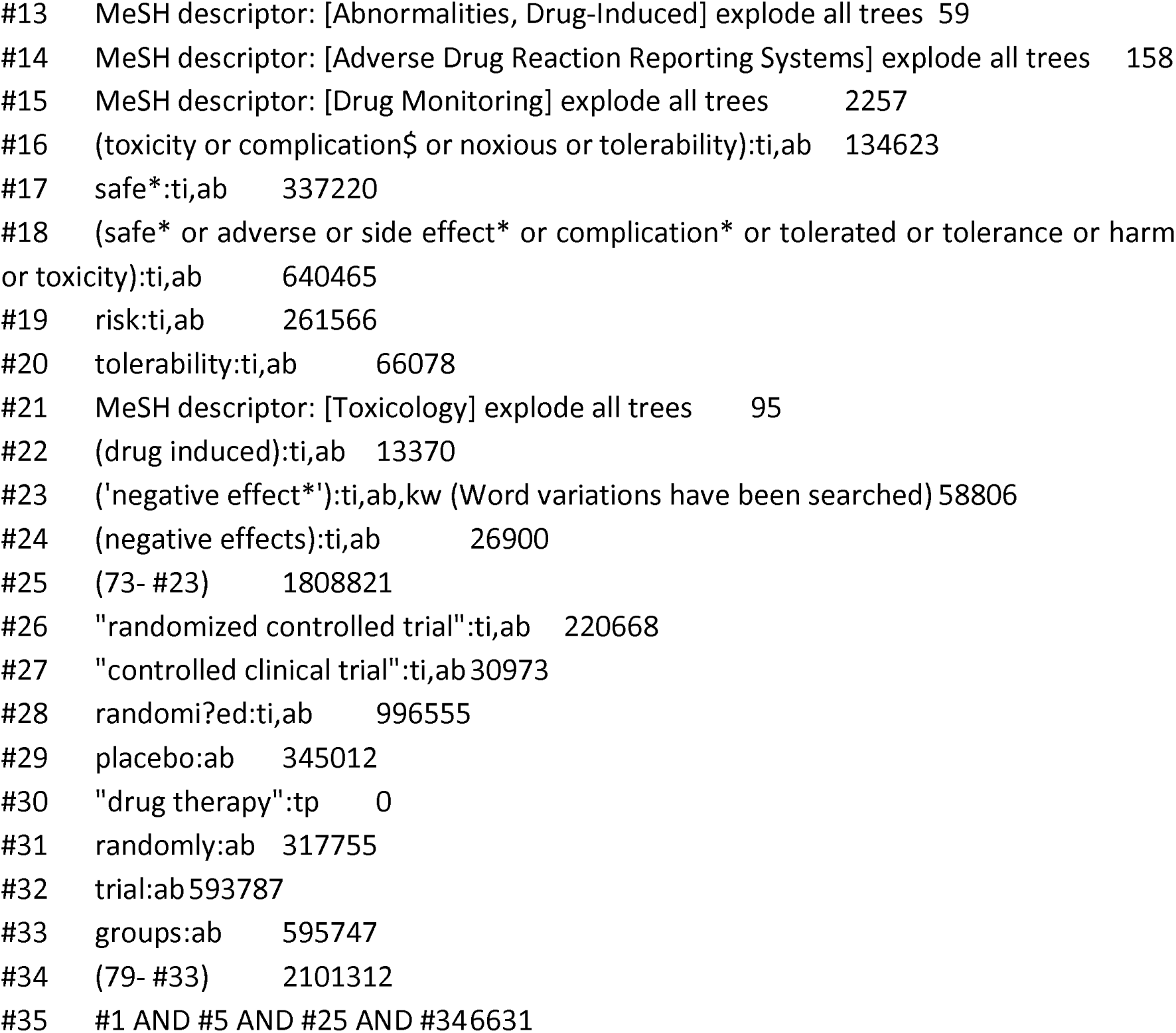

