## Supplemental Material for "Harms of short-course systemic corticosteroids among children and adolescents: a systematic review and meta-analysis of randomized controlled trials"

5. ICEMAN criteria - GI Bleeding........................................................................................................85

***6.*** ***Sensitivity Analyses ...........................................................................................................................88***

***7. Subgroup Analysis …………………………………………………………………………………….93***

### 1. Forest Plots

#### 1.1 Forest plot – Serious Adverse Events

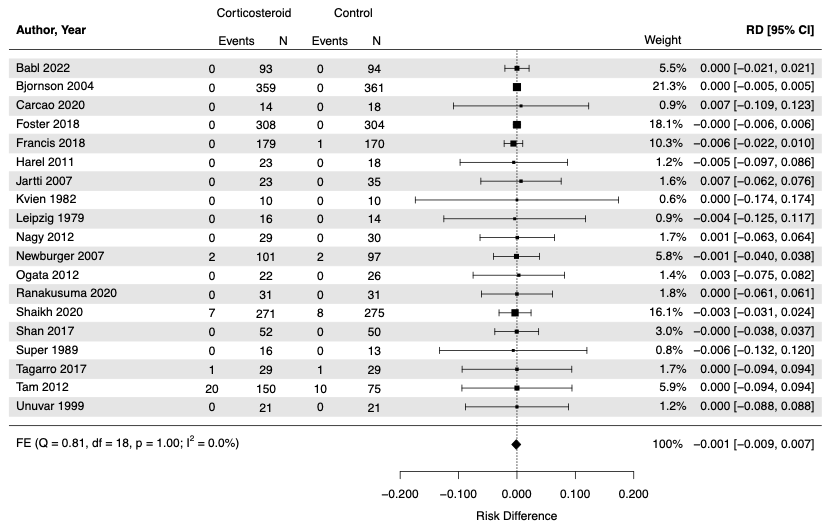

#### 1.2 Forest plot – Adverse Events Leading to discontinuation

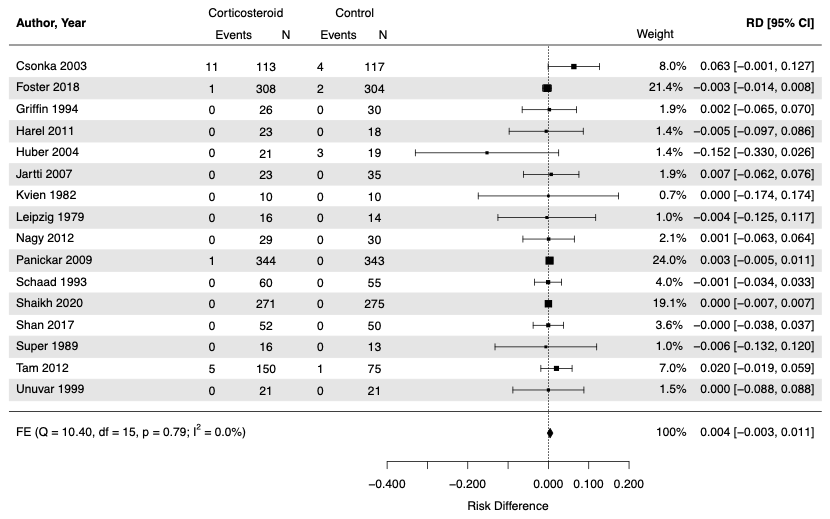

#### 1.3 Forest plot – Abdominal Pain

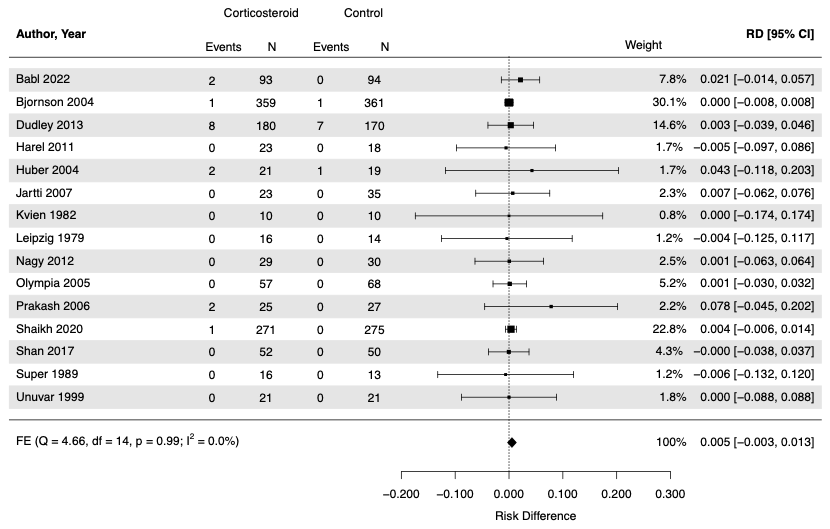

#### 1.4 Forest plot – Diarrhea

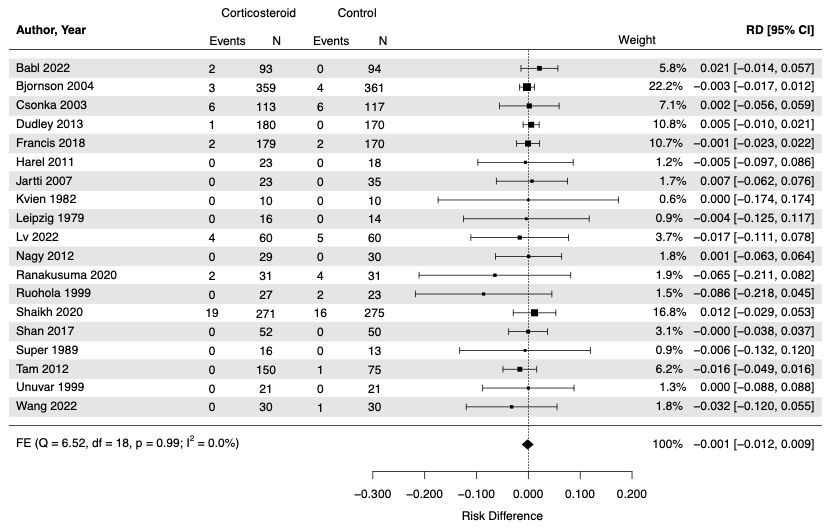

#### 1.5 Forest plot – Gastritis

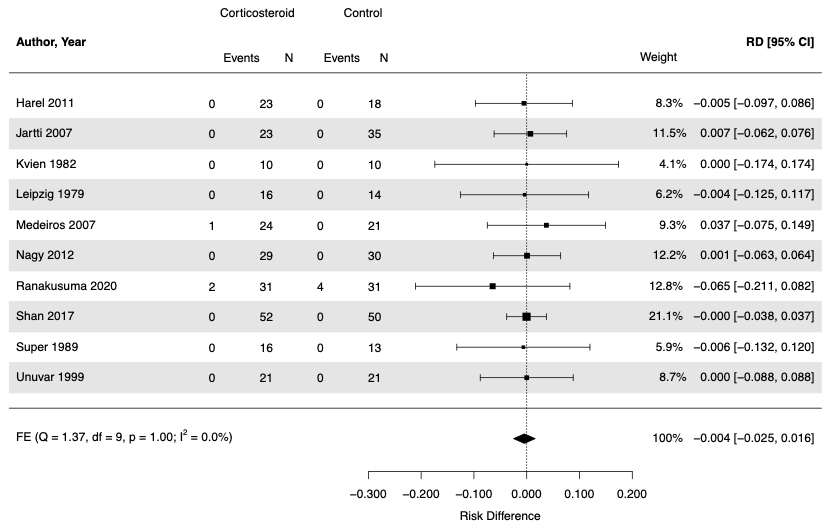

#### 1.6 Forest plot – Gastrointestinal Bleeding

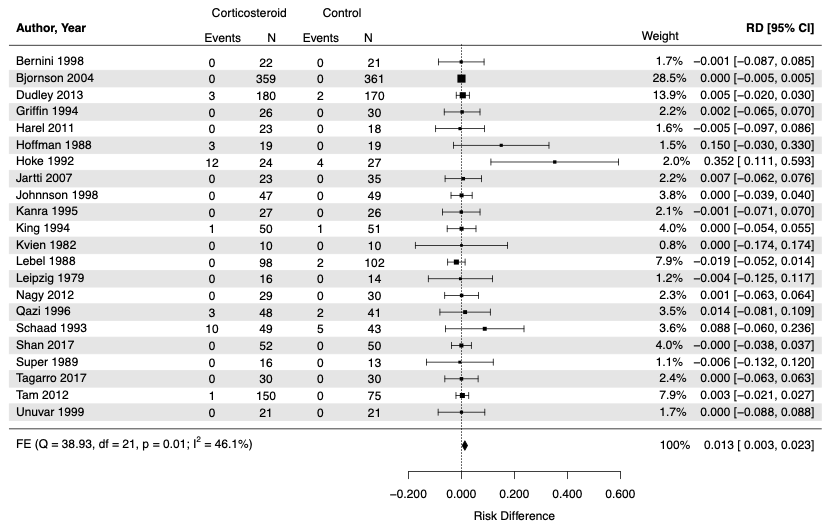

#### 1.7 Forest plot – Hemoccult Positive Stool

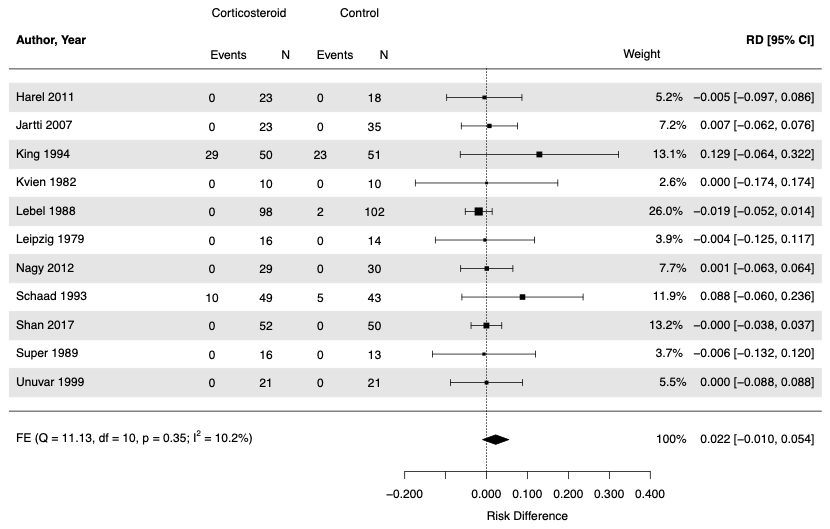

#### 1.8 Forest plot – Intussusception

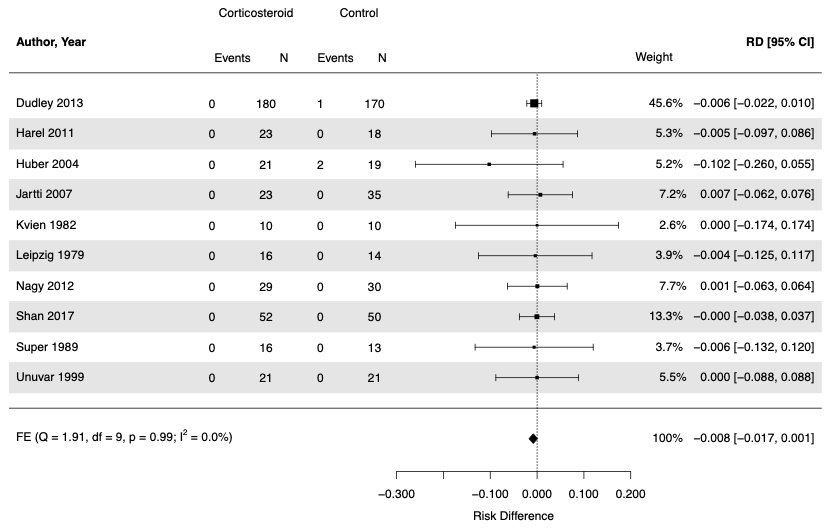

#### 1.9 Forest plot – Nausea

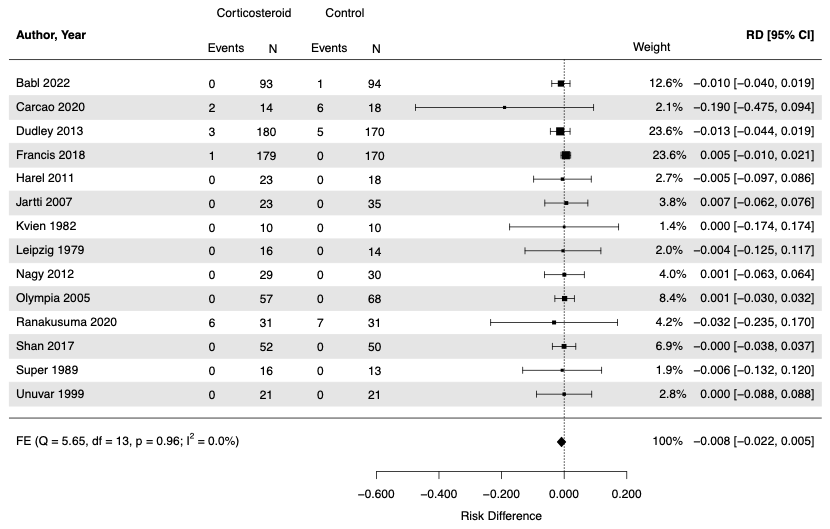

#### 1.10 Forest plot – Vomiting

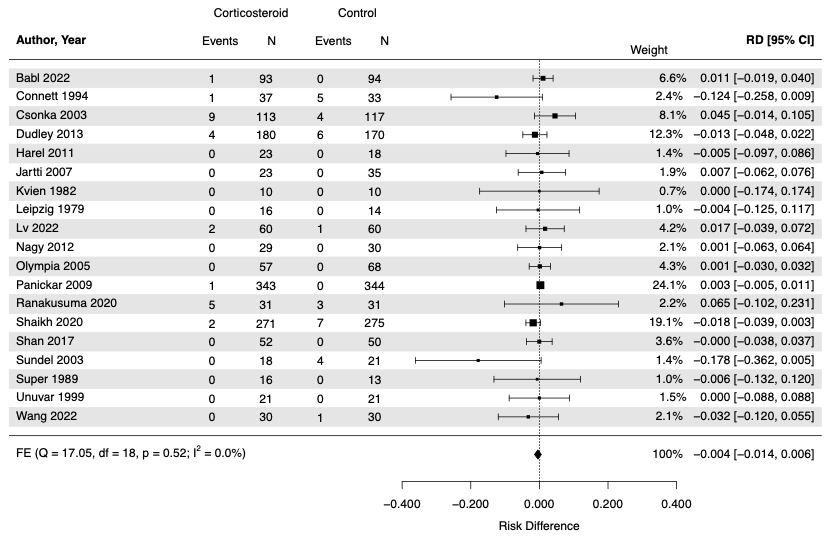

#### 1.11 Forest plot – Anemia

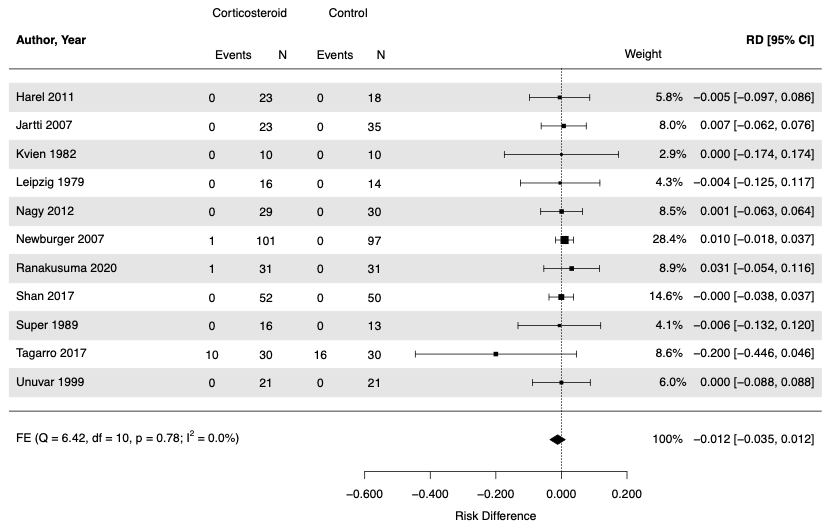

#### 1.12 Forest plot – Candidiasis

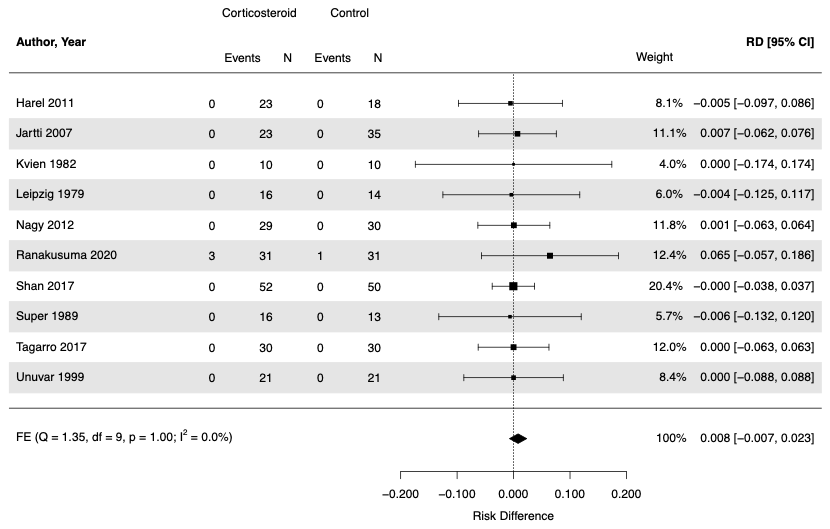

#### 1.13 Forest plot – Change in behaviour

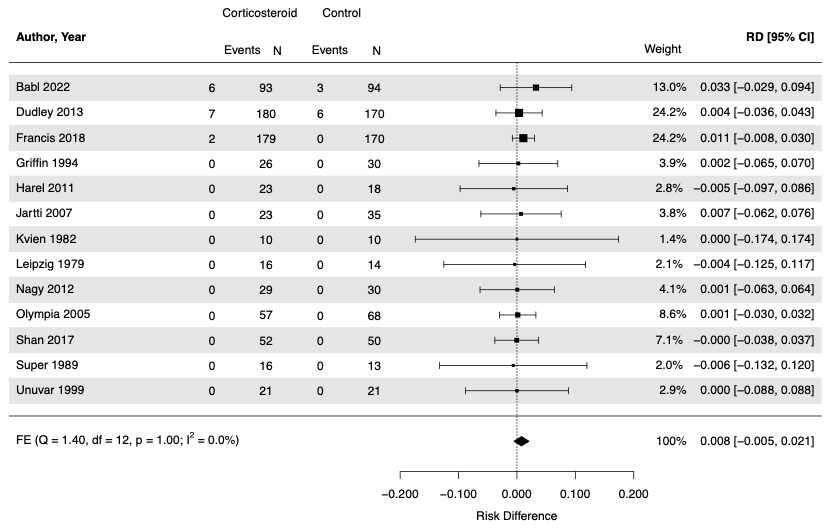

#### 1.14 Forest plot – Congestive Heart Failure

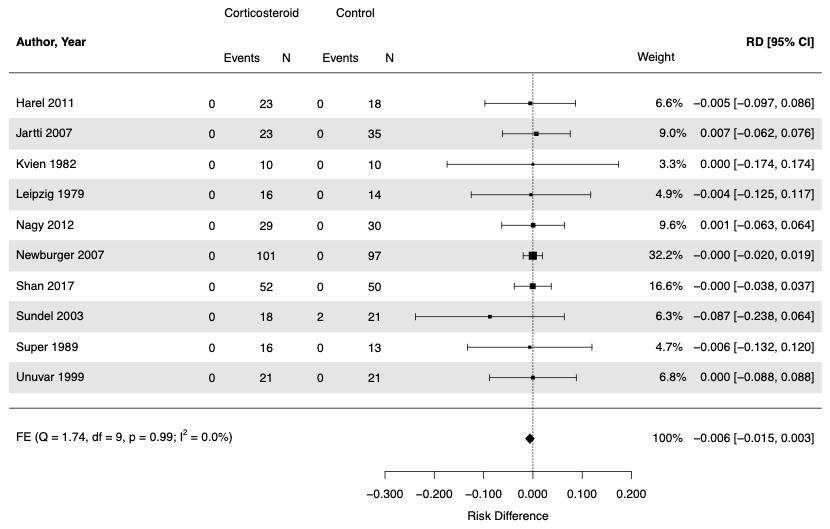

#### 1.15 Forest plot – Convulsion/Seizure

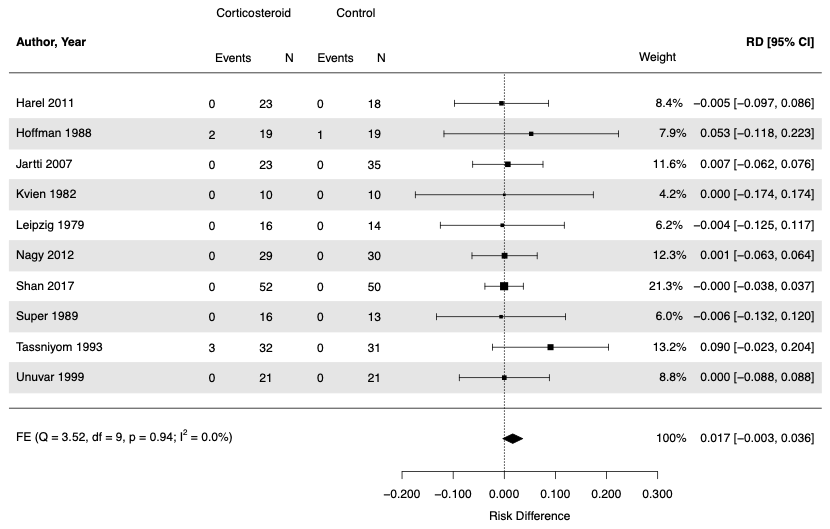

#### 1.16 Forest plot – Decreased Appetite

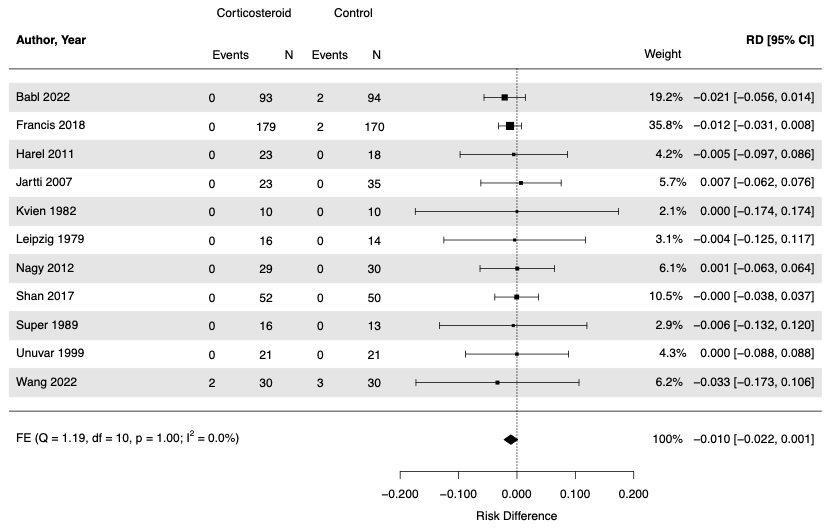

#### 1.17 Forest plot – Dizziness

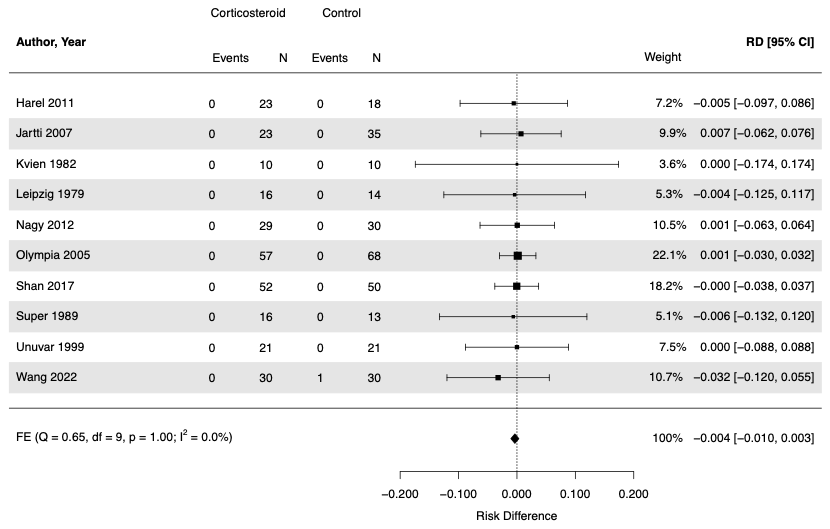

#### 1.18 Forest plot – Fatigue

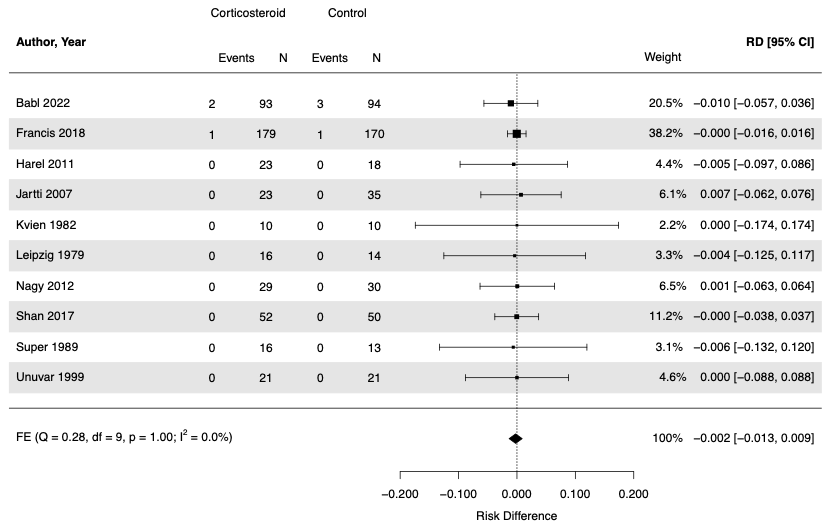

#### 1.19 Forest plot – Febrile Convulsion

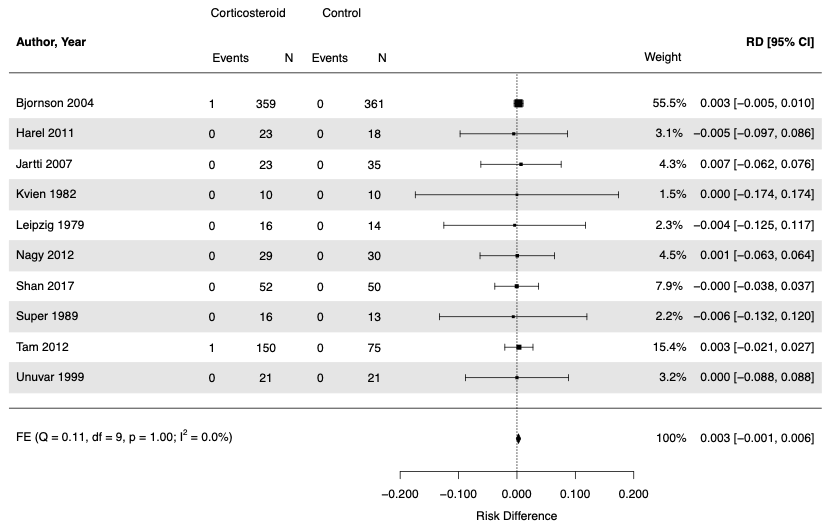

#### 1.20 Forest plot – Glycosuria

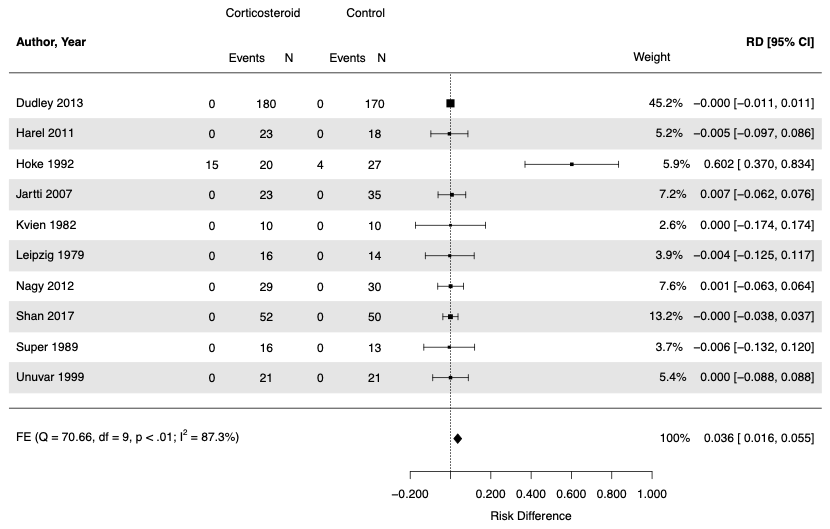

#### 1.21 Forest plot – Headache

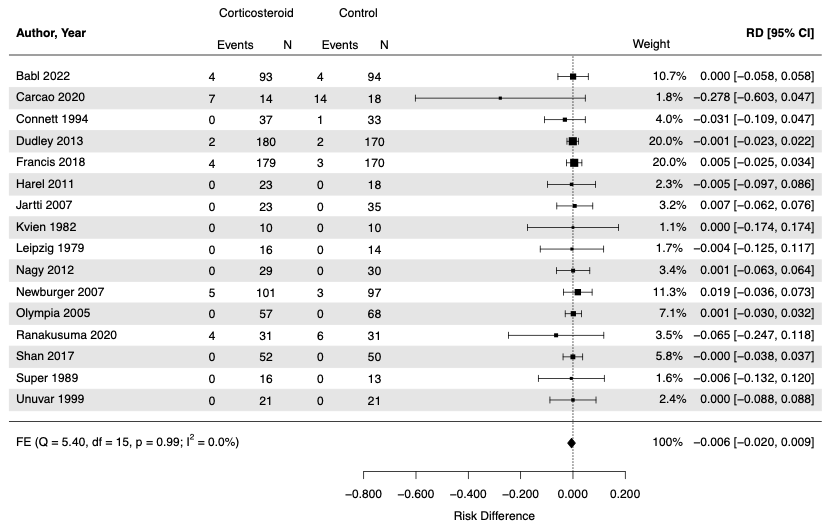

#### 1.22 Forest plot – Hyperglycemia

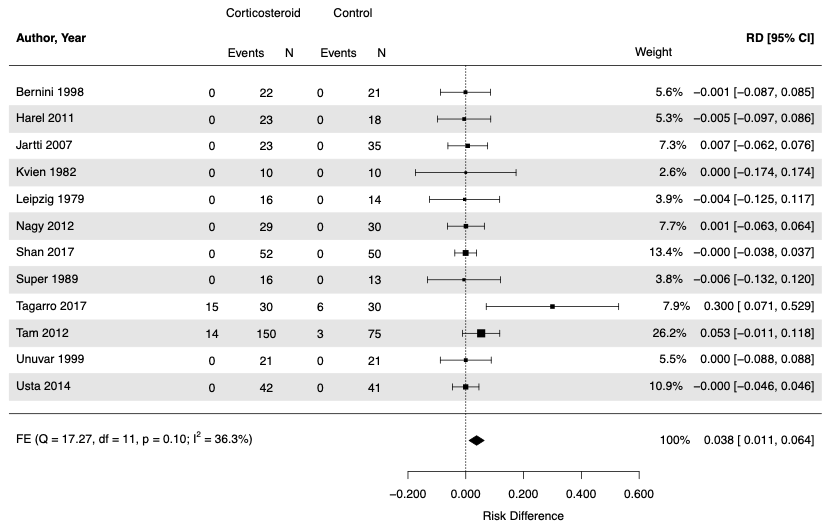

#### 1.23 Forest plot – Hypertension

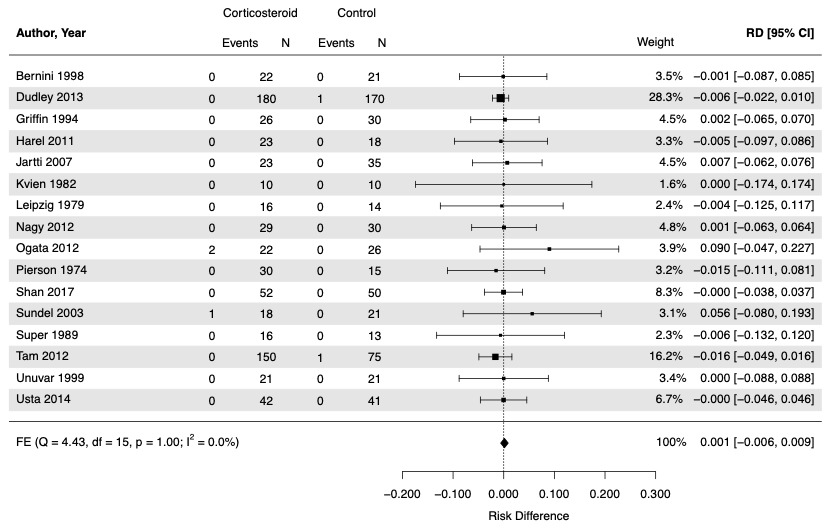

#### 1.24 Forest plot – Increased Appetite

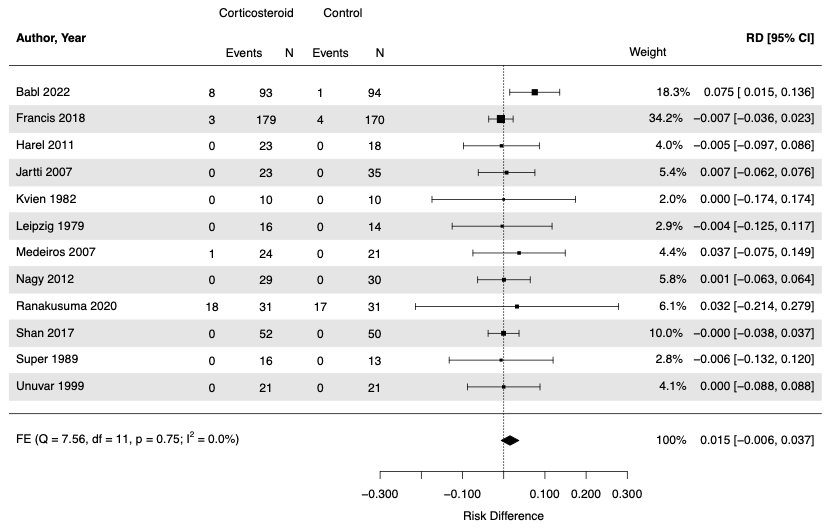

#### 1.25 Forest plot – Irritability

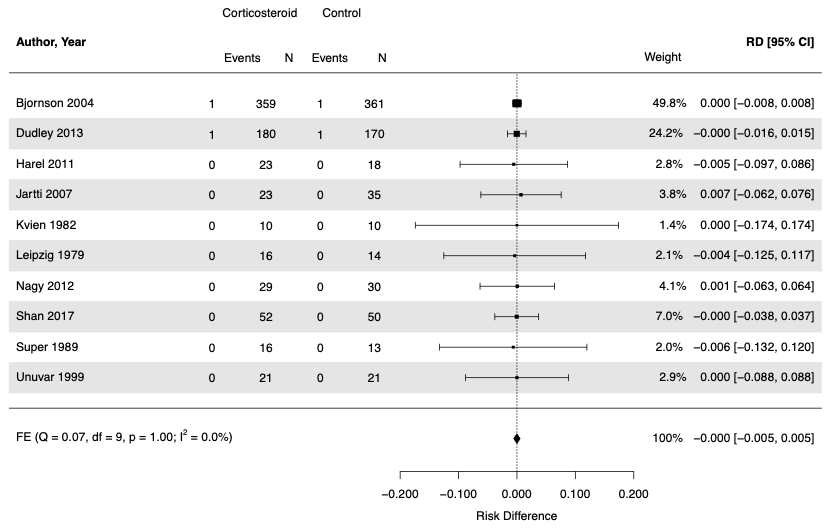

#### 1.26 Forest plot – Local Site Pain

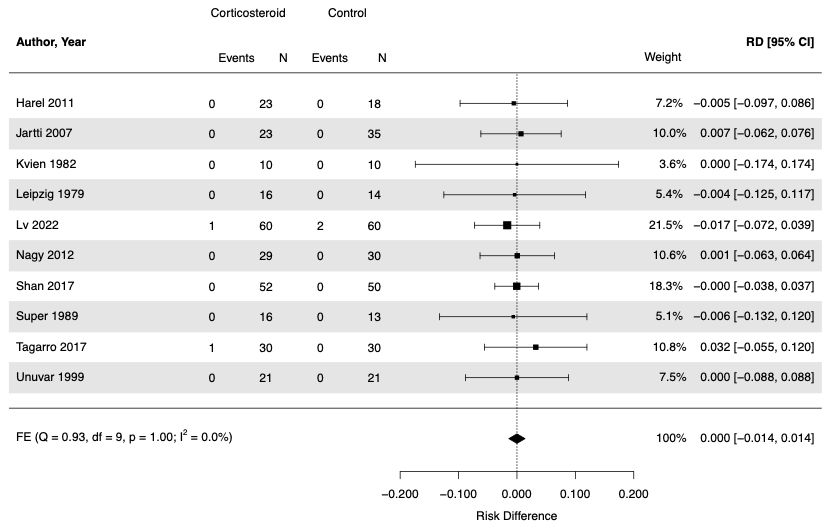

#### 1.27 Forest plot – Musculoskeletal Pain

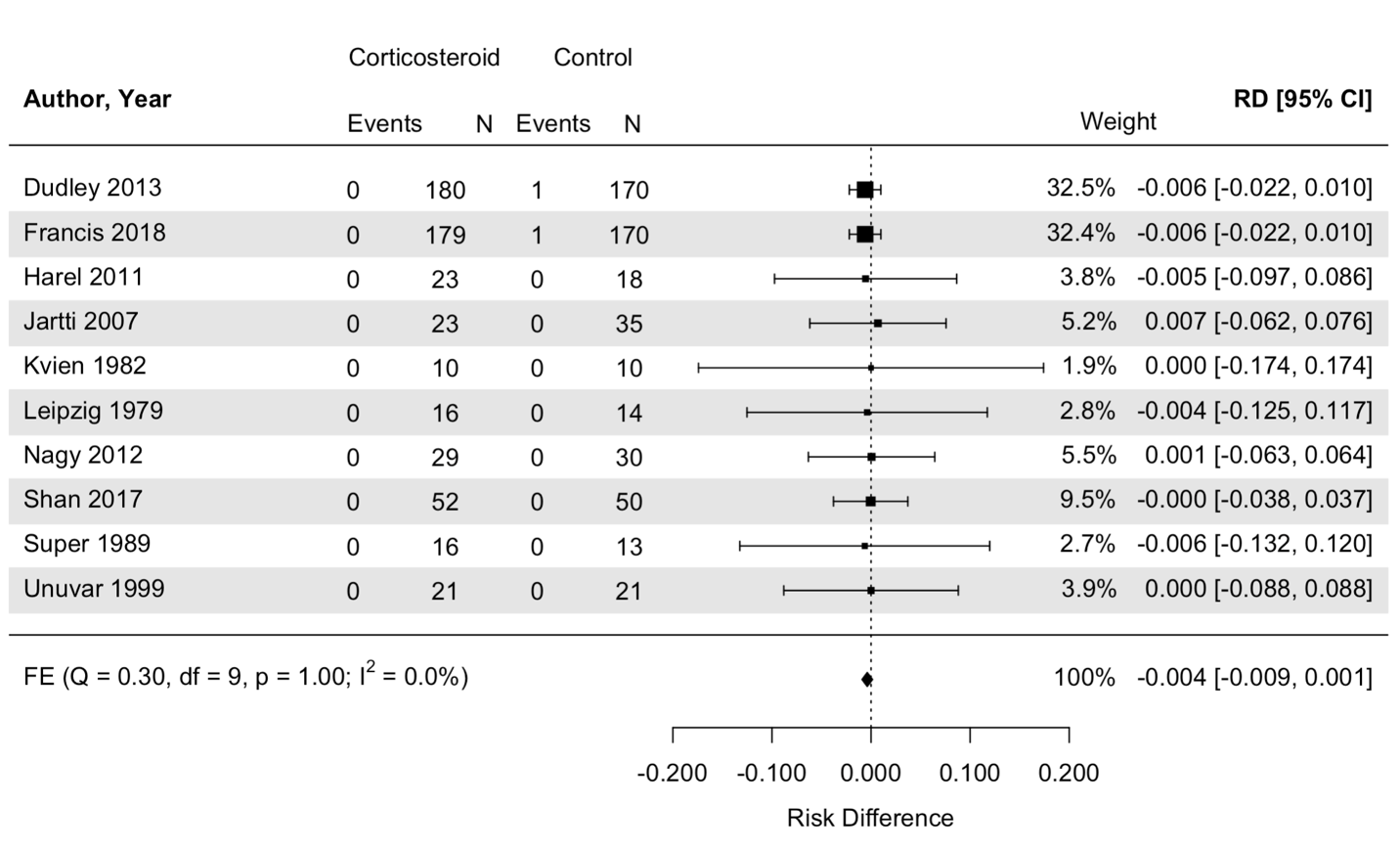

#### 1.28 Forest plot – Myalgia

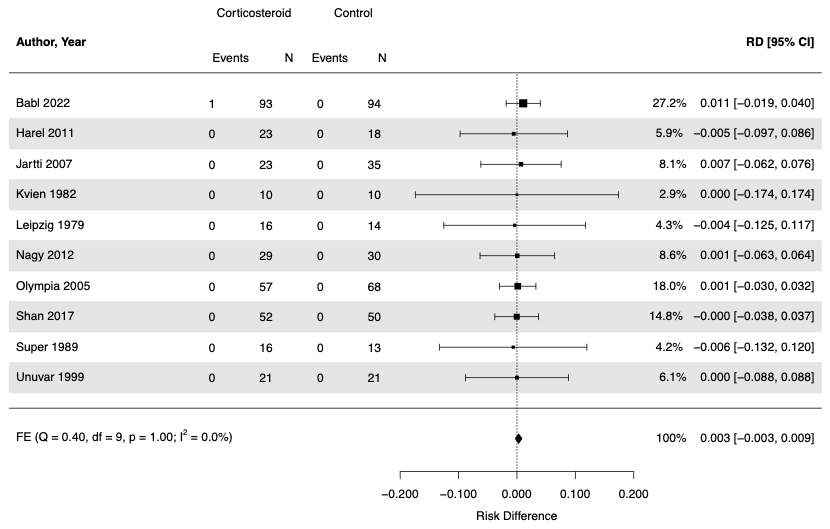

#### 1.29 Forest plot – Otitis Media

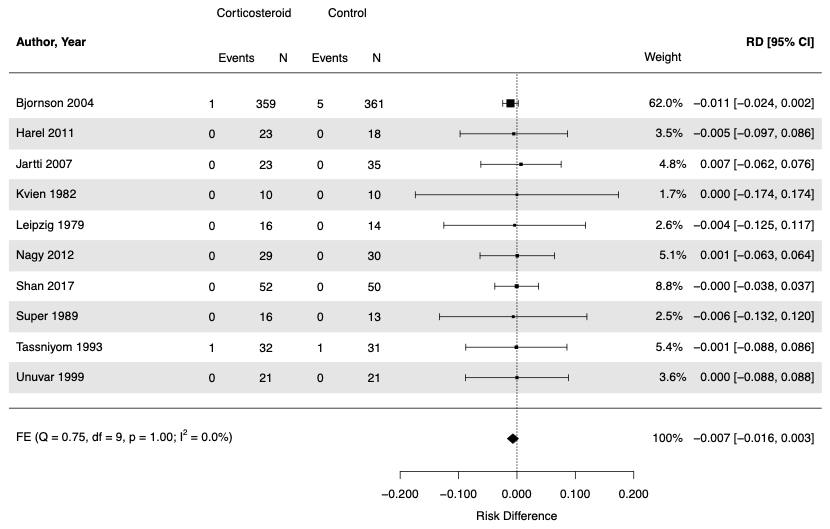

#### 1.30 Forest plot – Pneumonia

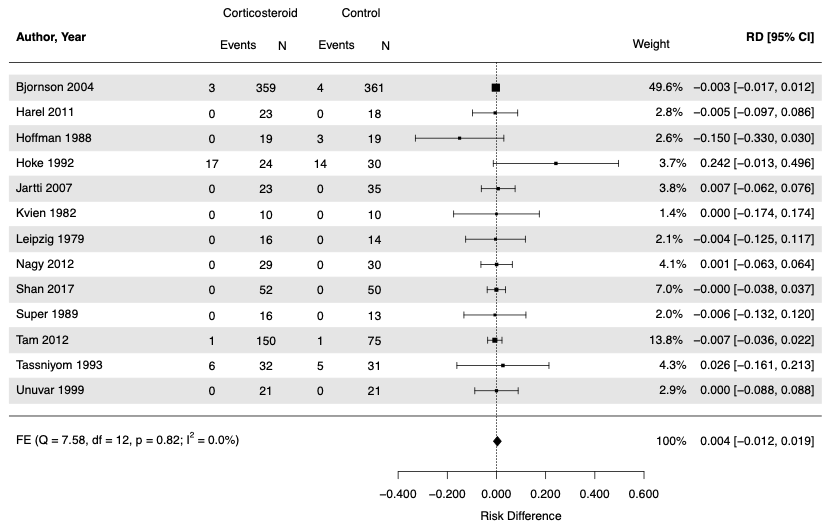

#### 1.31 Forest plot – Polyuria

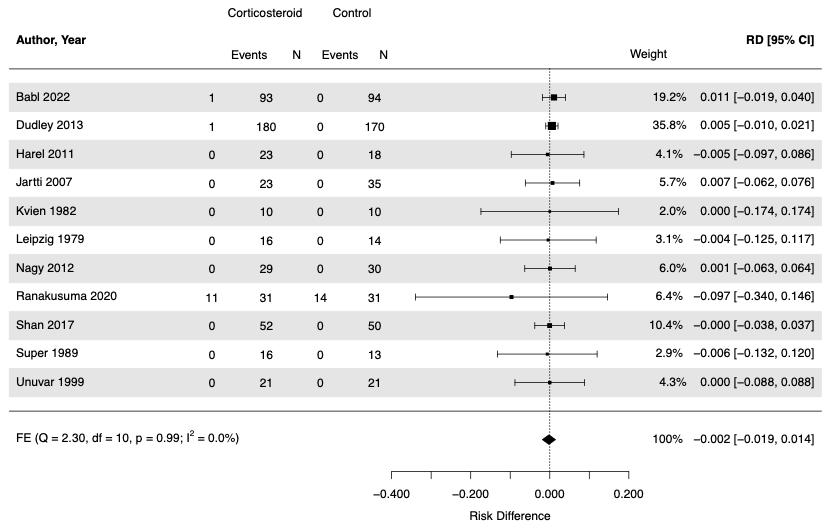

#### 1.32 Forest plot – Rash or Urticaria

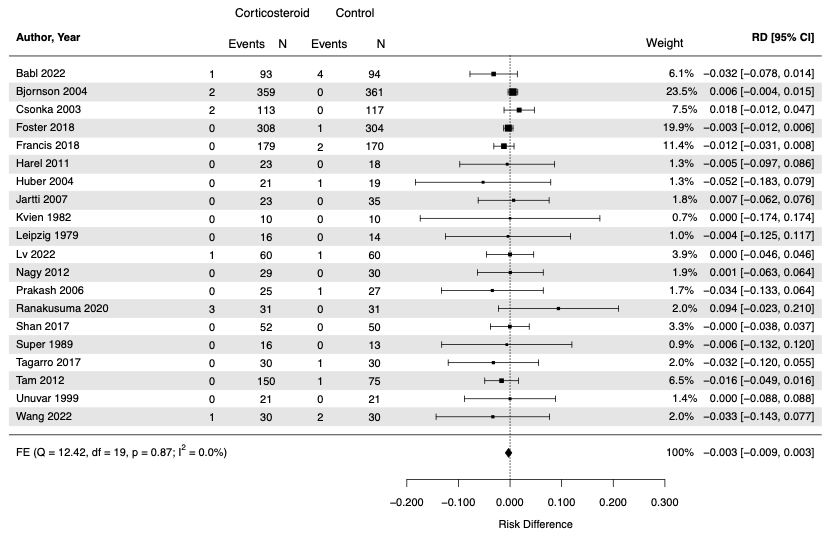

#### 1.32 Forest plot – Secondary (Opportunistic) Infection

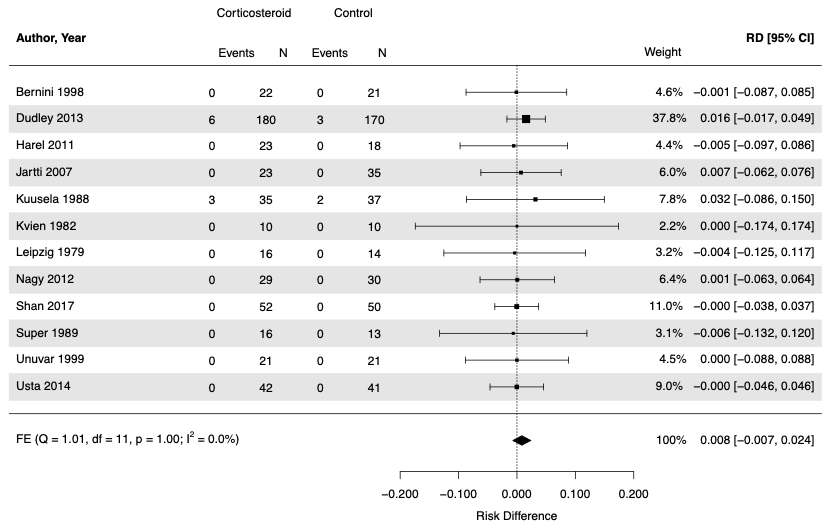

#### 1.33 Forest plot – Secondary Fever

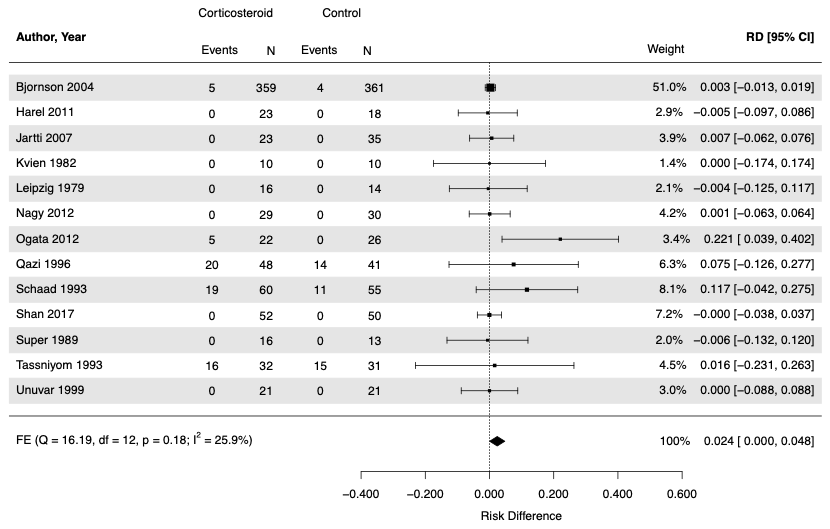

#### 1.34 Forest plot – Sleep Problems

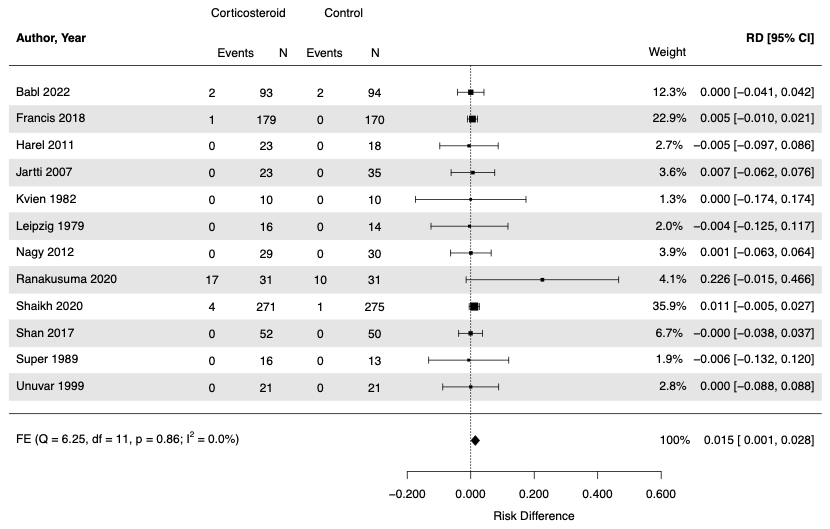

#### 1.35 Forest plot – Tremor or Hyperactivity

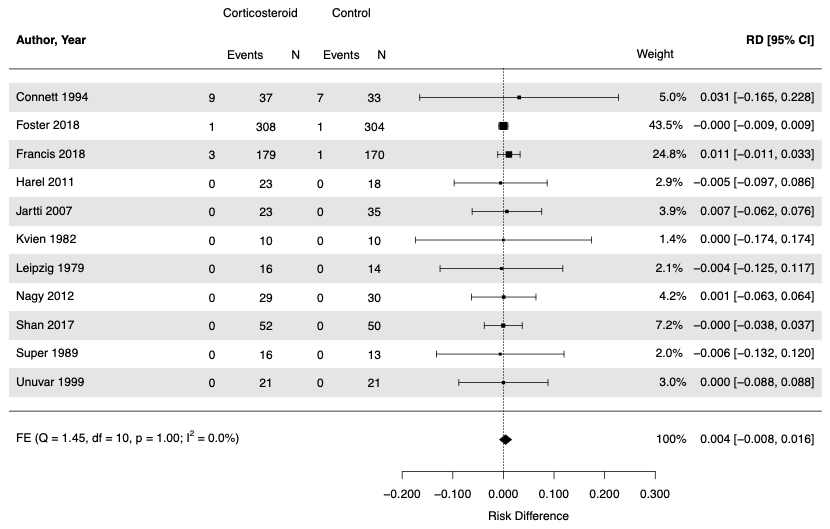

#### 1.36 Forest plot – Urinary Tract Infection

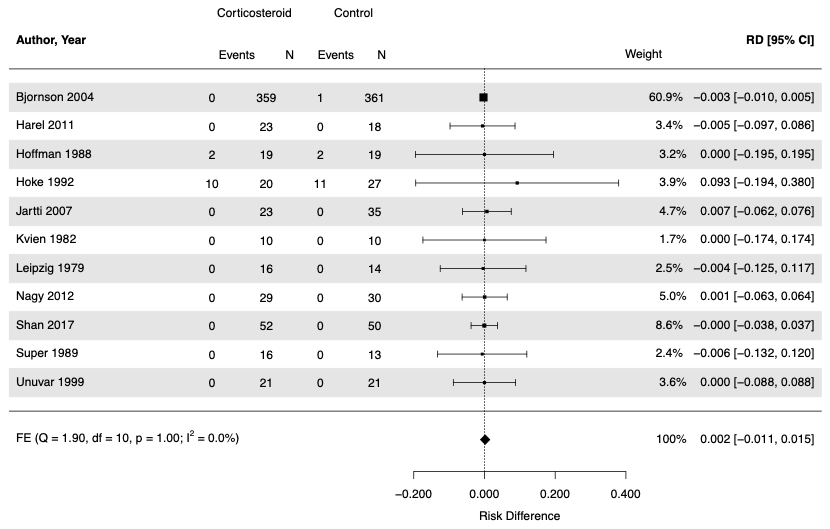

### 2. Funnel Plots

#### 2.1 Funnel plot – Serious Adverse Events

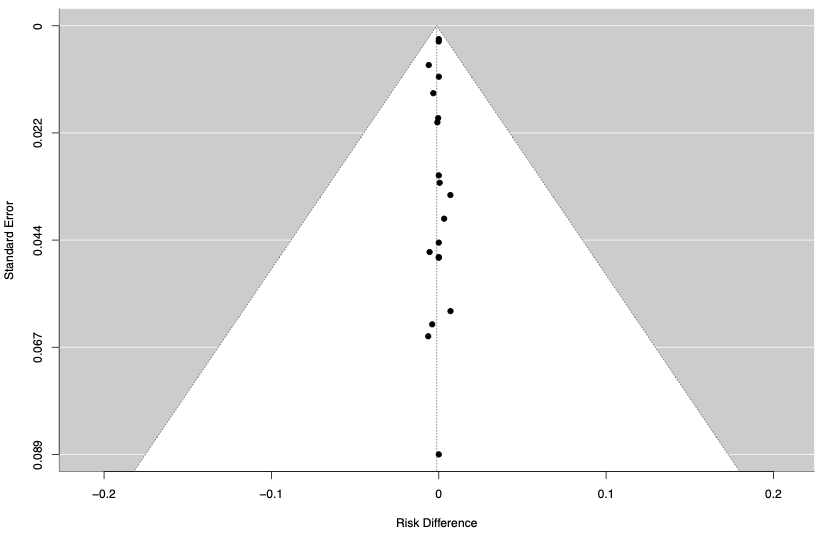

#### 2.2 Funnel plot – Adverse Events Leading to discontinuation

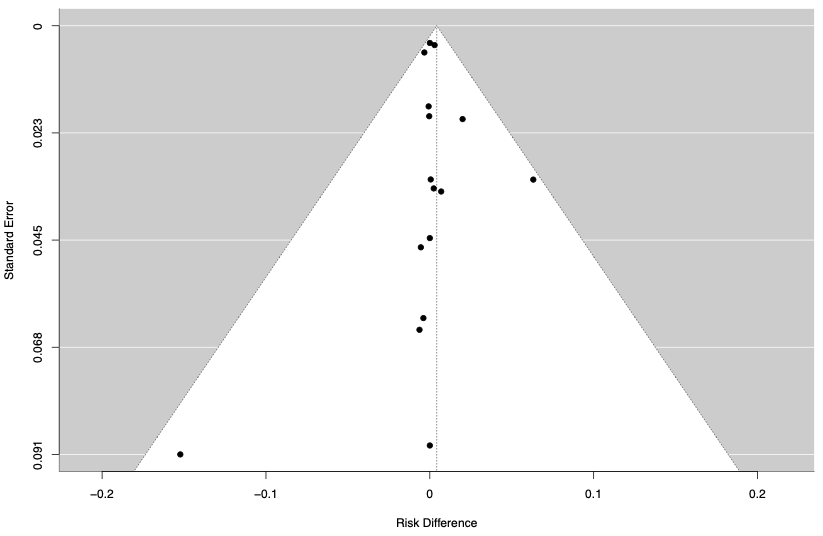

#### 2.3 Funnel plot – Abdominal Pain

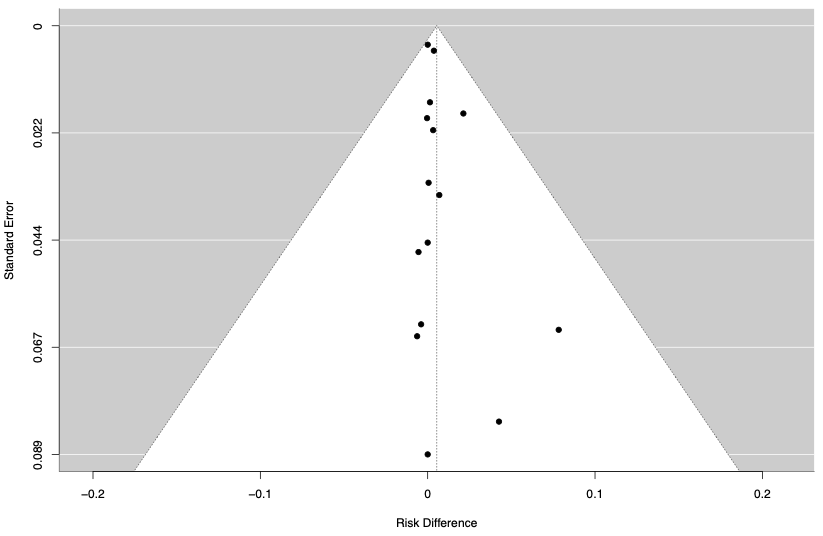

#### 2.4 Funnel plot – Diarrhea

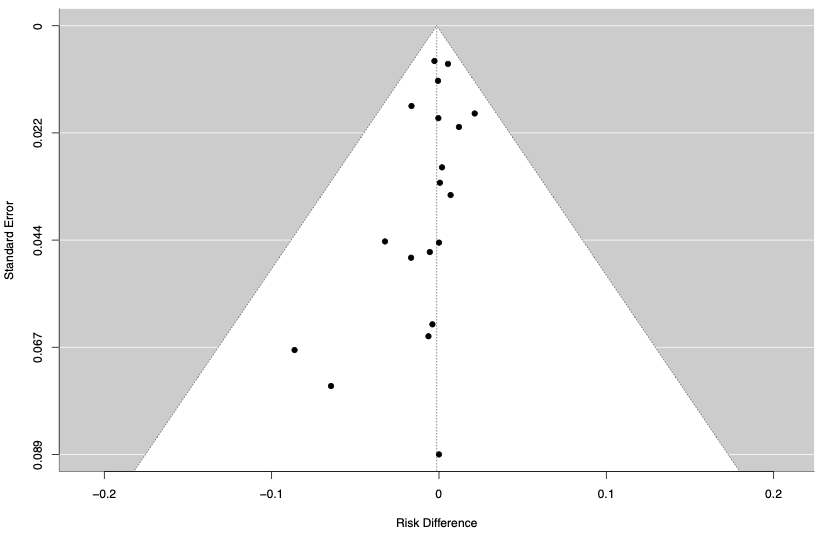

#### 2.5  Funnel plot – Gastritis

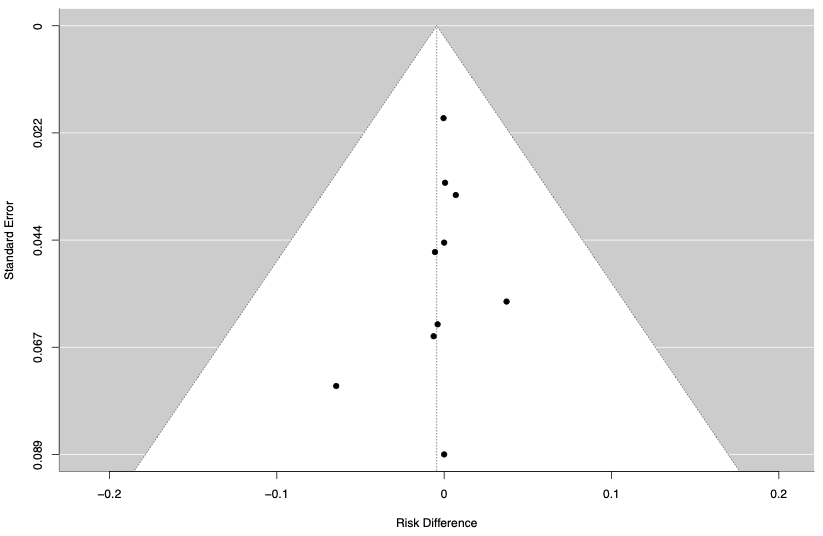

#### 2.6 Funnel plot – Gastrointestinal Bleeding

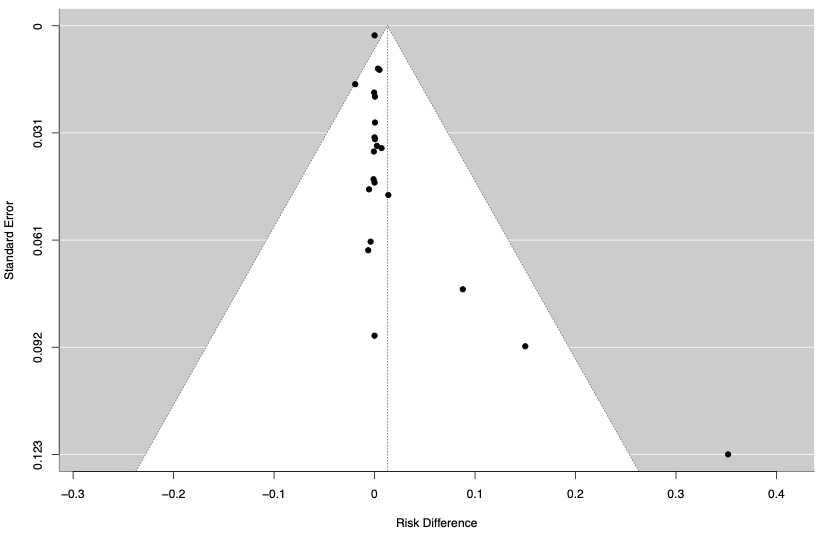

#### 2.7 Funnel plot – Hemoccult Positive Stool

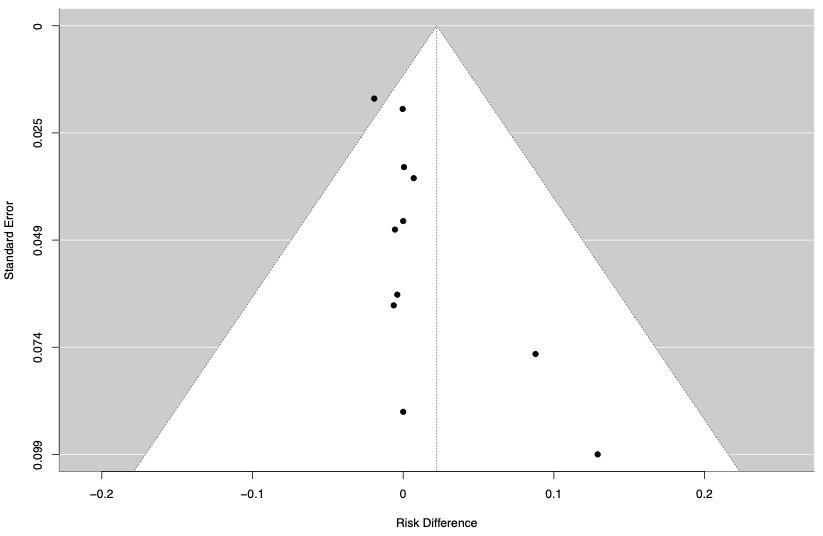

#### 2.8 Funnel plot – Intussusception

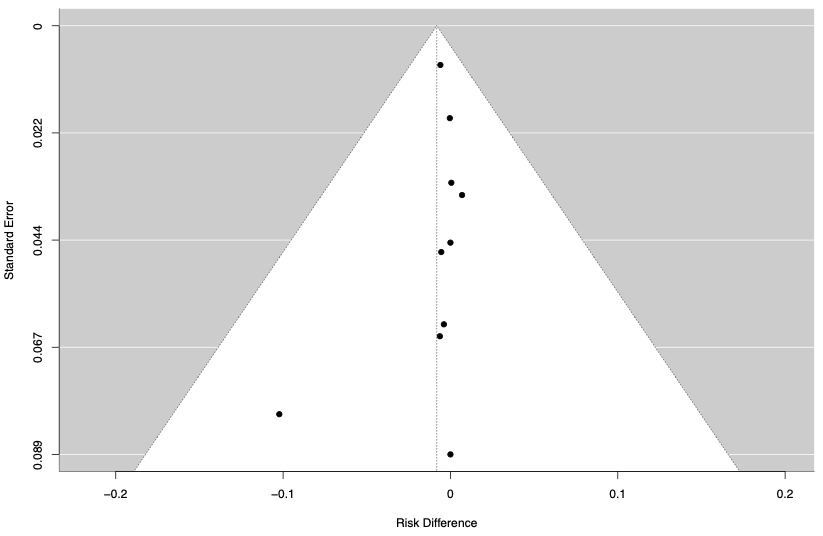

#### 2.9  Funnel plot – Nausea

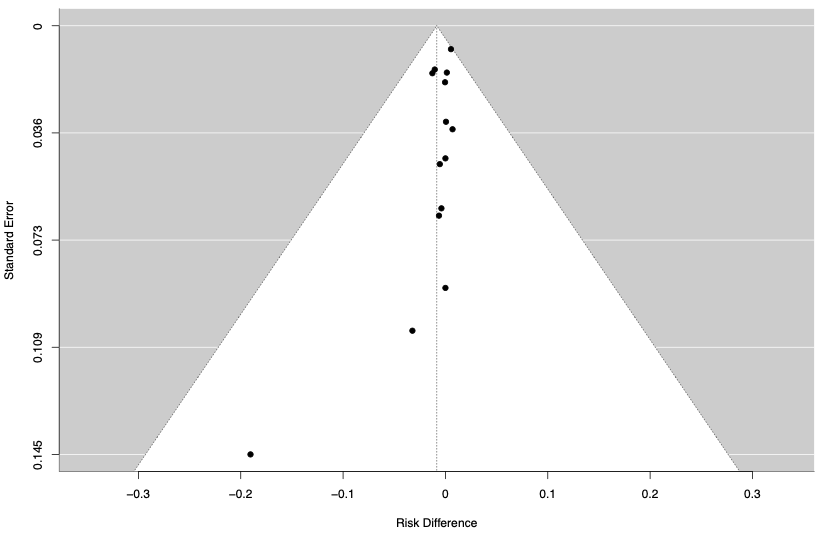

#### 2.10  Funnel plot – Vomiting

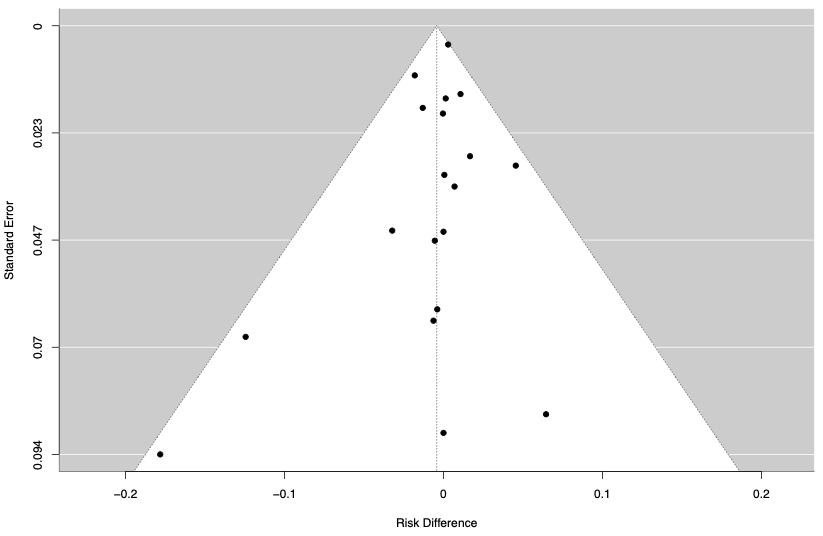

#### 2.11  Funnel plot – Anemia

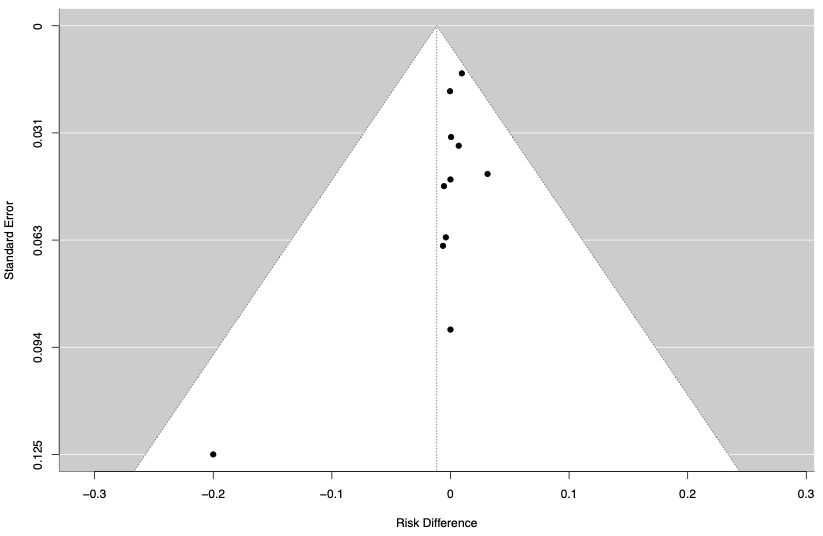

#### 2.12 Funnel plot – Candidiasis

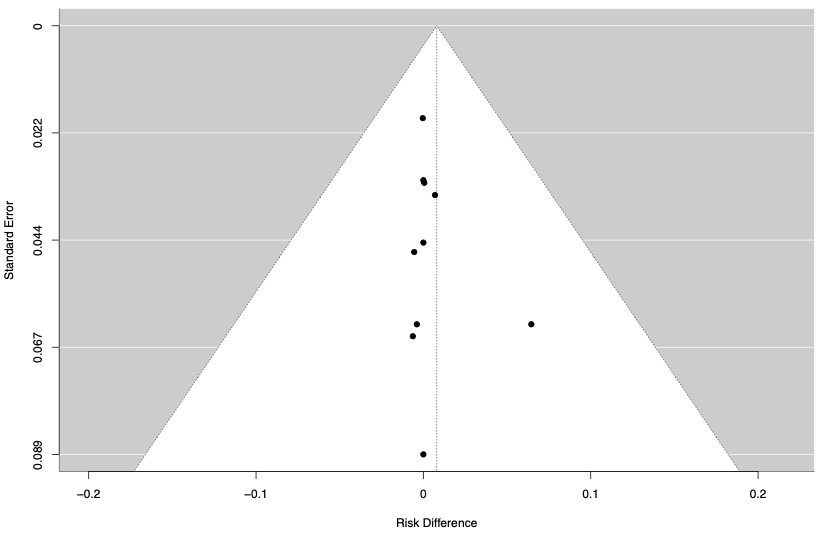

#### 2.13 Funnel plot – Change in behaviour

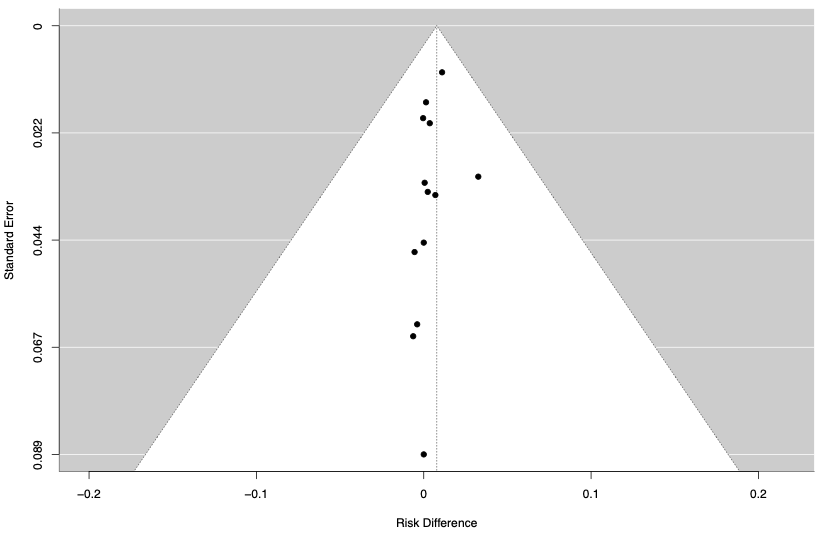

#### 2.14 Funnel plot – Congestive Heart Failure

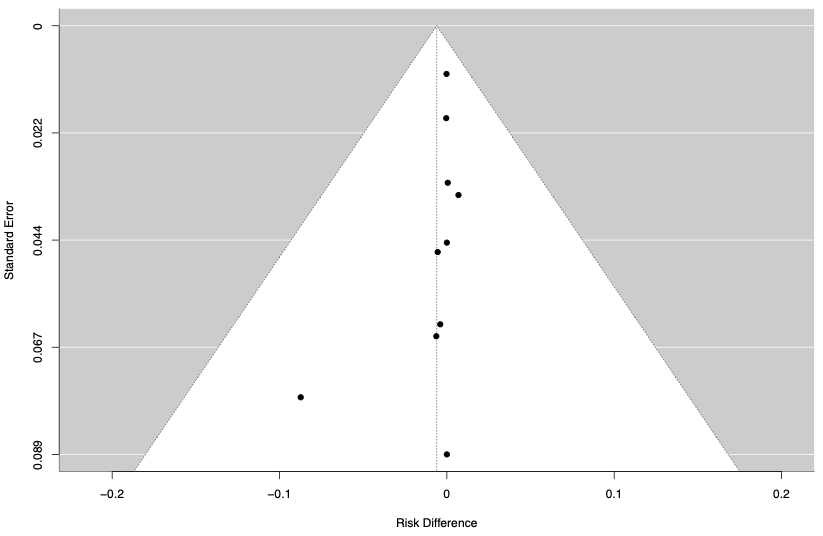

#### 2.15 Funnel plot – Convulsion/Seizure

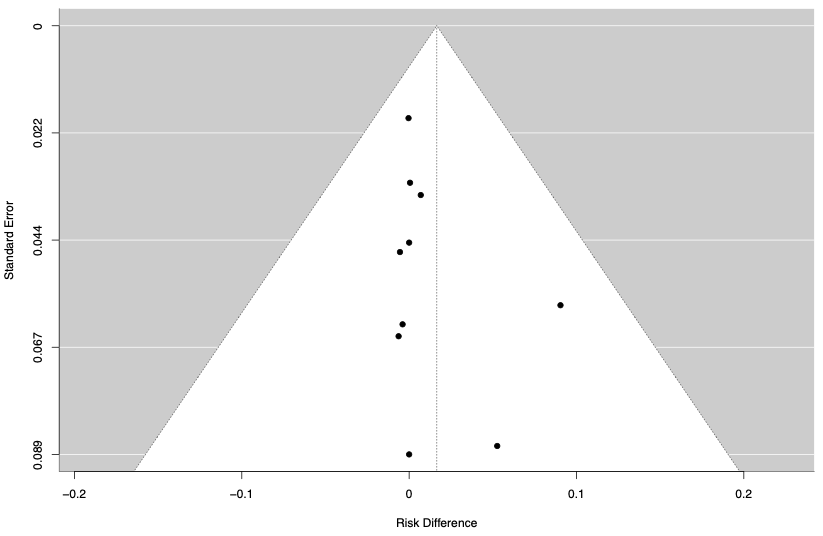

#### 2.16 Funnel plot – Decreased Appetite

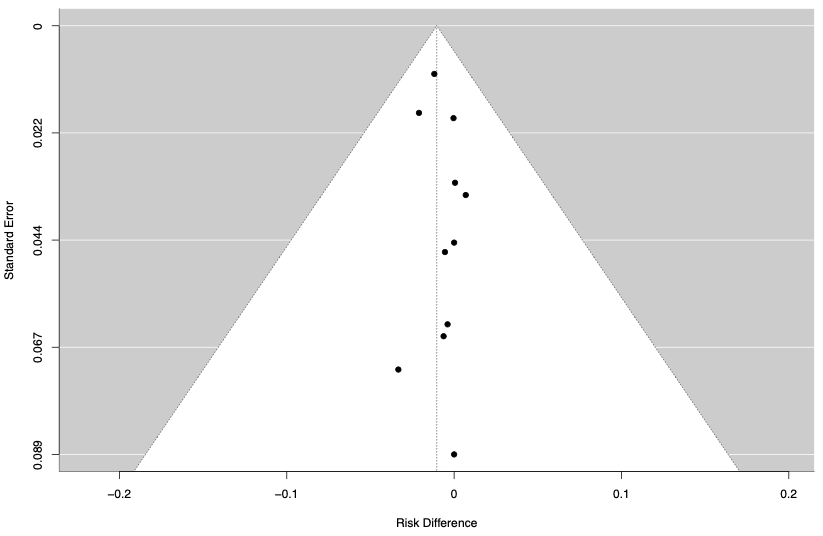

#### 2.17 Funnel plot – Dizziness

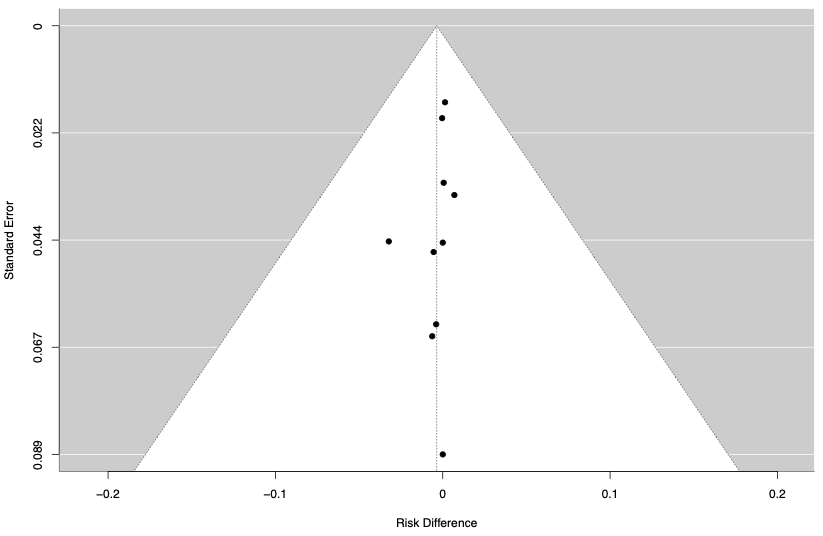

#### 2.18 Funnel plot – Fatigue

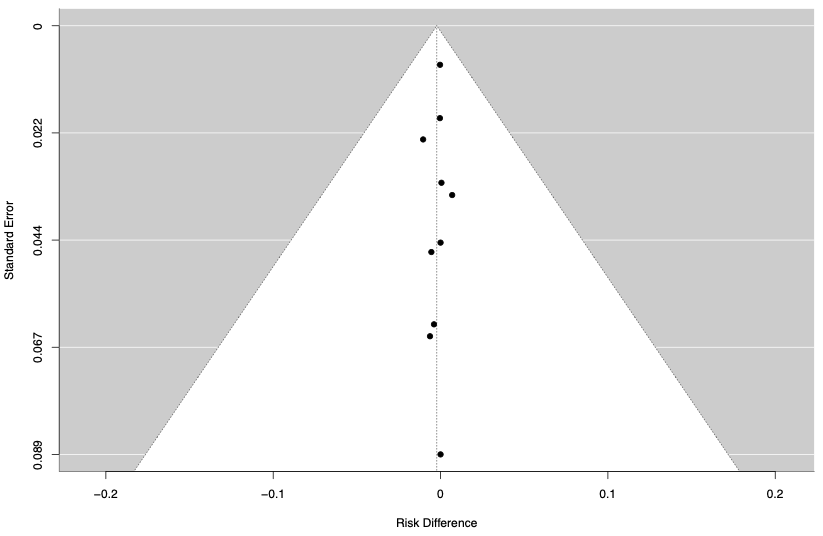

#### 2.19 Funnel plot – Febrile Convulsion

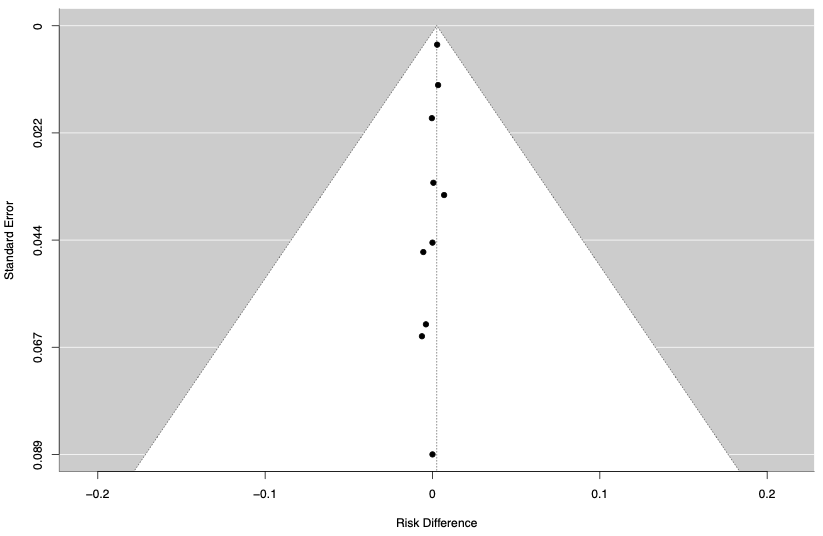

#### 2.20 Funnel plot – Glycosuria

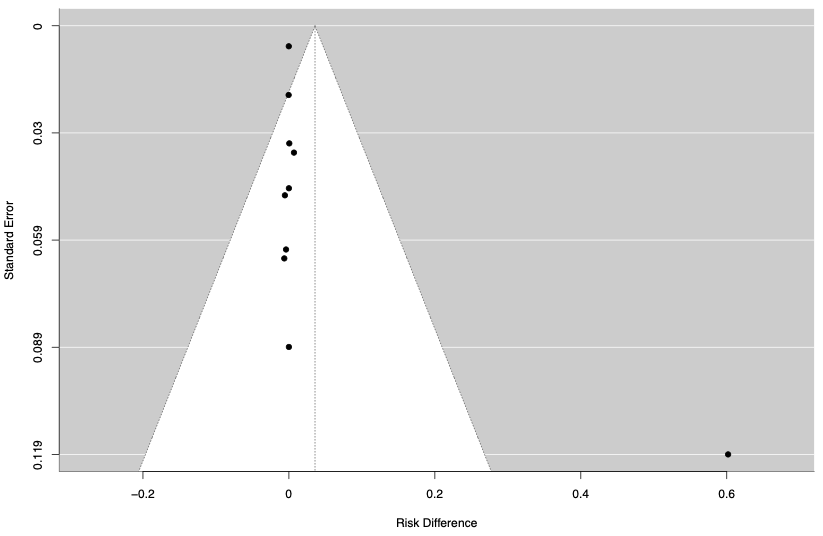

#### 2.21 Funnel plot – Headache

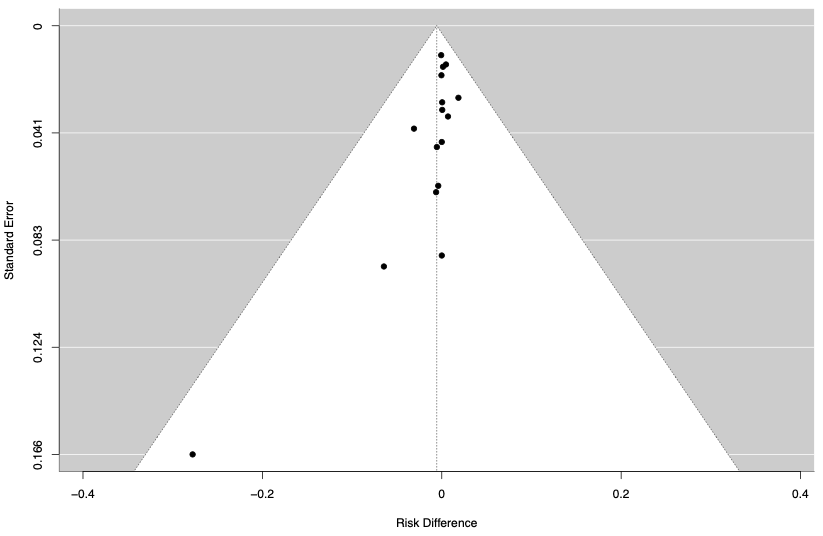

#### 2.22 Funnel plot – Hyperglycemia

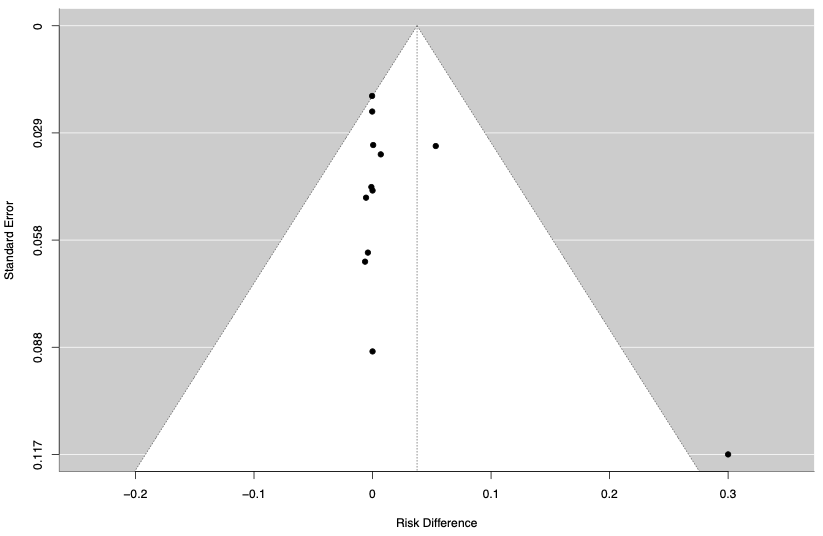

#### 2.23 Funnel plot – Hypertension

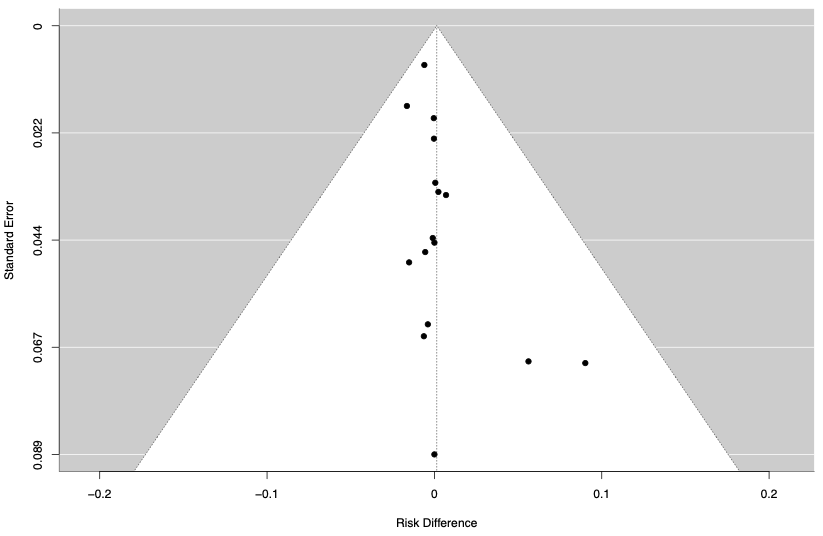

#### 2.24 Funnel plot – Increased Appetite

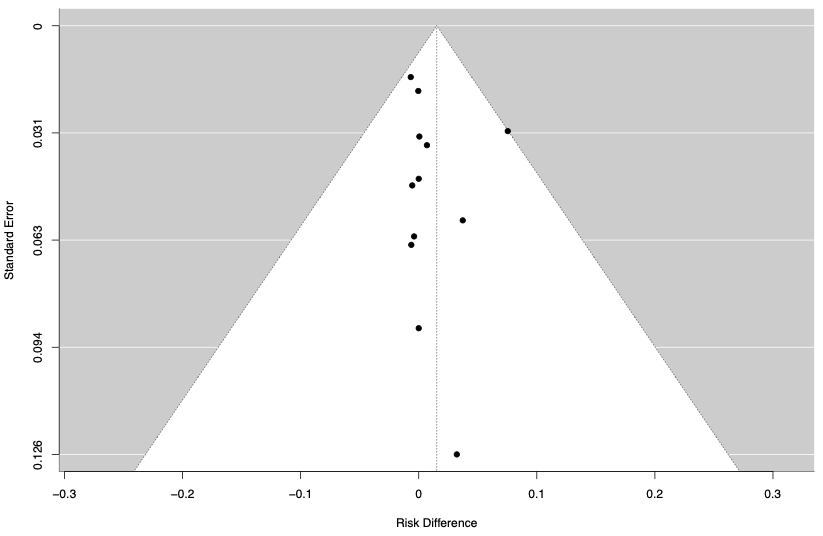

#### 2.25 Funnel plot – Irritability

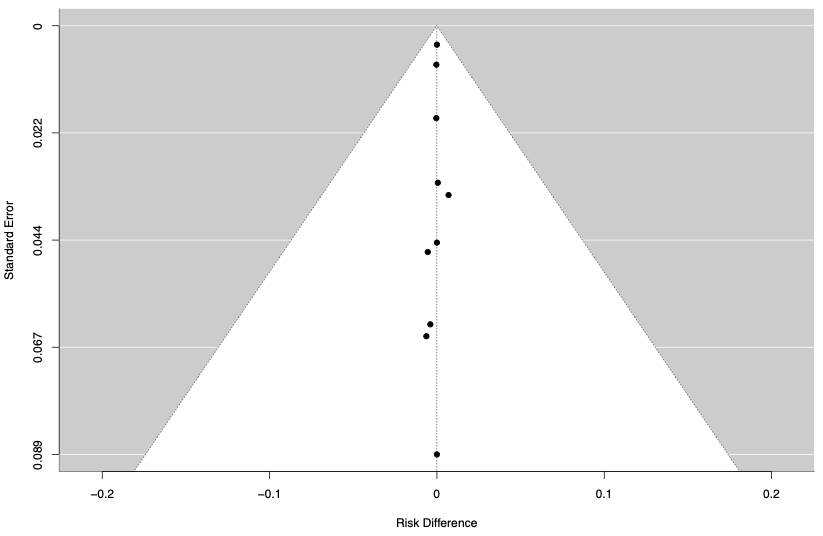

#### 2.26 Funnel plot – Local Site Pain

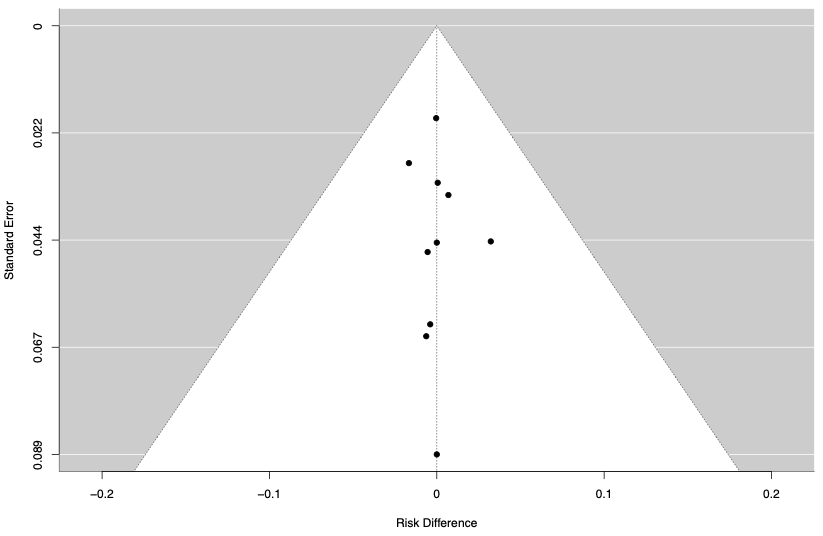

#### 2.27 Funnel plot – Musculoskeletal Pain

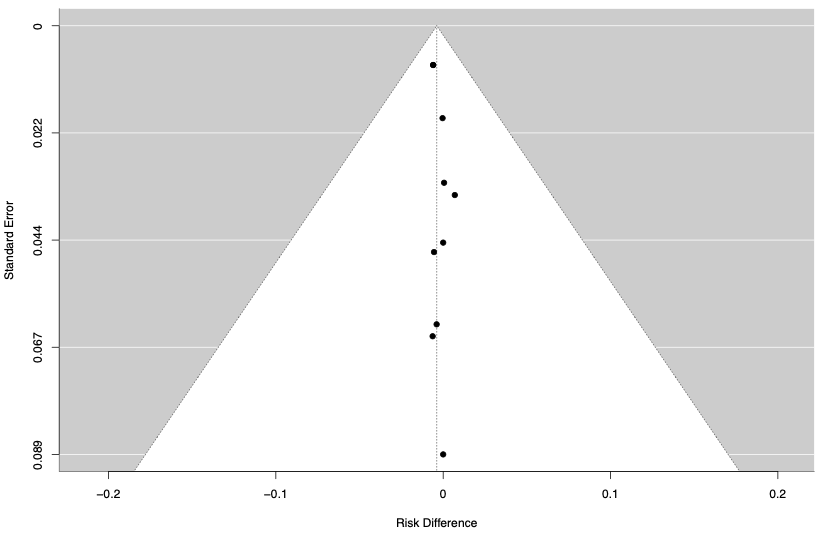

#### 2.28 Funnel plot – Myalgia

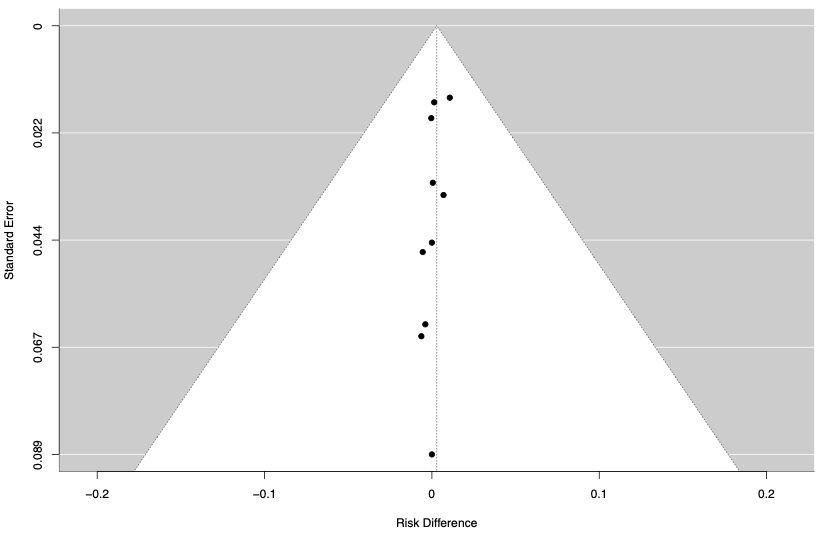

#### 2.29 Funnel plot – Otitis Media

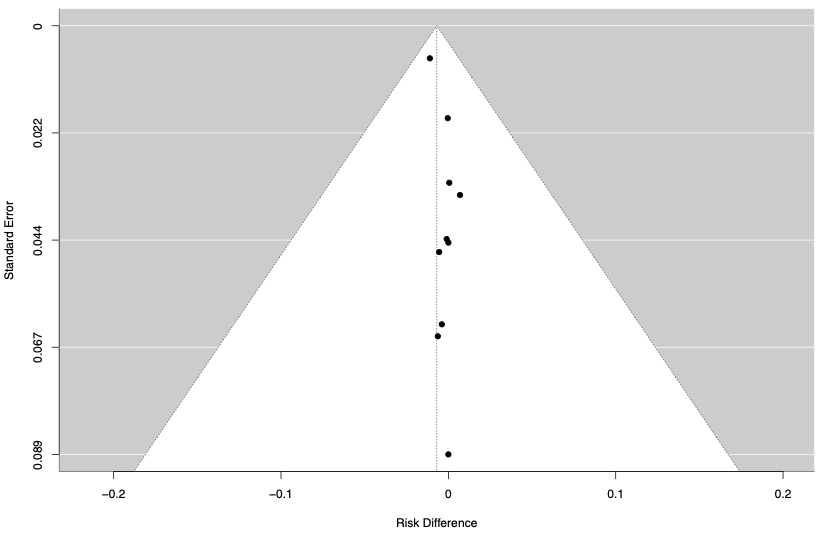

#### 2.30 Funnel plot – Pneumonia

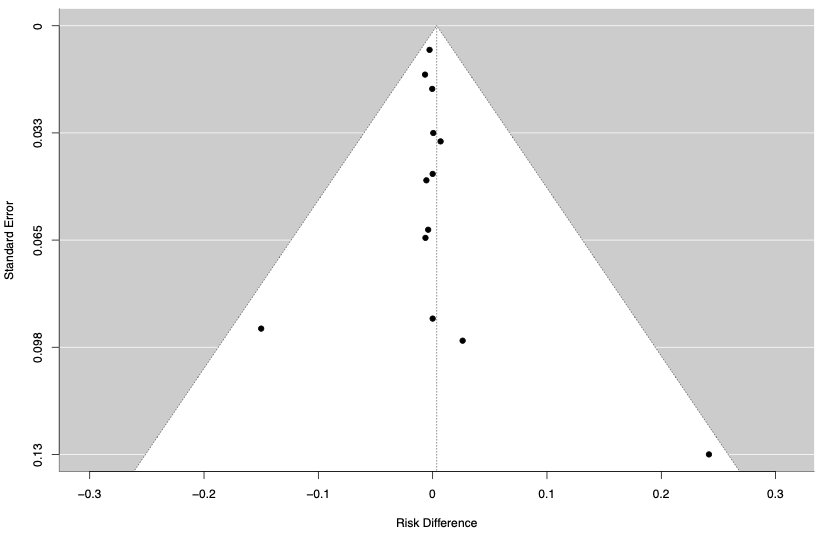

#### 2.31 Funnel plot – Polyuria

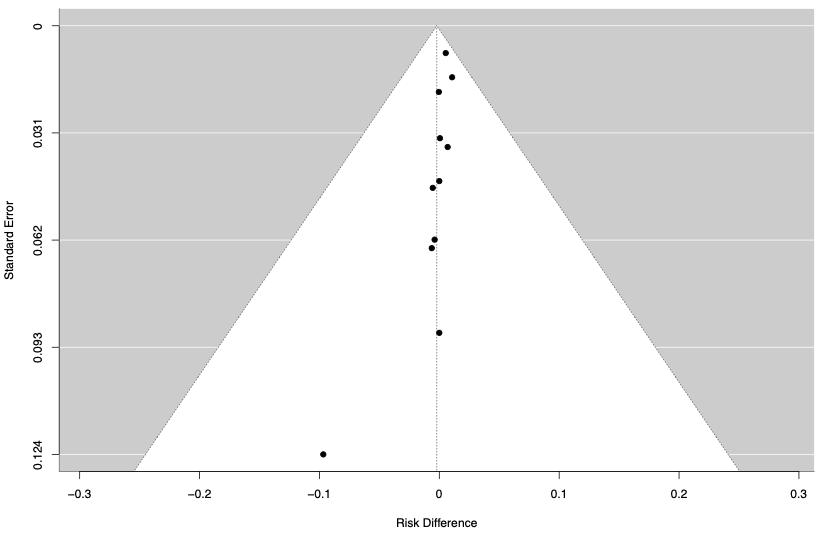

#### 2.32 Funnel plot – Rash or Urticaria

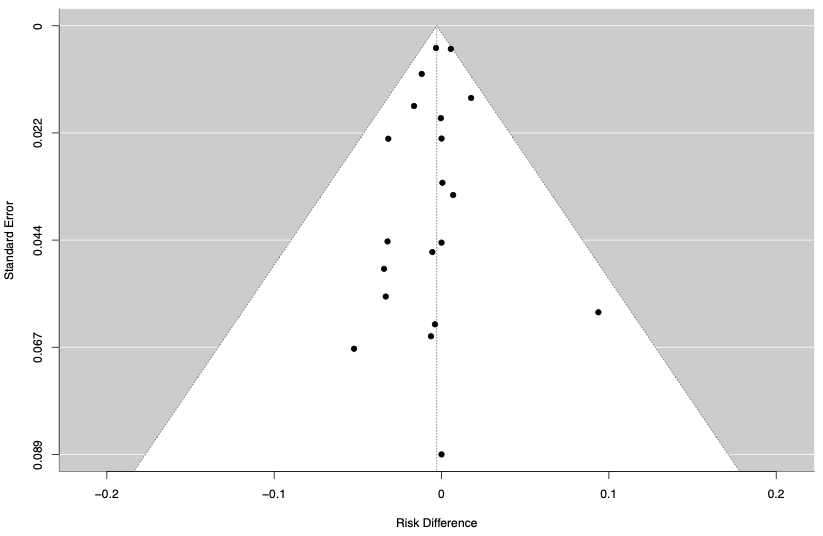

#### 2.33 Funnel plot – Secondary (Opportunistic) Infection

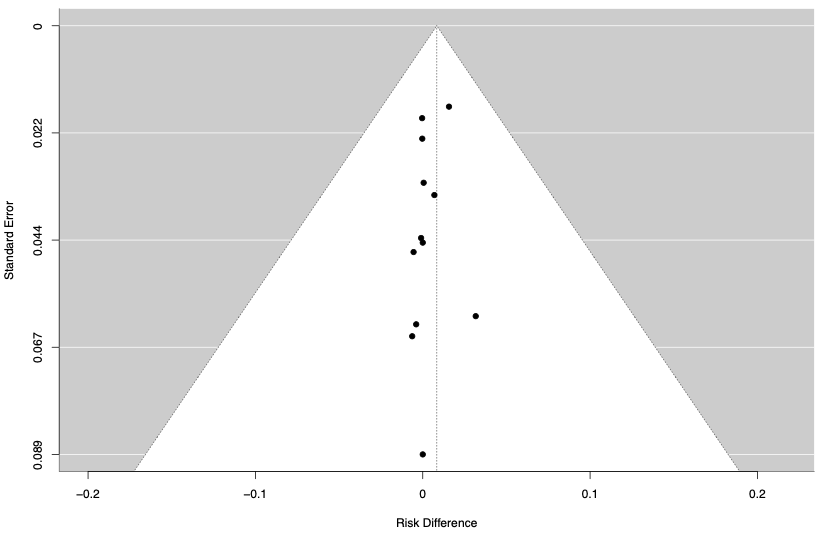

#### 2.34 Funnel plot – Secondary Fever

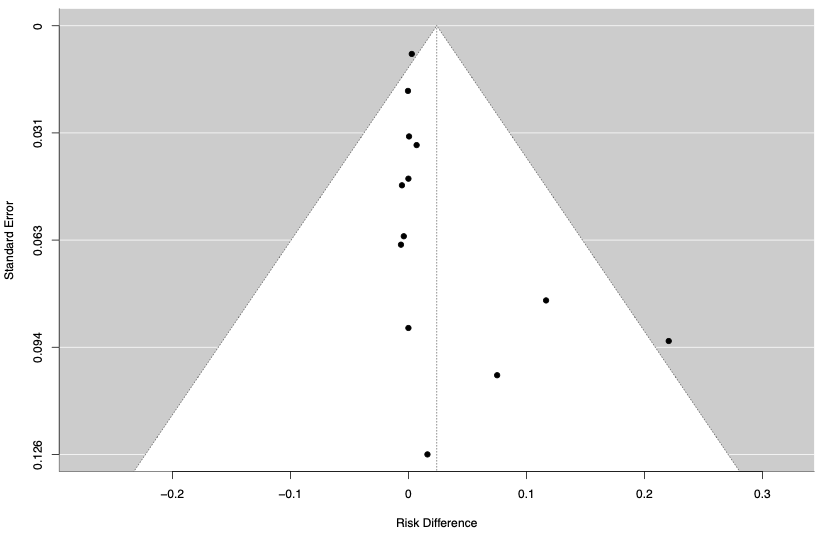

#### 2.35 Funnel plot – Sleep Problems

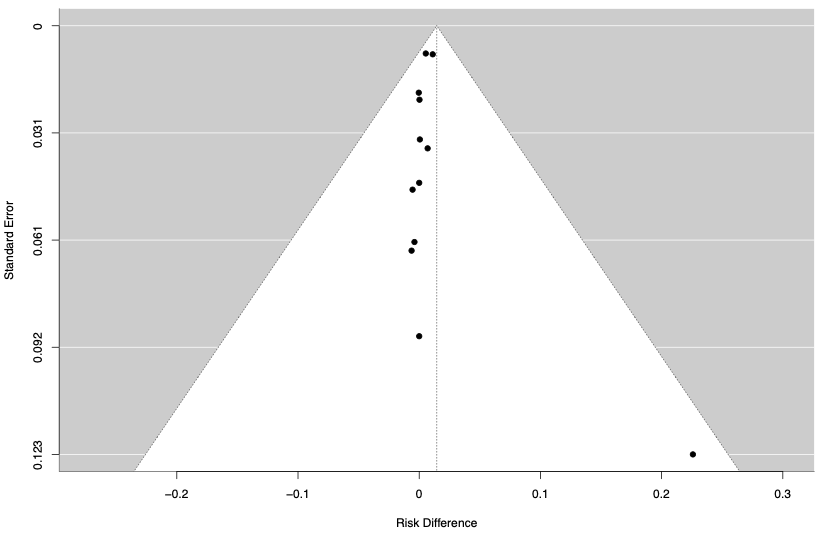

#### 2.36 Funnel plot – Tremor or Hyperactivity

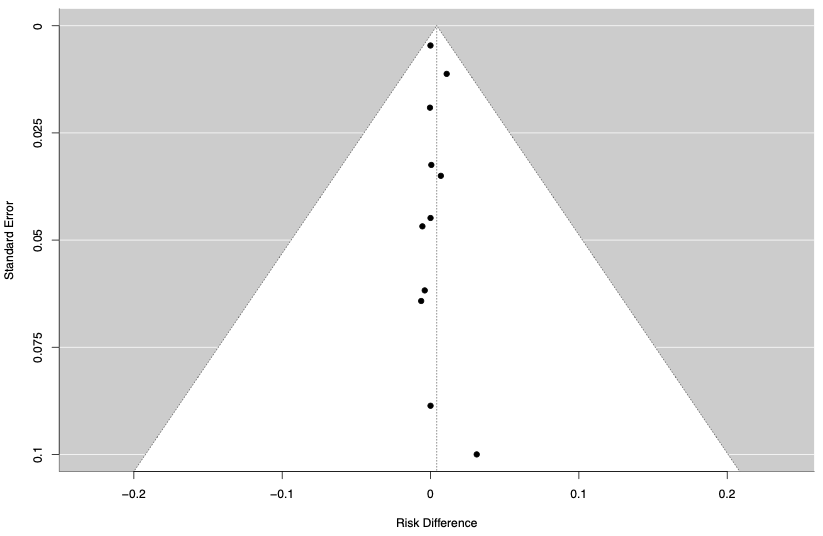

#### 2.37 Funnel plot – Urinary Tract Infection

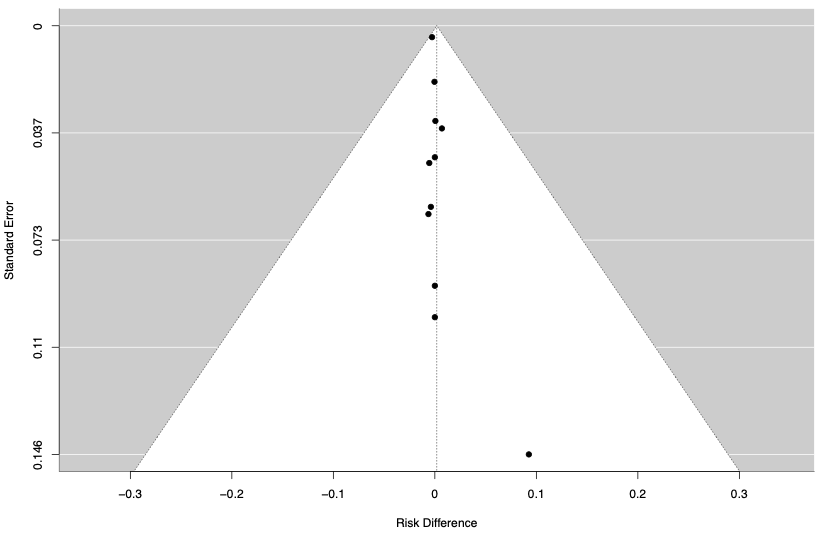

### 3. Method used to capture adverse events in included studies

| **Author, Year** | **Method** |
| --- | --- |
| Babl 2022 | Fourteen days after randomization, the parent/guardian/ participant received a phone call from the study team to assess AEs. One month after randomization, participants attended a visit at the study site where they had been recruited. If the participant was unable to attend the study site, this was completed through videoconferencing. A specialist clinician reviewed the participant for AEs. |
| Bernini 1998 | NR |
| Carcao 2020 | Follow-up AE questionnaires were completed by the parents/children in clinic or by telephone by local study coordinators. |
| Connett 1994 | NR |
| Csonka 2003 | Diary card recordings were made twice daily for 14 days. Children were examined by the study physician 14 to 21 days after the initial ED visit. Diary entries and patient files were reviewed, and patients who did not return for reassessment were contacted by telephone. |
| Dudley 2013 | AEs assessed by clinician ar the end of the 4-week visit. |
| Foster 2018 | AEs were captured at follow-up via completion of a 7-day symptom diary and at a 10-day and 3-month phone call by masked research assistants. During follow-up phone calls, legal guardians were asked for the presence of vomiting or any other reasons for not completing the study drug. |
| Francis 2018 | Legal guardians were provided with a symptom diary to complete at home during the first 5 weeks used to record AEs. |
| Griffin 1994 | NR |
| Harel 2011 | NR |
| Hoffman 1988 | NR |
| Hoke 1992 | NR |
| Huber 2004 | Structured diary was completed by legal guardians to document AEs during 14 days. |
| Jartti 2007 | NR |
| Johnnson 1998 | NR |
| Kanra 1995 | NR |
| King 1994 | NR |
| Kuusela 1988 | NR |
| Kvien 1982 | NR |
| Lebel 1988 | NR |
| Leipzig 1979 | NA |
| Lv 2022 | NR |
| Medeiros 2007 | NR |
| Nagy 2012 | NR |
| Newburger 2007 | NR |
| Ogata 2012 | AEs were documented in the daily medical record reviews by nurses. |
| Olympia 2005 | The child or child’s legal guardian was contacted by telephone daily by the research team from the time of discharge to the time of complete resolution of the sore throat. |
| Panickar 2009 | NR |
| Pierson 1974 | NR |
| Prakash 2006 | NR |
| Qazi 1996 | The presence of AEs, including death,and their possible relation to the administration of the study drugs was assessed daily. These events included secondary fever (defined as recurrence of temperatures higher than 37.8°C after 24hours or more and evidence of blood testing at least three times per week) gastrointestinal bleeding assessed by occult. |
| Ranakusuma 2020 | NR |
| Ruohola 1999 | NR |
| Schaad 1993 | NR |
| Shaikh 2020 | Staff reviewed the child's medical record for any medical care visits since their last study contact for any AEs. If child was seen or had any new symptoms within 2 weeks of taking study product, the symptoms were reported as AEs. After 2 weeks of taking study product, if a child was seen for fever or urinary symptoms but not diagnosed with a urinary tract infection these symptoms were reported as an AE. |
| Shan 2017 | NR |
| Sundel 2003 | Safety of treatment was assessed by the occurrence of prospectively defined aAEs that were documented during daily medical record reviews by the study nurse. |
| Super 1989 | NA |
| Tagarro 2017 | NR |
| Tam 2012 | NR |
| Tassniyom 1993 | NR |
| Unuvar 1999 | NA |
| Usta 2014 | Patients in the study group were monitored for the AEs of the treatment including hyperglycemia, high blood pressure, electrolyte disturbances, and infections. |
| Wang 2022 | NR |

NR: Not reported

### 4. Non-poolable adverse events

| **Author, Year** | **Adverse Events** | **Corticosteroids (events/n)** | **Placebo or usual care (events/n)** |
| --- | --- | --- | --- |
| Babl 2022 | Change in vision | 1/93 | 0/94 |
|  | Hair loss | 0/93 | 2/18 |
| Bernini 1998 | Osteonecrosis | 0/22 | 1/97 |
|  | Psychosis | 0/22 | 6/31 |
| Bjornson 2004 | Bleeding from ear | 0/359 | 1/361 |
|  | Bronchitis | 3/359 | 1/361 |
|  | Dehydration | 1/359 | 0/361 |
|  | Eye discharge | 1/359 | 0/361 |
|  | Nasal discharge | 1/359 | 3/170 |
|  | Respiratory syncytial virus (RSV) infection | 1/259 | 0/94 |
|  | Sinusitis | 0/359 | 1/170 |
|  | Sore throat | 1/359 | 0/361 |
|  | Streptococcal throat infection | 1/359 | 13/31 |
|  | Uncomplicated varicella | 0/359 | 6/31 |
| Carcao 2020 | Allergic reaction | 0/14 | 2/18 |
| Csonka 2003 | Restlessness | 3/113 | 0/41 |
| Dudley 2013 | Bruising/skin problems | 5/180 | 3/170 |
|  | Jaundice | 1/180 | 1/170 |
|  | Malaise | 1/180 | 0/170 |
|  | Nose Bleed | 2/180 | 1/170 |
|  | Stevens Johnson syndrome | 1/180 | 8/275 |
| Francis 2018 | Constipation | 1/179 | 1/170 |
|  | Ear Pain | 1/179 | 1/170 |
|  | Finger infection | 0/179 | 1/170 |
|  | Flushed cheeks | 1/179 | 0/55 |
|  | Frustration | 0/179 | 1/19 |
|  | Parotitis | 0/179 | 0/15 |
| Hoffman 1988 | Bacteremia | 1/19 | 2/19 |
|  | Hypoglycemia | 0/19 | 0/26 |
|  | Pulmonary edema | 1/19 | 0/21 |
| Newburger 2007 | Anaphylaxis | 0/101 | 1/97 |
|  | Hypotension | 5/101 | 5/170 |
|  | Shock | 1/101 | 0/15 |
| Ogata 2012 | Bradycardia | 2/22 | 0/26 |
|  | Hypothermia | 6/22 | 0/170 |
| Olympia 2005 | Swollen legs | 0/57 | 0/21 |
| Pierson 1974 | Hypokalemia | 0/30 | 1/170 |
|  | Muscular weakness | 0/30 | 0/15 |
|  | Psychological reactions | 0/30 | 0/21 |
| Ranakusuma 2020 | Anxiety | 4/31 | 6/31 |
|  | Drowsiness | 23/31 | 13/31 |
|  | Dry mouth | 7/31 | 6/31 |
|  | Weight gain | 13/31 | 2/117 |
| Schaad 1993 | Haemopoietic Abnormatities | 0/60 | 3/21 |
|  | Hepatic dysfunction | 0/60 | 0/97 |
|  | Renal Abonormatities | 0/60 | 1/361 |
| Shaikh 2020 | Fussiness | 25/271 | 2/361 |
| Sundel 2003 | Idiopathic thrombocytopenic purpura | 1/18 | 0/170 |
|  | Rigors | 1/18 | 4/31 |
| Tagarro 2017 | Tranfusion | 1/30 | 1/361 |
| Tam 2012 | Upper respiratory infection | 4/150 | 0/68 |
| Tassniyom 1993 | Abscess at cut-down area | 1/32 | 1/31 |
|  | Gingivitis | 1/32 | 1/361 |
|  | Stiff joint | 2/32 | 1/75 |
| Usta 2014 | Electrolyte Disturbances | 0/42 | 0/41 |

#### 5. Instrument to assess the Credibility of Effect Modification Analyses (ICEMAN) in a meta-analysis of randomized controlled trials - Version 1.1- Outcome: Gastrointestinal Bleeding

**Consider the following important instructions informed by common misapplications of ICEMAN in studies using the instrument**

- Complete a separate credibility assessment per each effect modifier (e.g., age, comorbidity, drug dose, etc.), outcome (e.g., mortality, stroke, duration of hospital stay), time-point (e.g., 3 months, 6 months), and effect measure (e.g. relative risk, risk difference).
- Do not apply ICEMAN if the interaction p-value is 0.1 or larger, i.e., provides very little statistical support for the existence of an effect modification (ICEMAN is designed to address the possible claim of an effect modification rather than the claim of no effect modification).
- Response options on the left indicate definitely or probably reduced credibility, response options on the right probably or definitely increased credibility
- Completely unclear should be interpreted as probably reduced credibility.
- To ensure transparency, provide a supporting comment under each question that provides a rationale for the rating.
- To ensure transparency, provide a copy of the completed ICEMAN instrument in the supplement of your article.

| **CREDIBILITY ASSESSMENT** | | | |
| --- | --- | --- | --- |
| **Essential preliminary considerations to define the possible effect modification of interest** | | | |
| State a single candidate effect modifier (e.g., age or comorbidity): Route (Intravenous or Intramuscular vs Oral) | | | |
| Was the effect modifier measured before or at randomization? [ **X**] yes, continue [ ] no, stop here and refer to manual for further instructions | | | |
| State a single outcome and time-point (e.g., mortality at 1 year follow-up): Gastrointestinal Bleeding, latest follow-up | | | |
| State a single effect measure (e.g., relative risk or risk difference): Risk Diffence | | | |
| **1: Is the analysis of effect modification based on comparison within rather than between trials?** | | | |
| [ **X** ] Completely between | [ ] Mostly between or unclear | [ ] Mostly within | [ ] Completely within |
| *Subgroup analysis or meta-regression comparing overall effects of each individual trial. This is typical for aggregate data meta-analysis.* | *Subgroup analysis or meta-regression with most information coming from overall effects, but some trials providing within-trial subgroup information* | *Most trials providing within-trial subgroup information; or individual participant data analysis that combines within and between trial information* | *All trials providing within-trial subgroup information or individual participant data; and the analysis separates within from between trial information, e.g., meta-analysis of interactions* |
| Comment: NA | | | |
| **2: For within-trial comparisons, is the effect modification similar from trial to trial?** [X] Not applicable: no or one within-RCT comparison | | | |
| [ ] Definitely not similar | [ ] Probably not similar or unclear | [ ] Mostly similar | [ ] Definitely similar |
| *Effect modification reported for two or more trials and clearly different directions* | *Effect modification not reported for individual trials or too imprecise to tell* | *Effect modification reported for two or more trials, mostly similar in direction, but considerable differences in magnitude* | *Effect modification reported for two or more trials, similar in direction, only some differences in magnitude* |
| Comment: NA | | | |
| **3: For between-trial comparisons, is the number of trials large?** [ ] Not applicable: no between RCT comparison | | | |
| [ ] Very small | [ ] Rather small or unclear | [X] Rather large | [ ] Large |
| *1 or 2 or in smallest subgroup; 5 or less in continuous meta-regression* | *3-4 in smallest subgroup; 6-10 in continuous meta-regression* | *5-9 in smallest subgroup; 11 to 15 in continuous meta-regression* | *10 or more in smallest subgroup; more than 15 in continuous meta-regression* |
| Comment: NA | | | |
| **4: Was the direction of effect modification correctly hypothesized a priori?** | | | |
| [ ] Definitely no | [ ] Probably no or unclear | [ ] Probably yes | [ **X**] Definitely yes |
| *Clearly post-hoc or results inconsistent with hypothesized direction or biologically very implausible* | *Vague hypothesis or hypothesized direction unclear* | *No prior protocol available but unequivocal statement of a priori hypothesis with correct direction of effect modification* | *Prior protocol available and includes correct specification of direction of effect modification, e.g., based on a biologic rationale* |
| Comment: NA | | | |
| **5: Does a test for interaction suggest that chance is an unlikely explanation of the apparent effect modification?** (consider irrespective of number of effect modifiers) | | | |
| [ ] Chance a very likely explanation | [ ] Chance a likely explanation or unclear | [ **X**] Chance may not explain | [ ] Chance an unlikely explanation |
| *Interaction or meta-regression p-value >0.05* | *Interaction or meta-regression p-value ≤0.05 and >0.01, or no test of interaction reported and not computable* | *Interaction or meta-regression p-value ≤0.01 and >0.005* | *Interaction or meta-regression p-value ≤0.005* |
| Comment: NA | | | |
| **6: Did the authors test only a small number of effect modifiers or consider the number in their statistical analysis?** | | | |
| [ ] Definitely no | [ ] Probably no or unclear | [ ] Probably yes | [ **X**] Definitely yes |
| *Explicitly exploratory analysis or large number of effect modifiers tested (e.g., greater than 10) and multiplicity not considered in analysis* | *No mention of number or 4-10 effect modifiers tested and number not considered in analysis* | *No protocol available but unequivocal statement of 3 or fewer effect modifiers tested* | *Protocol available and 3 or fewer effect modifiers tested or number considered in analysis* |
| Comment: | | | |
| **7: Did the authors use a random effects model?** | | | |
| [ **X**] Definitely no | [ ] Probably no or unclear | [ ] Probably yes | [ ] Definitely yes |
| *Fixed (or common) effect or fixed effects model explicitly stated* | *Probably fixed effect(s) model* | *Probably random (or mixed) effects* | *Random (or mixed) effects explicitly stated* |
| Comment: | | | |
| **8: If the effect modifier is a continuous variable, were arbitrary cut points avoided?** [X] not applicable: not continuous | | | |
| [ ] Definitely no | [ ] Probably no or unclear | [ ] Probably yes | [ ] Definitely yes |
| *Analysis based on exploratory cut point(s), e.g., picking cut point associated with highest interaction p-value* | *Analysis based on cut point(s) of unclear origin* | *Analysis based on pre-specified cut point(s), e.g., suggested by prior RCT* | *Analysis based on the full continuum, e.g., assuming a linear or logarithmic relationship* |
| Comment: NA | | | |
| **9 Optional: Are there any additional considerations that may increase or decrease credibility?** (manual section 3.9) [X] not applicable | | | |
|  | [ ] Yes, probably decrease | [ ] Yes, probably increase | |
| Comment: NA **10: How would you rate the overall credibility of the proposed effect modification?** The overall rating should be driven by the items that decrease credibility. The following provides a sensible strategy: - All responses definitely or probably decrease credibility or unclear 🡪 very low - **Two or more responses definitely decrease credibility 🡪 maximum usually low even if all other responses satisfy credibility criteria** - One response definitely decreases credibility 🡪 maximum usually moderate even if all other responses satisfy credibility criteria - Two responses probably decrease credibility 🡪 maximum usually moderate even if all other responses satisfy credibility criteria - No response options definitely or probably decrease credibility 🡪 high very likely Place a mark on the continuous line (or type “x” in editable version) \| \| \| \| \|; \| Very low credibility \| ***Low credibility*** \| Moderate credibility \| High credibility \|; \| Minimal to no support for effect modification; Use overall effect for each subgroup \| ***Some but insufficient support for effect modification;*** ***Use overall effect for each subgroup but note remaining uncertainty*** \| Likely effect modification; Use separate effects for each subgroup but note remaining uncertainty \| Very likely effect modification; Use separate effects for each subgroup \|; Comment: NA \| \| \| \| \| | | | |

### 6. Sensitivity Analyses – Peto Odds Ratio

#### 6.1 Serious Adverse Events (SAE)

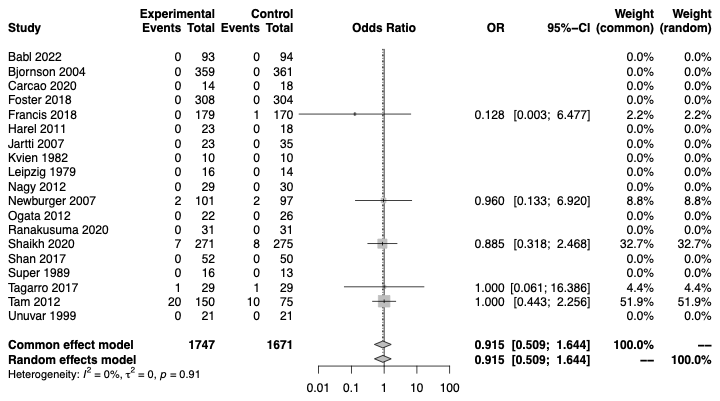

#### 6.2 Adverse events leading to discontinuation

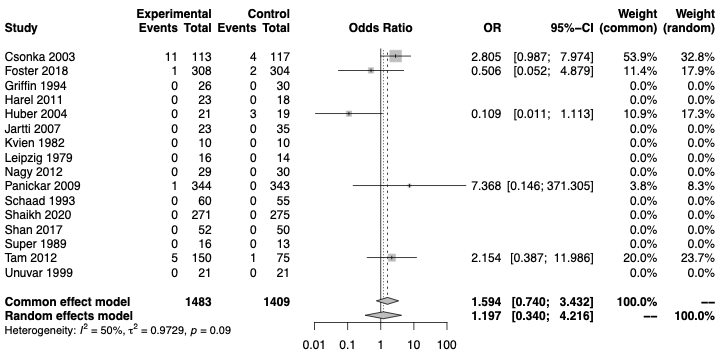

#### 6.3 Gastrointestinal Bleeding

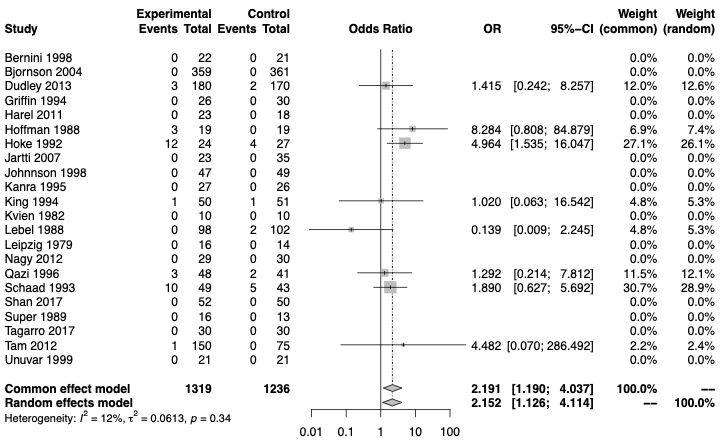

#### 6.4 Sleep Problems

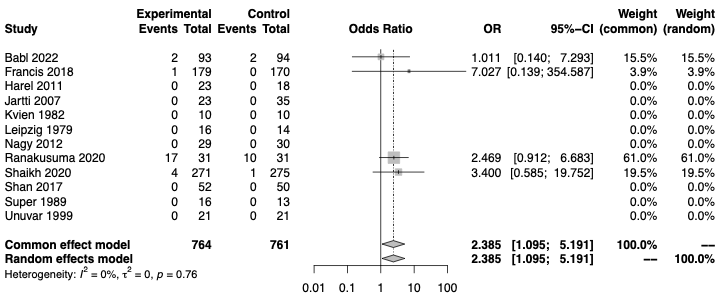

#### 6.5 Change in Behaviour

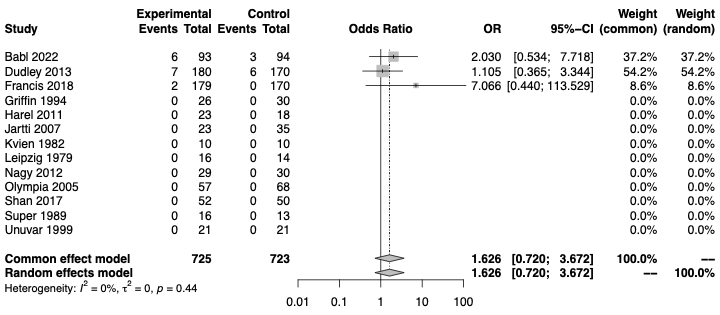

#### 6.6 Hyperglycemia

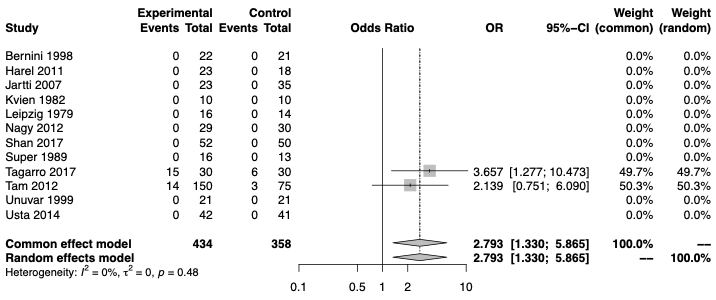

### 7. Subgroup Analysis –

#### 7.1. Gastrointestinal bleeding (Subgroup - route: intravenous/intramuscular vs oral)

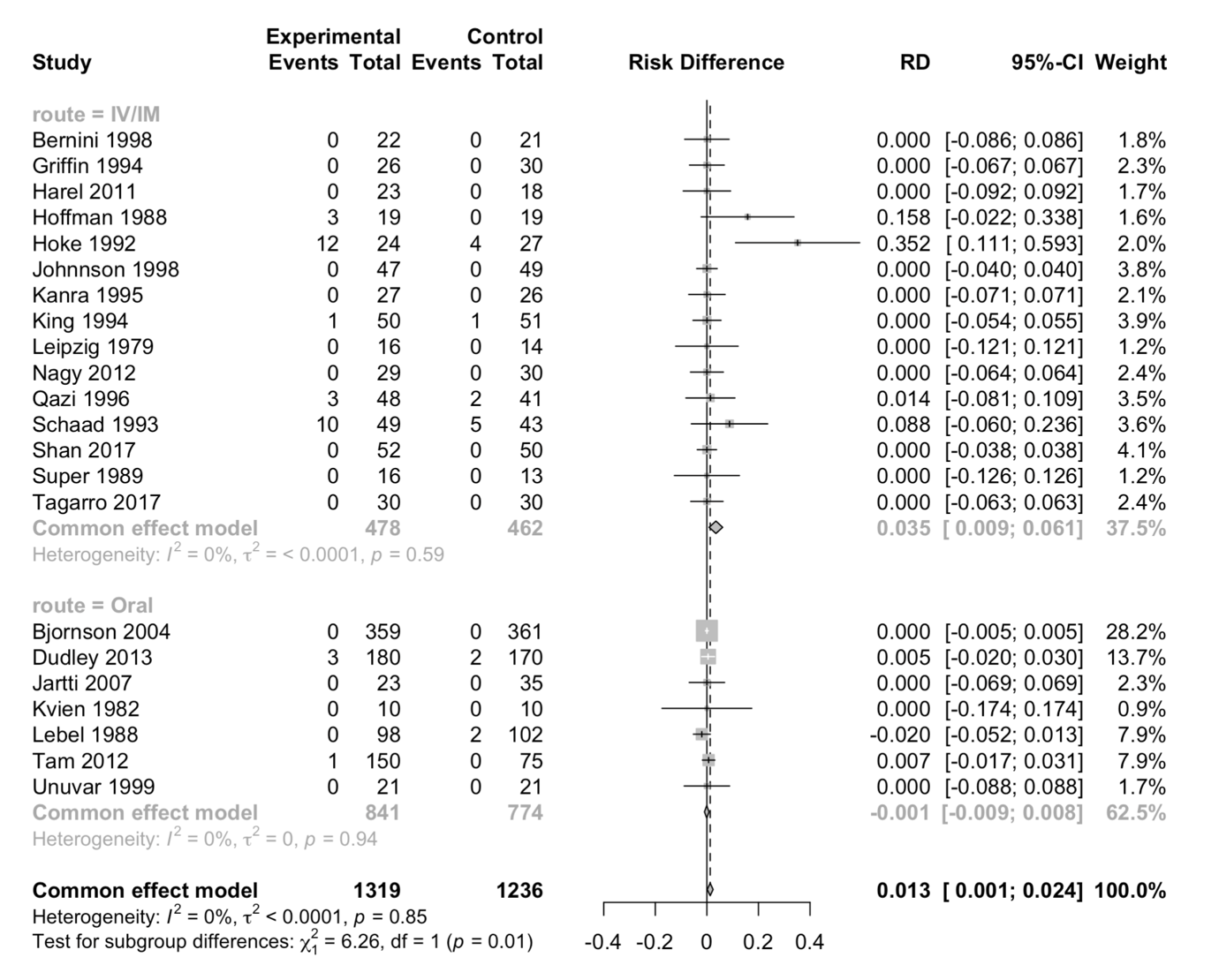

CI: confidence interval IV/IM: intravenous/intramuscular; RD: risk difference

#### 7.2 Gastrointestinal bleeding (subgroup: condition)

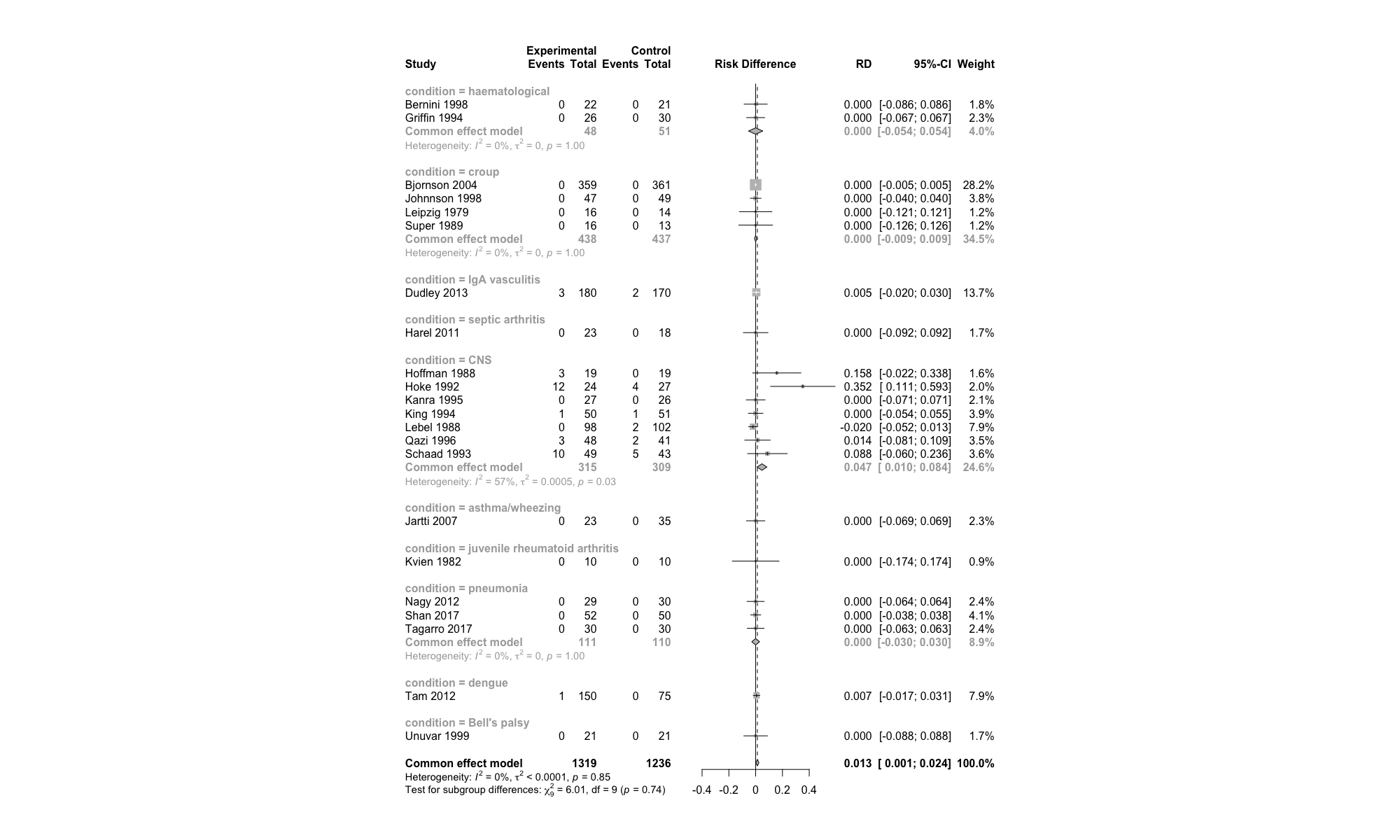

CNS= central nervous system; CI= confidence interval RD=risk difference

#### 7.3 Sleep problems (subgroup: condition)

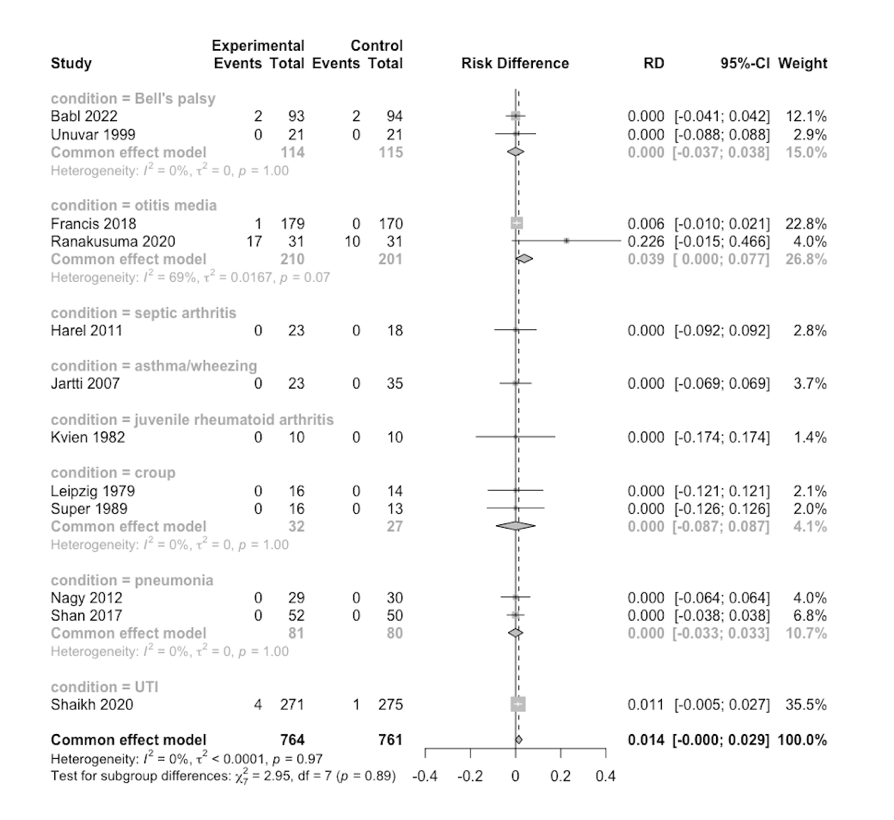

#### 7.4 Change in behavior (subgroup: condition)

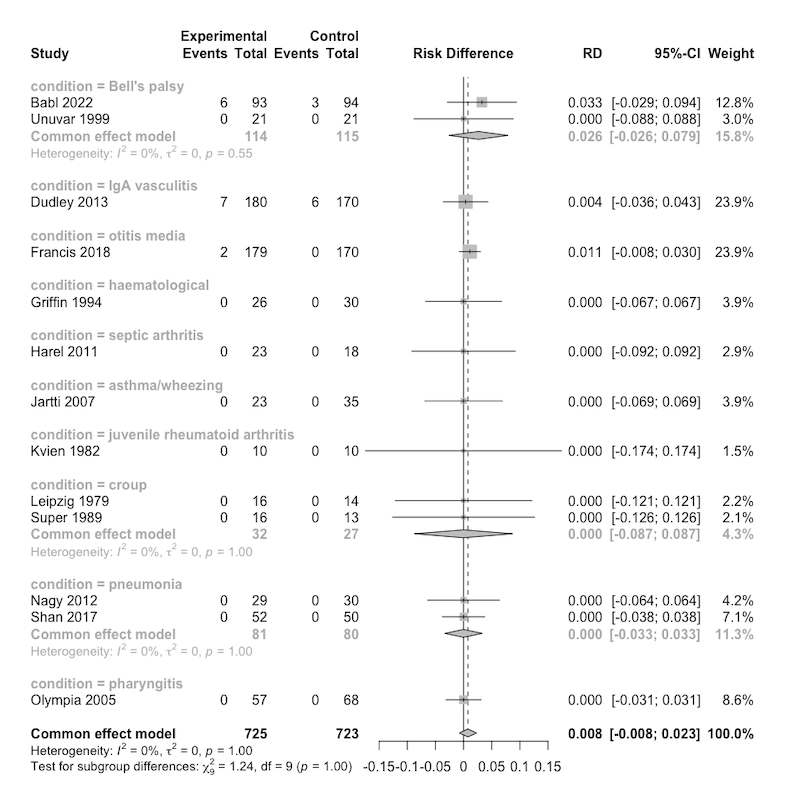

#### 7.5 hyperglycemia (subgroup: condition)

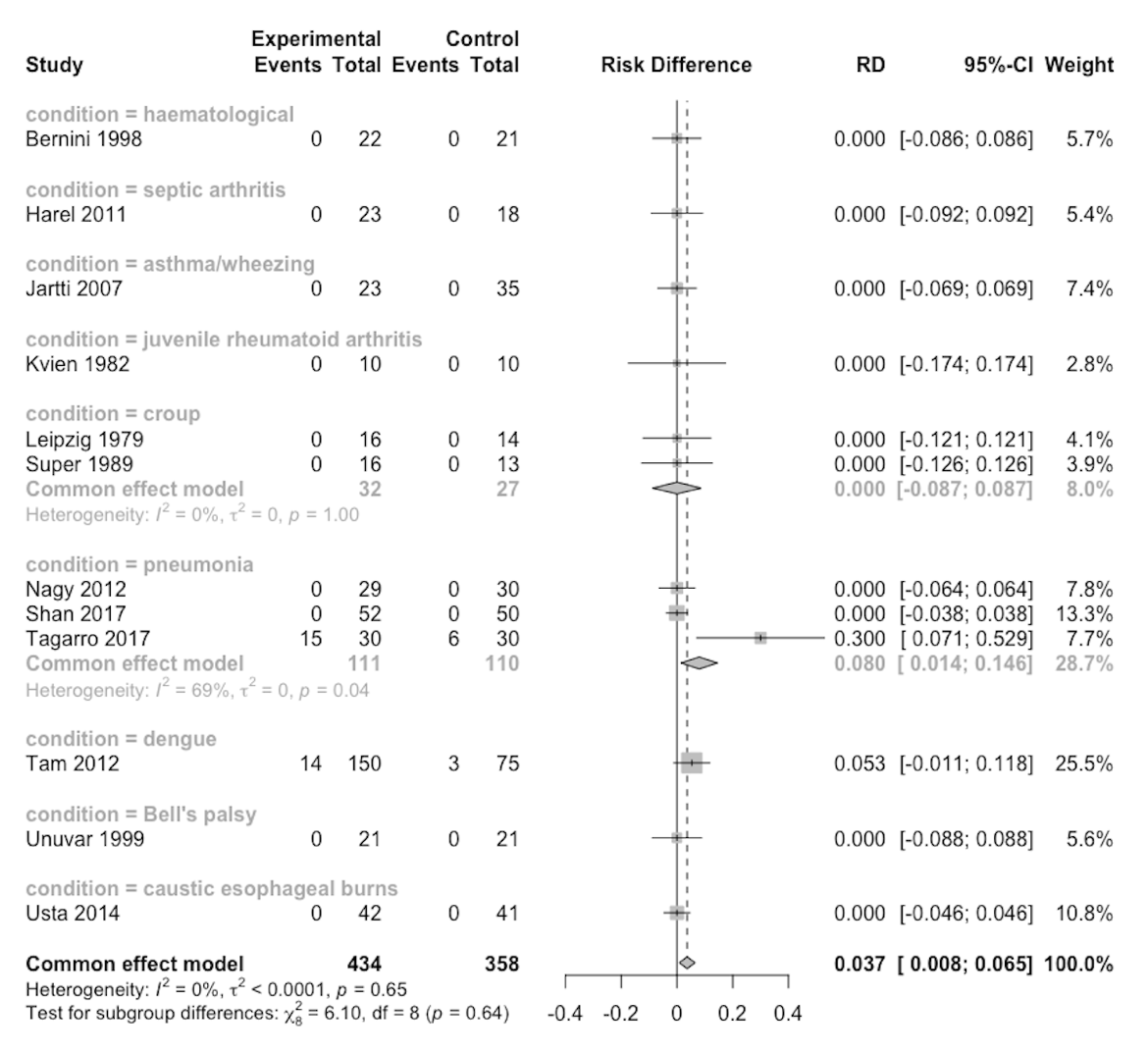

#### 7.6 Serious Adverse Events (subgroup: condition)

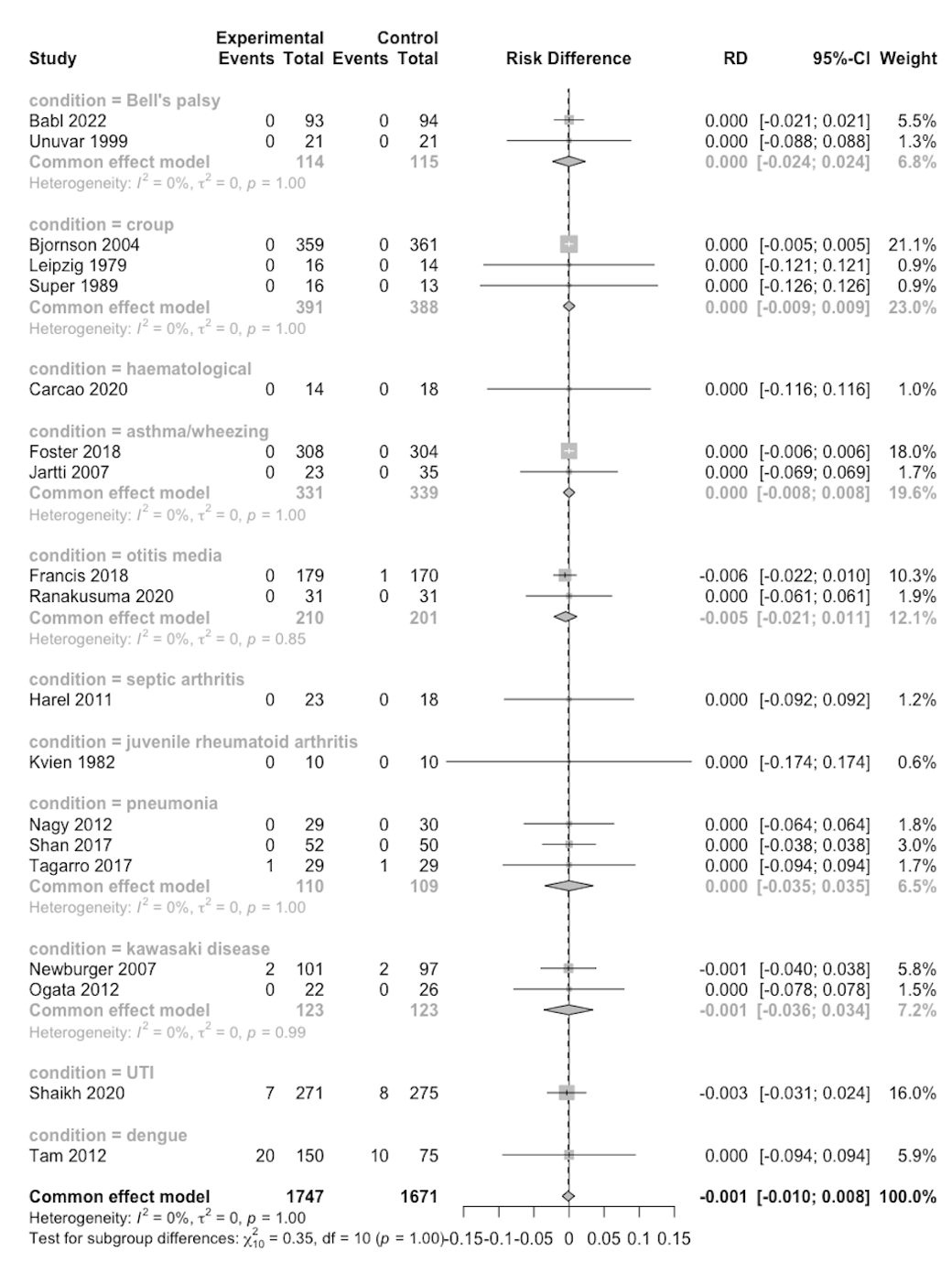

#### 7.7 Adverse events leading to discontinuation (subgroup: condition)

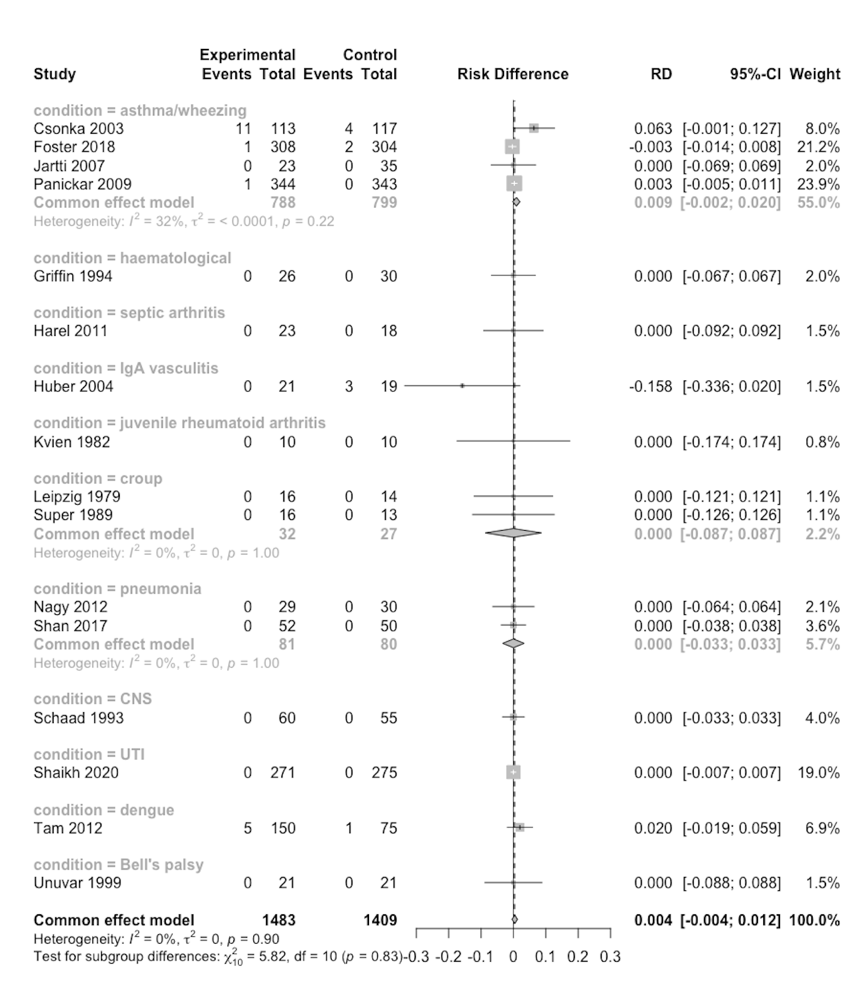

### 8. Previous Systematic Reviews

| Author, Year | Main findings | Limitations |
| --- | --- | --- |
| Aljebab, 2016 ­­­ | - Among the most frequent AEs with short-course oral corticosteroids in children were vomiting, behavioral changes and sleep disturbance were among the most frequent AEs. - Increase in infection rates | - Limited to oral interventions, - Exclusion of studies at high risk of bias - Fails to assess the certainty of evidence - Included observational studies and did not perform meta-analyses - Inclusion of observational studies - No meta-analyses performed |
| Fernandes, 2019 | - Among young children (<6 years) with respiratory conditions using inhaled or systemic corticosteroids (<15 days), there was little to difference on the risk of secondary infections. | - Fails to assess the certainty of evidence - Limited number of clinical conditions - Inclusion of observational studies |
| Chaudhuri, 2024 | - In children and adults with sepsis, acute respiratory distress syndrome and community-accquired pneumonia, corticosteroids are associated with hyperglycemia and hypernatremia with no effect on gastrointestinal bleeding or secondary infections. | - Limited number of clinical conditions |
| Kulkarni, 2022 | - In adults using systemic corticosteroid therapy, is associated with increased risk of metabolic adverse events (e.g., hyperglycemia, hypertension, weight gain and hyperlipidaemia) | - Limited number of adverse events studied - Fails to assess the certainty of evidence |
| Efraij, 2018 | - The use of oral corticosteroids in adults with asthma is associated with complications such as diabetes, hypertension and bone and muscle complications. | - Limited number of clinical conditions - Fails to assess the certainty of evidence |
